# Succinate dehydrogenase B (SDHB) overexpression with enzymatic dysfunction defines a distinct subtype of undifferentiated pleomorphic sarcoma

**DOI:** 10.1101/2025.06.23.25330132

**Authors:** Miguel Esperança-Martins, Hugo Vasques, Manuel Sokolov Ravasqueira, Filipa Santos, Filipa Fonseca, António Syder Queiroz, João Boavida, Daniel Martins Jordão, Joaquim Soares do Brito, Patrícia Corredeira, Marta Martins, Ângela Afonso, Jorge Antonio López, Richard S.P.Huang, Cecília Melo Alvim, Isabel Fernandes, Dolores López Presa, Maria Manuel Lemos, Brian A. Van Tine, Alliny Bastos, Sandra Casimiro, Nuno Abecasis, Luís Costa, Emanuel Gonçalves, Iola Duarte, Sérgio Dias

## Abstract

Undifferentiated pleomorphic sarcoma (UPS) remains one of the most clinically aggressive and poorly characterized soft tissue sarcoma (STS) subtypes. To uncover distinctive molecular traits for UPS, a multi-omics analysis of UPS compared to leiomyosarcoma (LMS), and liposarcoma (LPS) was performed. Transcriptomic profiling revealed that UPS exhibits overexpression of genes encoding succinate dehydrogenase (SDH) subunits, particularly SDHB, SDHC, and SDHD, distinguishing it from LMS and dedifferentiated LPS (DDLPS). This finding was validated using the TCGA-SARC dataset. High SDHB expression in UPS was significantly associated with shorter overall survival (OS) and shorter OS from the date of first metastasis. Immunohistochemistry validated elevated SDHB protein levels in UPS and LMS relative to DDLPS. Despite overexpression of SDH subunits, metabolomic profiling demonstrated a significantly higher succinate-to-fumarate ratio in UPS, suggesting functional impairment of SDH enzymatic activity potentially due to post-translational modifications, altered assembly of SDH subunits, or imbalanced tricarboxylic acid (TCA) flux. This paradoxical phenotype of SDH overexpression with enzymatic dysfunction defines a unique molecular and metabolic subtype of UPS with prognostic significance. Recognition of this distinct SDH-associated molecular and metabolic phenotype provides insight into UPS pathogenesis, identifies a potential novel prognostic biomarker, and suggests a new avenue for metabolic-targeted therapy.

## Introduction

Soft tissue sarcomas (STS) are a group of rare and heterogeneous malignancies arising from mesenchymal tissues, accounting for approximately 1% of adult cancers and up to 15% of pediatric solid tumors [1]. Their rarity, biological diversity and heterogeneity, with over 100 recognized histopathological subtypes, pose significant challenges to understanding their molecular pathogenesis and to discovering and developing effective targeted therapies [2]. While subtype-specific genetic alterations have been identified in certain sarcomas, such as KIT/PDGFRA mutations in gastrointestinal stromal tumors or the MDM2/CDK4 amplification in dedifferentiated liposarcomas (DDLPS), the majority of high-grade STS lack clearly defined and targetable molecular drivers [3]. As a result, clinical outcomes for advanced or recurrent disease remain poor, and therapeutic advances have been limited for most of the high-grade sarcomas [4,5].

Among adult STS, undifferentiated pleomorphic sarcoma (UPS), and L-type sarcomas (leiomyosarcoma (LMS), and liposarcoma (LPS)) are the three most prevalent histopathological subtypes [6]. UPS represents ∼17% of STS, and has 5- and 10-year overall survival (OS) rates of approximately 60% and 48%, respectively, with high rates of local (14.1%) and distant (7.8%) recurrence [7,8,9]. Despite its significant prevalence among STS and dismal prognosis, UPS remains poorly characterized at the molecular level outside of the TGCA [10]. Previous studies have suggested similarities in gene expression and immunohistochemical profiles between UPS and LMS, suggesting that they share common oncogenic pathways and may correspond to different stages of the same oncologic entity [11-13], but these observations have not delineated a unique molecular or metabolic feature for UPS.

Recent evidence has highlighted the central role of tumor metabolism in driving oncogenesis, disease progression, and therapeutic resistance [14-18]. Cancer cells undergo metabolic adaptations to meet the bioenergetic and biosynthetic demands of rapid proliferation [19]. Mitochondrial enzymes, particularly those involved in the tricarboxylic acid (TCA) cycle and oxidative phosphorylation (OXPHOS), play a critical role in this adaptation [19,20]. Succinate dehydrogenase (SDH), also known as mitochondrial complex II, is at the interface of the TCA cycle and the electron transport chain (ETC), oxidizing succinate to fumarate while transferring electrons to ubiquinone [21]. Inactivating mutations or epigenetic silencing of SDH subunits leading to succinate accumulation, are well-established oncogenic mechanisms in several tumor types, including gastrointestinal stromal tumors, paragangliomas, and renal cell carcinomas [21,22]. In these contexts, SDH deficiency and succinate accumulation promote tumorigenesis through stabilization of hypoxia-inducible factors (HIFs) and widespread epigenetic dysregulation [23].

In contrast to these known SDH-deficient neoplasms, the role of SDH in UPS and other high-grade sarcomas has not been systematically investigated. Whether UPS harbors alterations in SDH expression or activity, and whether such alterations contribute to its aggressive clinical behavior, remains unknown. To address these gaps, the present study employed an integrative multi-omics approach to define the molecular and metabolic biology of UPS. These findings provide evidence that SDH overexpression and enzymatic dysfunction in UPS has prognostic significance and potential therapeutic implications.

## Materials and Methods

### Gene expression analysis

#### Tissue cohort

The gene expression cohort consisted of 102 formalin-fixed paraffin-embedded (FFPE) tumor samples from 101 patients with a high-grade (grade 3) STS, including 51 patients with a UPS, 25 patients with a high-grade LMS, and 25 patients with a dedifferentiated LPS (DDLPS). Clinical, pathological, and treatment characteristics have been described previously [24].

#### DNA and RNA sequencing

DNA and RNA were co-extracted from FFPE sections and analyzed at Foundation Medicine Inc. (Cambridge, MA, USA), a CLIA-certified, CAP-accredited, and New York State–approved laboratory. DNA sequencing was performed using the FoundationOne®CDx assay (F1CDx) [25], and RNA sequencing (RNA-seq) was performed using the research-use–only FoundationOne®RNA assay (F1RNA) [26], as previously described [24].

#### Quality Control

Of the 102 tumor samples (51 UPS, 25 LMS and 26 DDLPS), 79 passed F1CDx quality control, and 75 passed F1RNA quality control (QC). Samples failing F1RNA QC were predominantly archival, with a median age of 4.7 years. One additional outlier was excluded from the RNA-seq expression analysis based on principal component analysis (PCA). A total of 74 samples (43 UPS, 15 LMS and 16 DDLPS) were included in downstream gene expression analyses.

#### RNA-seq-based differential gene expression analysis

Computational analyses were conducted in R v4.4.0. Lowly expressed genes were removed using the edgeR v4.2.1 [27] filterByExpression method, followed by Voom normalization. Differential gene expression (DGE) analyses between histopathological subtypes were performed using limma v3.60.4 [28], and p-values were adjusted for multiple testing with the Benjamini–Hochberg False Discovery Rate (FDR). Genes with an adjusted p-value ≤ 0.05 were considered differentially expressed. The concordance between metabolic pathways and genes included in F1RNA was identified using KEGG pathway annotations [29].

#### Clinical significance of differentially expressed genes (study cohort)

Survival analyses were performed, within and between each histopathological subtype and for each gene baited for expression and included in F1RNA, in R using the survival package v3.7.0 [30] for Overall Survival (OS), Metastasis-Free Survival (MFS), Recurrence-Free Survival (RFS), Progression-Free Survival (PFS), and OS from the date of metastasis. Samples were stratified into high or low expression groups for specific genes based on whether that gene expression levels were above or below the mean expression of the gene. Log-rank tests were then applied for each gene. Over- representation analysis (ORA) [31] was conducted on differentially expressed genes (DEGs), stratified by directionality of expression (over- or underexpressed). This ORA was performed for sets of gene whose expression was significantly associated with survival (log-rank p < 0.05), further dividing by favorable vs. unfavorable prognostic associations.

#### External validation using TCGA-SARC (Independent Cohort)

The TCGA-SARC dataset [32] was used as an independent validation cohort, filtered to only include UPS, LMS, and DDLPS samples based on the updated histopathologic classification. RNA-seq data were processed and analyzed and survival analysis and ORA were performed similarly to which was done for the study cohort.

To assess concordance between cohorts, we evaluated whether the direction of expression changes between histopathological subtypes was consistent across cohorts. For each pairwise comparison (UPS vs LMS, UPS vs DDLPS, LMS vs. DDLPS), we identified genes with an adjusted p-value < 0.05 in both cohorts and intersected these sets. Within the overlapping gene set, we then compared the direction of expression changes by analyzing log fold-change (logFC) values to determine whether genes were consistently over expressed (logFC > 0) or under expressed (logFC < 0) in both datasets. Statistical significance of the directional concordance was assessed using a hypergeometric test. In addition to DGE concordance, we investigated the consistency of survival associations. Focusing on the UPS subtype, we selected genes significantly associated with OS, defined by a log-rank test p-value < 0.05, in each cohort. For each gene, we determined whether higher expression was associated with a favorable or unfavorable prognosis. We then intersected the sets of significant genes from the study cohort and TCGA and compared the directionality of their survival associations—i.e., whether high expression was consistently linked to either improved or worsened survival across datasets. This concordance was also evaluated using a hypergeometric test, following the same logic as in the DGE comparison.

### Immunohistochemical evaluation of SDHB protein levels

All FFPE tumor sections were originally submitted to histopathological review by a sarcoma pathologist at the Pathology Department of Instituto Português de Oncologia de Lisboa Francisco Gentil (IPOLFG) (Lisboa, Portugal). All cases were fixed in 10% formalin. SDHB immunohistochemistry was performed on deparaffinized sections using a standard avidin–biotin–peroxidase complex method in an automated platform (BenchMark ULTRA, Ventana, Tucson, AZ, USA). FFPE blocks were qualitatively analyzed for the expression of a rabbit monoclonal anti-SDHB antibody (clone EP288, ref MAD-000739QD, Vitro Master Diagnóstica ™, Granada, Spain). Gastrointestinal stromal tumor tissue sections served as positive and negative controls. Two pathologists, blinded to clinical, pathological and molecular data, scored staining intensity. Definite cytoplasmic granular staining was considered positive. Immunostaining intensity was classified using a newly-developed three-tier system: Score 1: focal and weak staining (<10% of cells); Score 2: diffuse and intermediate staining (10–90% of cells); Score 3: diffuse and strong staining (>90% of cells). Additionally, cases lacking unequivocal cytoplasmic granular staining were considered negative and cases lacking a definite positive internal control were considered non-informative.

### Nuclear Magnetic Resonance (NMR) Metabolomics

#### Tissue cohort

Fresh frozen tumor and normal tissue samples were collected from 16 STS patients undergoing surgery between 2021 and 2023 at two Portuguese sarcoma centers: IPOLFG and Unidade Local de Saúde de Santa Maria (ULSSM). Tumor samples (n=15) included three UPS, three LMS, two LPS (one DDLPS, one myxoid LPS), and additional rare sarcomas (dermatofibrosarcoma protuberans, solitary fibrous tumor, synovial sarcoma, chondrosarcoma, clear-cell sarcoma-like tumor of the gastrointestinal tract, endometrial stromal sarcoma, and adamantinoma). Histopathologic diagnoses were confirmed by a dedicated sarcoma pathologist at IPOLFG. Clinical data were obtained from institutional records and national electronic health records. Data were anonymized and follow-up was closed on April 21, 2025.

#### Metabolite extraction from fresh tissue

Samples were first macerated using an agate mortar and pestle under liquid nitrogen and then stored at –80 °C. Metabolites were extracted from ∼50 mg tissue by sequential addition of 650 µL of cold methanol 80%, 520 µL of cold chloroform, and 260 µL of cold Milli-Q water, followed by vortexing for 1 minute after each addition. After the final addition of water and vortexing, samples were incubated at 4 °C for 20 minutes, centrifuged at 3,000 ×g for 10 minutes. A 500 µL aliquot of the resulting upper aqueous phase was collected, dried in a speed vacuum concentrator (Centrivap, model 73100, Labconco, Kansas City, MO, USA), and stored at –80 °C until further analysis.

#### ^1^H NMR data acquisition and analysis

Dried extracts were reconstituted in deuterated phosphate buffer (pH 7.4) with 0.1 mM TSP-d4 and transferred to 3 mm NMR tubes. Spectra were acquired at 298 K on a Bruker Avance III HD 500 MHz spectrometer using a standard noesypr1d sequence with water suppression, 512 scans, and processed in TopSpin 4.0.3. 2D ^1^H–^1^H TOCSY and J-resolved spectra were acquired for selected samples. Metabolite identification was based on Chenomx and BBIOREFCODE reference libraries.

Spectral integration and total area normalization were performed in Amix-Viewer 3.9.15. Data normality was assessed using the Shapiro–Wilk test. For comparisons among three groups, one-way ANOVA with Tukey’s post hoc test (parametric) or Kruskal–Wallis with Dunn’s post hoc test (non-parametric) was used. Two-group comparisons were made using unpaired Student’s t-test with Welch’s correction or the Mann–Whitney test, as appropriate. A p-value < 0.05 was considered statistically significant.

## Results

### Clinical and pathologic characteristics of the study cohort

102 FFPE tumor samples from 101 patients with STS (51 UPS, 25 high-grade LMS, and 26 DDLPS) were studied (Table 1). One patient contributed with two independent samples. The cohort included mostly primary high-grade STS from diverse anatomical sites, representing the topographical heterogeneity of high-grade STS.

**Table 1.**
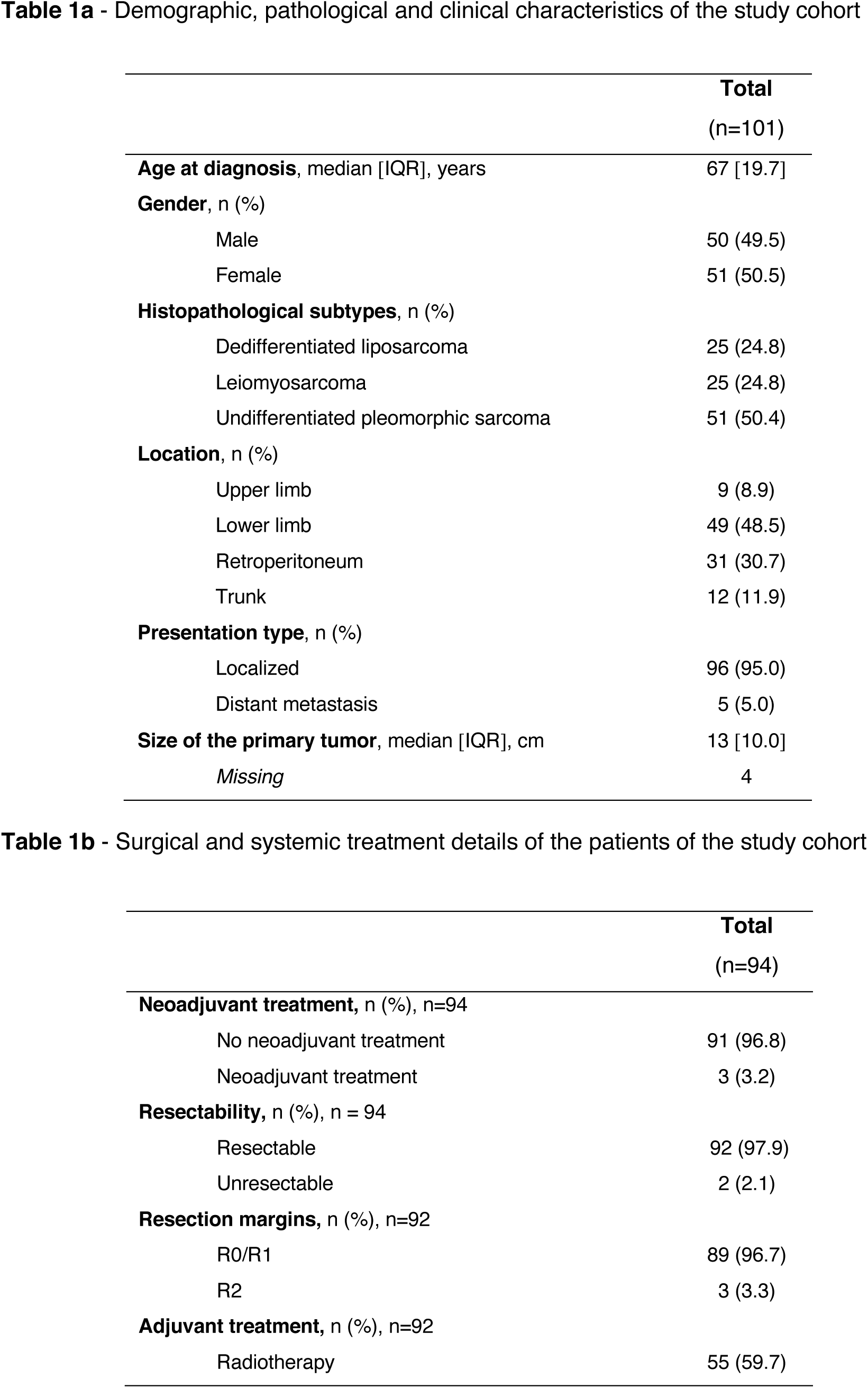
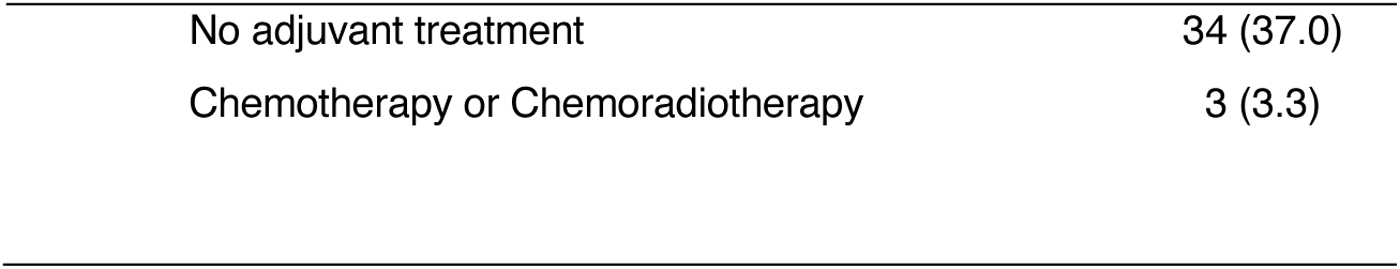
Demographic, pathological, clinical and treatment characteristics of the study cohort that was considered for the gene expression profiling evaluation.

### UPS displays distinct metabolic gene signatures

Differential gene expression analysis revealed that UPS display a closer gene expression pattern to high-grade LMS than to DDLPS (Supplementary Figure 1). The lower number of DEGs between UPS and high-grade LMS (67 DEGs; Supplementary Table 1) compared to UPS and DDLPS (241 DEGs; Supplementary Table 2), as well as the smaller magnitude of differential gene expression, with a mean log fold change (logFC) of 0.009 for UPS vs. high-grade LMS and 0.01 for UPS vs. DDLPS, support this finding.

To identify features specific to UPS, we examined DEGs and pathways, either globally and across metabolism-related genes. Although the F1RNA gene panel contained limited representation of metabolic genes (84 of 1519) (Supplementary Figure 2 and Supplementary Table 3), three TCA cycle and OXPHOS-related genes - *SDHB*, *SDHC*, and *SDHD* - and one arachidonic acid metabolism-related gene - *ALOX12* - were found to be significantly overexpressed in UPS relatively both to LMS and also to DDLPS (Supplementary Figures 3 and 4; Supplementary Table 4).

### *SDHB*, *SDHC* and *SDHD* are overexpressed in UPS relatively to high-grade LMS and DDLPS

Limma-Voom differential expression analysis confirmed that *SDHB*, *SDHC*, and *SDHD* were significantly overexpressed in UPS in comparison with both LMS and DDLPS, displaying positive logFC values and significant adjusted *p-values* (Figure 1). These genes encode subunits of the SDH complex, which bridges TCA cycle flux and electron transport chain (linking the TCA cycle and OXPHOS). These results suggest that mitochondrial activity is relatively upregulated in UPS.

**Figure 1.**
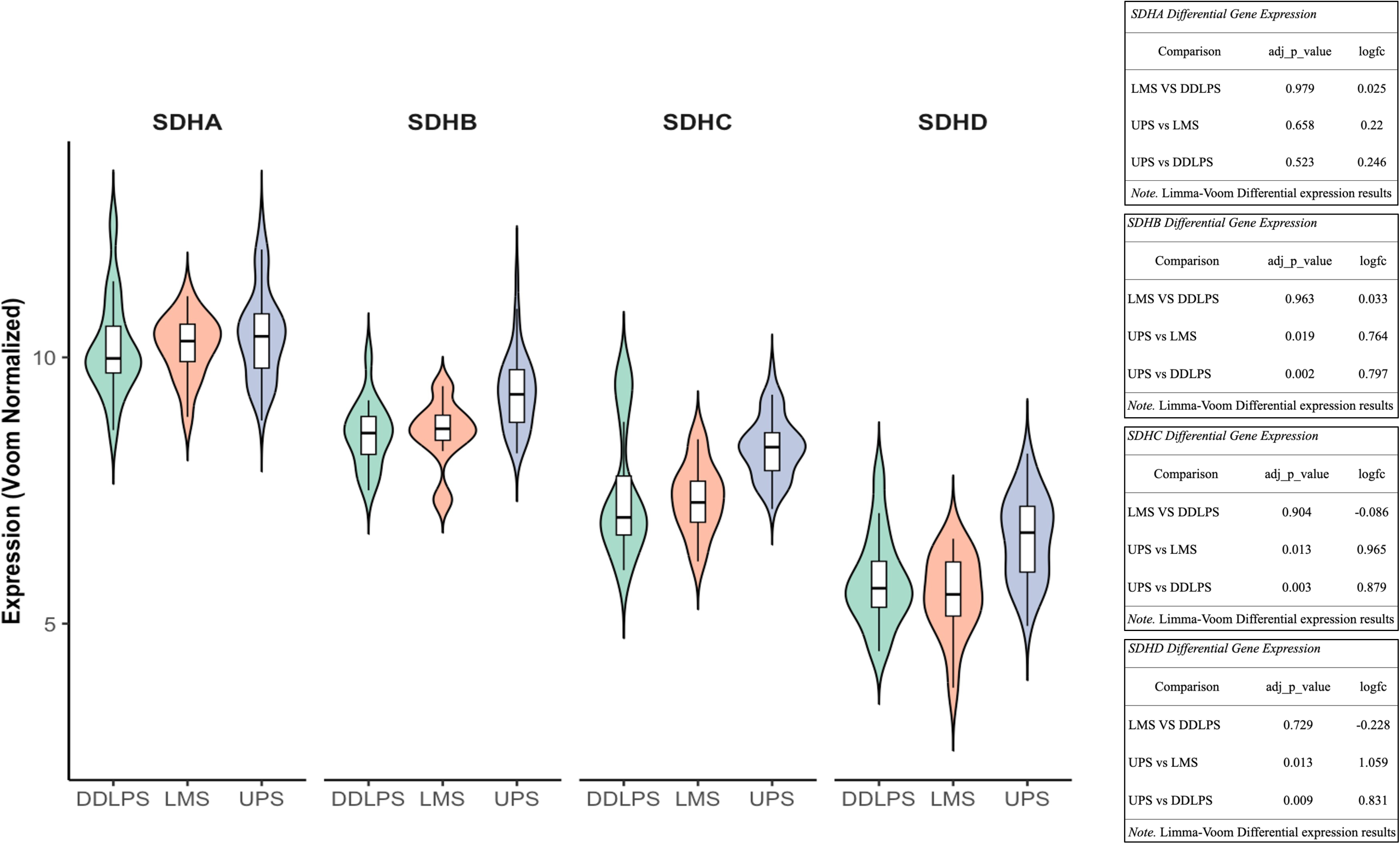
Limma-Voom differential expression of SDHA, SDHB, SDHC and SDHD for comparative evaluations between UPS, high-grade LMS, and DDLPS. For each SDH differential gene expression analysis (between different high-grade STS subtypes) adjusted p-values and logFC values are provided. SDHB, -C, and -D are all significantly overexpressed in UPS vs. high-grade LMS (adjusted p-value < 0.05) and in UPS vs. DDLPS (adjusted p-value < 0.05).

### *SDHB* overexpression is associated with poor survival in UPS

*SDH* genes overexpression is correlated with clinical outcomes in UPS. High *SDHB* expression was significantly correlated with shorter OS and shorter OS from the date of first metastasis in UPS, as shown by both Kaplan–Meier (Figures 2a and 2c) and Cox Proportional Hazards Models (Figures 2b and 2d). Moreover, for this specific UPS population, *SDHB* overexpression was not significantly associated with either MFS, RFS and PFS, though trends toward poorer outcomes were observed (Supplementary Figure 5). *SDHC* and *SDHD* overexpression showed no significant associations with neither OS nor other survival endpoints in UPS (Supplementary Figures 6 and 7). *SDHB*, *SDHC* and *SDHD* overexpression did not show any significant association with any survival endpoint in the whole cohort population (Supplementary Figures 8 to 11).

**Figure 2.**
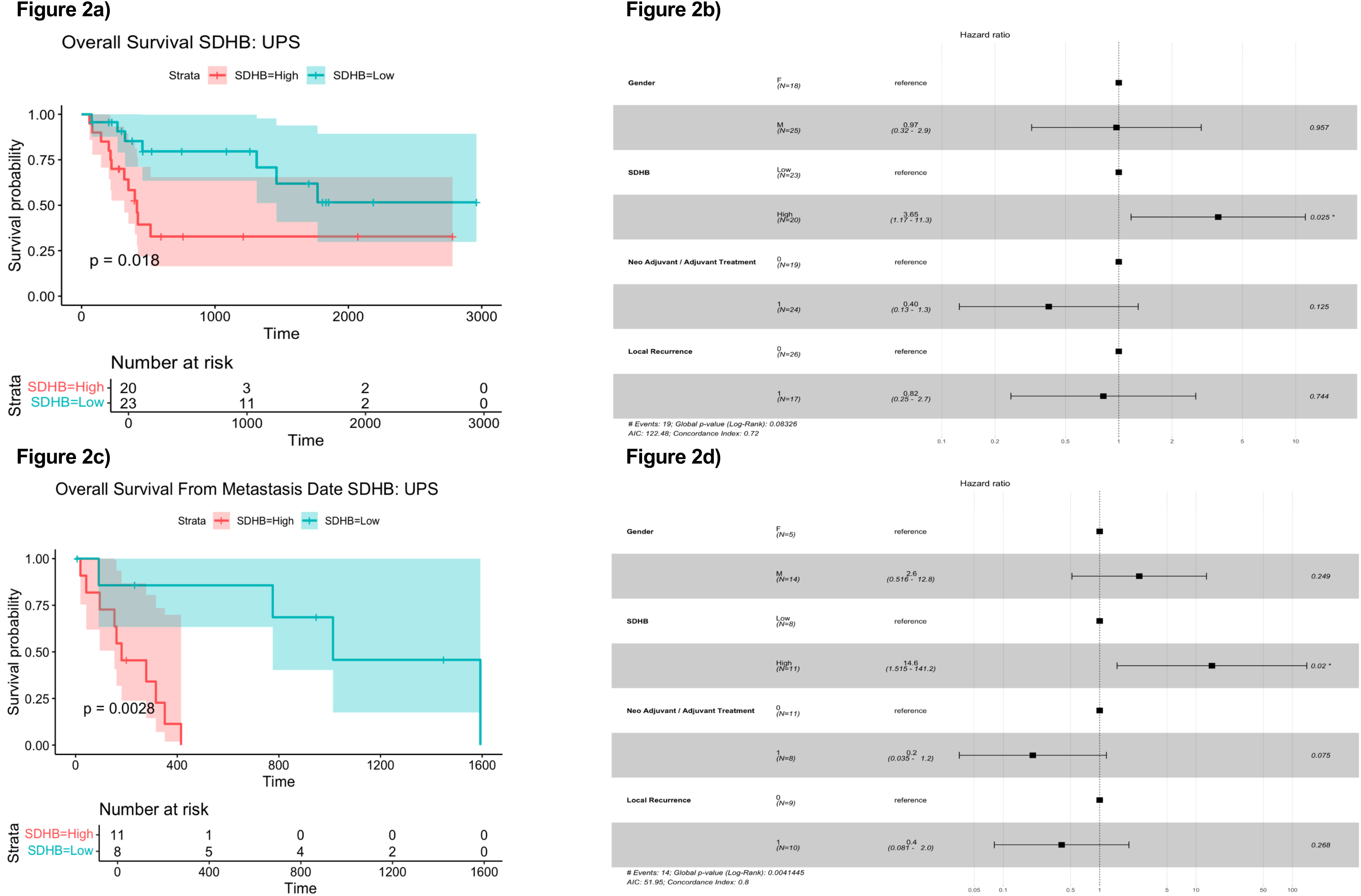
Within the UPS population, overexpression of SDHB is significantly correlated with shorter OS, either using a KMM (Figure 2a, p-value <0.05) and also using a CHPM (Figure 2b, p-value < 0.05), and also with a shorter OS from the date of first metastasis, either using a KMM (Figure 2c, p-value <0.05) and also using a CPHM (Figure 2d, p- value <0.05).

### TCGA-SARC validates UPS–LMS similarity and reveals UPS-specific pathway enrichment

In the independent TCGA-SARC cohort, UPS again demonstrated greater transcriptomic similarity to LMS than to DDLPS, with fewer DEGs and lower logFC between UPS and LMS (Supplementary Figure 12; Supplementary Tables 5 and 6). Gene set enrichment analysis (GSEA) and differential pathway expression analysis confirmed the significant upregulation of OXPHOS and pyrimidine metabolism in UPS compared with LMS (Supplementary Figure 13). Additional enrichment in immune activation-related pathways, such as antigen processing and presentation, and diverse DNA damage repair pathways/mechanisms was also observed in UPS. Pathway analysis reinforced the overexpression of diverse mitochondrial components and pathways, including SDH (complex II) and different respiratory chain and ATP synthesis pathways, in UPS relatively to LMS (Supplementary Figure 14).

### TCGA-SARC validates *SDHB* overexpression in UPS relatively to high-grade LMS and DDLPS

We evaluated whether the directionality of gene expression changes was conserved across both datasets. For each pairwise, we identified genes significantly differentially expressed in both cohorts and compared their logFC values to determine directional concordance (Supplementary Figures 15, 16 and 17). This directional agreement was statistically significant in all comparisons based on Chi-square tests (Supplementary Figure 18). Next, we performed similar differential gene expression analyses comparing UPS to high-grade LMS and to DDLPS samples from the TCGA-SARC cohort. In both comparisons, *SDHA* and *SDHB* were found to be differentially expressed. Notably, the differential expression of *SDHB* between UPS and both high-grade LMS and DDLPS was consistently observed in both the study cohort and the independent TCGA-SARC dataset. Limma-Voom differential expression analysis of *SDHA*, *SDHB*, *SDHC* and *SDHD* was performed for UPS vs. high-grade LMS vs. DDLPS (Figure 3). *SDHA* differential overexpression in UPS relatively to both high-grade LMS and DDLPS was found to be statistically significant. Similarly, *SDHB* differential overexpression in UPS relatively to both high-grade LMS and DDLPS was also found to be statistically significant. These results validate *SDHB* overexpression as a distinctive molecular feature of UPS.

**Figure 3.**
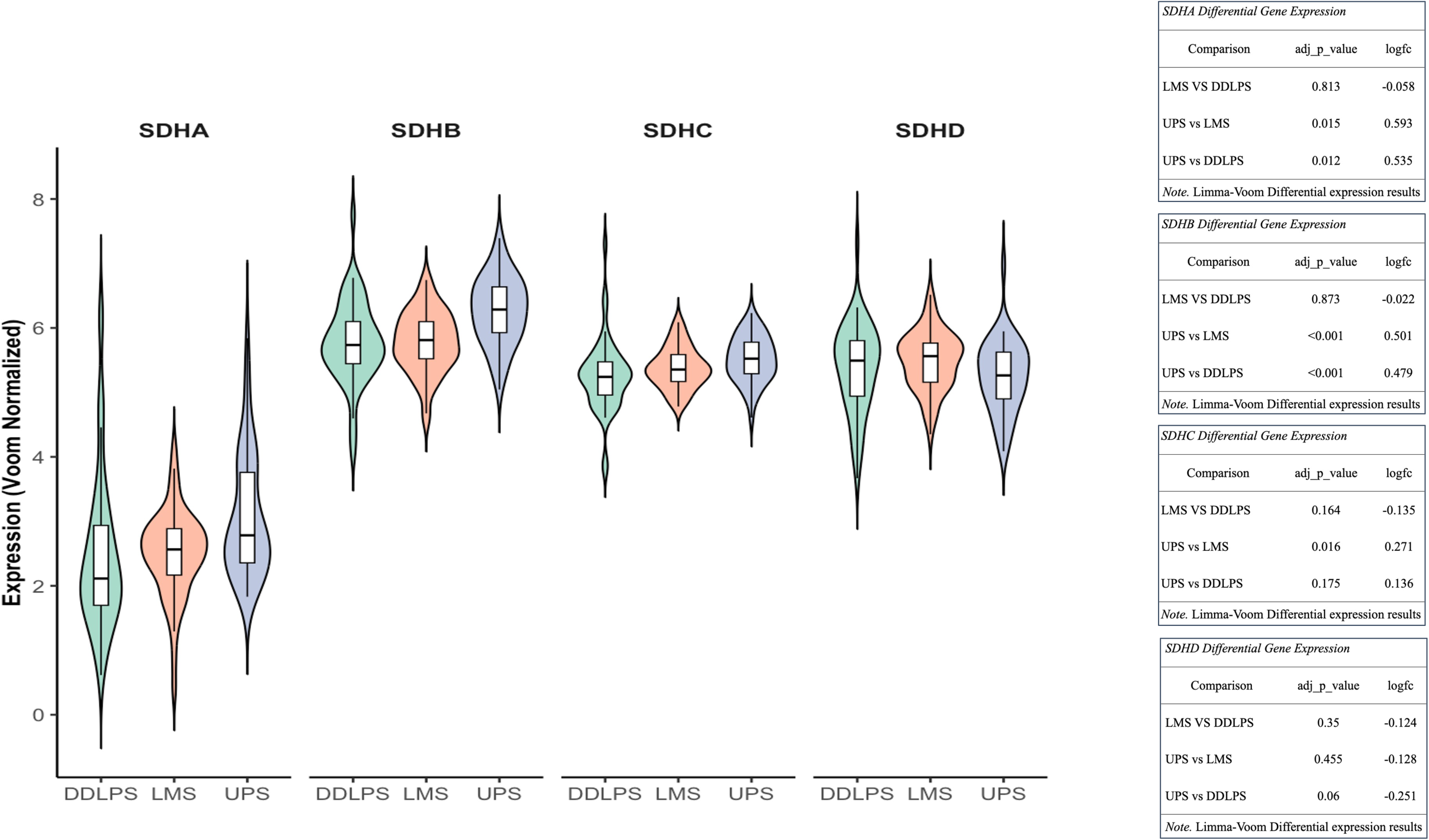
Limma-Voom differential expression of SDHA, SDHB, SDHC and SDHD for comparative evaluations between UPS, high-grade LMS, and DDLPS samples of an independent cohort (TCGA-SARC). For each SDH differential gene expression analysis (between different high-grade STS subtypes) adjusted p-values and logFC values are provided. SDHA and SDHB are significantly overexpressed in UPS vs. high-grade LMS (adjusted p-value < 0.05) and in UPS vs. DDLPS (adjusted p-value <0.05), while SDHC is only significantly overexpressed in UPS vs. high-grade LMS (adjusted p-value < 0.05).

### The associations of *SDHB* overexpression with survival endpoints in UPS could not be validated using TCGA-SARC

We attempted to validate the associations of *SDHB* overexpression with survival endpoints that have been observed in the study cohort in UPS. We used the TCGA-SARC cohort for this purpose.

We examined whether the direction of the association between gene expression and OS was consistent across both cohorts. For this purpose, we identified genes whose expression was significantly associated with OS in each cohort and determined whether their high expression predicted better or worse prognosis. The overlap of significant genes was then used to evaluate whether the direction of survival association was conserved. For the UPS-only analysis, the chi-squared test assessing survival direction concordance between both cohorts yielded a non-significant result (*p-value*=0.63) (Supplementary Figure 19). When the UPS, DDLPS and LMS samples were considered, the chi-squared test also yielded a non-significant result (*p-value*=0.71) (Supplementary Figure 20). This indicates that despite DGE concordance, survival associations do not seem to exhibit consistent directional agreement between cohorts.

We attempted to explore and examine potential causes of this discrepancy. An association between an enrichment in particular immune-related pathways (B cell and humoral-dependent immune pathways) and high expression levels of genes whose expression is correlated with improved prognosis within this universe of UPS samples was verified in the TCGA-SARC cohort (Supplementary Figure 21), but was not observed in our study cohort (Supplementary Figure 22), despite the coverage of the same immune-related pathways by the F1RNA gene set (Supplementary Figure 23). Interestingly, it is known that indels (generally) and that frameshift indels (more specifically) can serve as potent generators of highly immunogenic neoantigens, leading to increased immunogenicity and activation of humoral immune-related transcriptional programs [33]. In the study cohort, UPS did not exhibit either a higher general indel burden (Supplementary Figure 24), and did not show a higher specific frameshift indel burden (Supplementary Figure 25) compared to LMS or DDLPS. On the other hand, in the TCGA-SARC cohort, UPS samples displayed a markedly higher number of indels per patient relatively to the other two STS histotypes (Supplementary Figure 26), while no significant differences in terms of frameshift indels were found between UPS, LMS and DDLPS (even though UPS displayed a higher absolute number of frameshift indels when compared with LMS and DDLPS) (Supplementary Figure 27). It is important to note that within our study cohort we employed the F1CDx assay, which utilizes targeted sequencing, whereas TCGA-SARC relies on whole-exome sequencing (WES), which makes the absolute number and distribution of the detected indels non-directly comparable between cohorts. It is, indeed, possible that immunologically distinct features, likely driven by higher indel loads, influence survival patterns in the TCGA-SARC UPS cohort, limiting its utility for validating prognostic gene expression signatures identified in independent datasets. Moreover, the reduced size of the UPS samples pool (n=43 in the study cohort and n=44 in the TCGA-SARC cohort) of each cohort and differences in the composition of the gene sets employed for DNAseq in each cohort may also explain this discrepancy.

### SDHB protein is overexpressed in UPS and LMS relative to DDLPS

Immunohistochemical (IHC) staining of SDHB was performed on FFPE samples, and a novel three-tier scoring system was applied (Figure 4). UPS and LMS samples showed higher SDHB staining intensity than DDLPS. Specifically, 43% of UPS and 64% of LMS samples exhibited strong (score 3) staining, compared to 12% of DDLPS (Figure 5). Weak (score 1) staining was observed in 42% of DDLPS but was rare in UPS (6%) and absent in LMS. These findings confirm SDHB overexpression at the protein level in UPS and LMS.

**Figure 4.**
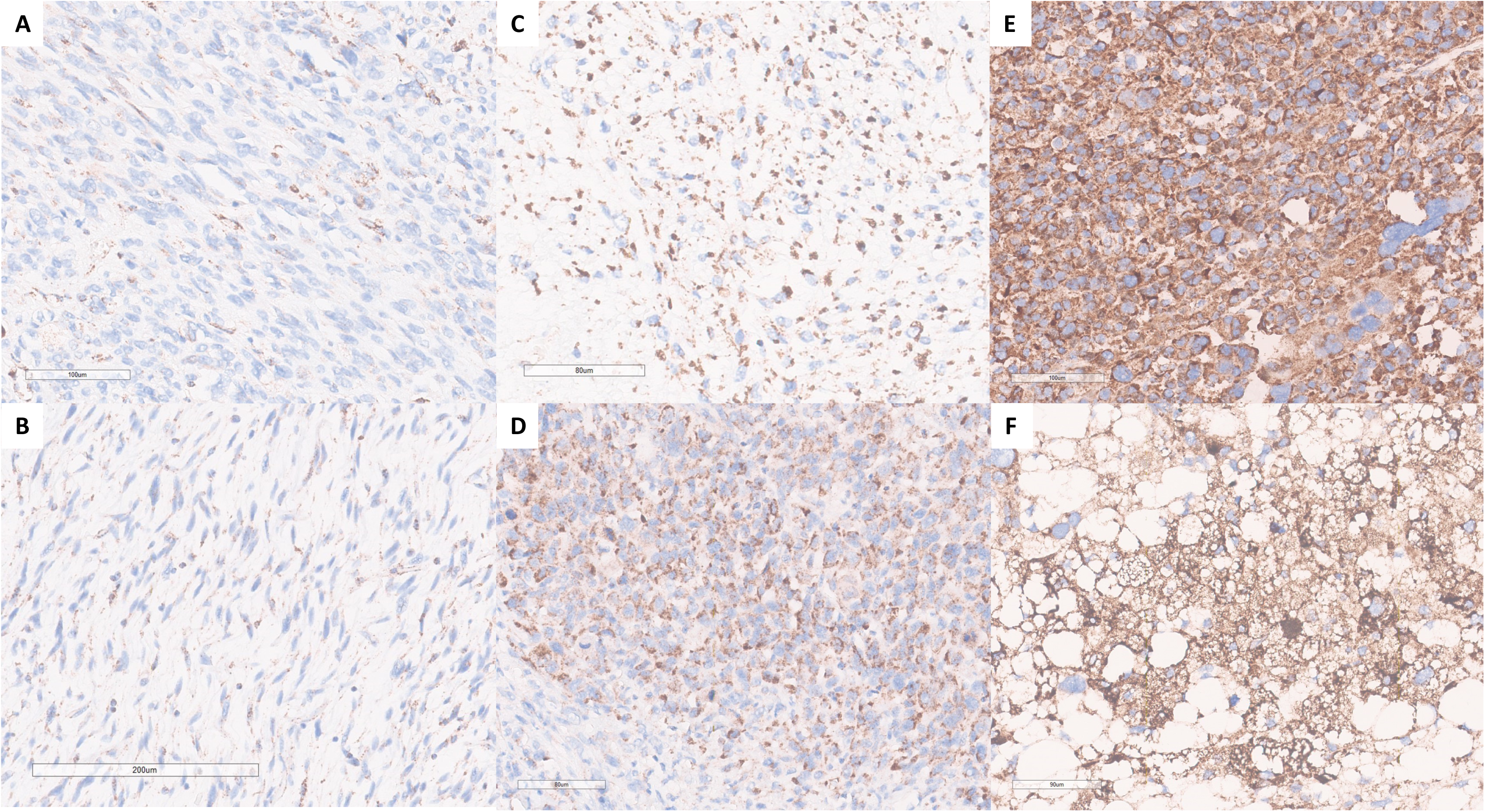
SDHB immunohistochemistry in sarcomas: Three-tier score for SDH immunostaining intensity - A and B with weak and focal granular cytoplasmic immunostaining (score 1); C and D with diffuse intermediate granular cytoplasmic immunostaining (score 2); E and F with diffuse and strong granular cytoplasmic immunostaining (score 3).

**Figure 5.**
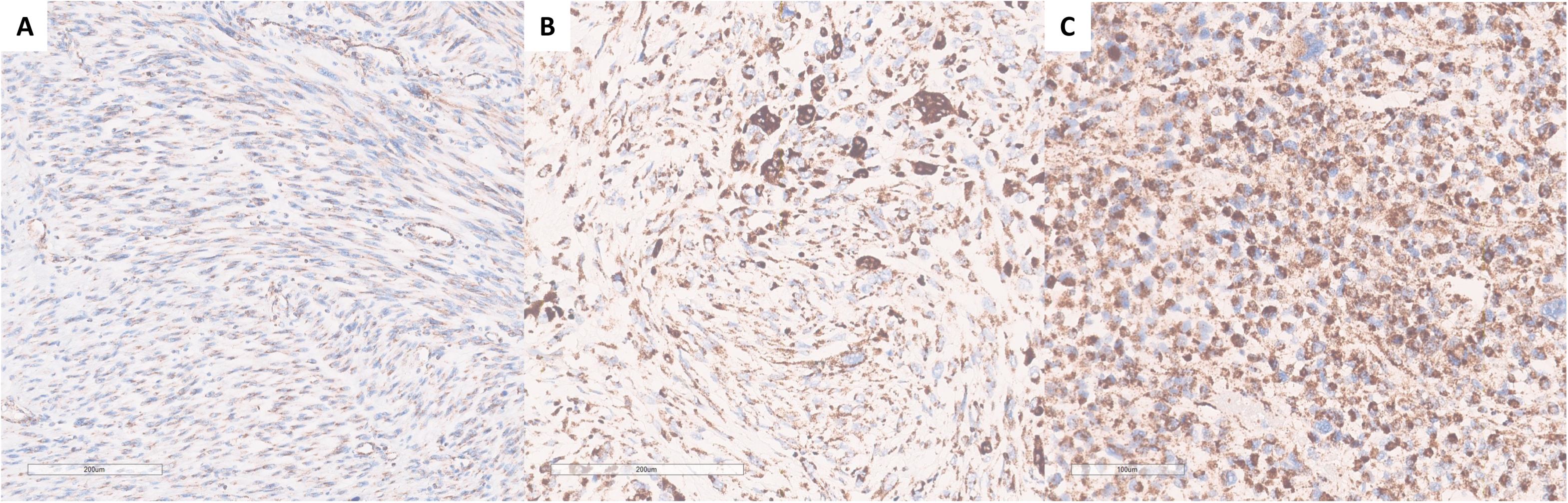
SDHB immunohistochemistry in sarcomas: Differences in immunostaining intensity related to tumoral heterogeneity and cell morphology - Spindle cells with scarce and ill-defined cytoplasm tend to show a more focal and weak staining (A); epithelioid and pleomorphic cells, as well as multinucleated giant cells, with a more voluminous cytoplasm tend to show more granularity and diffusely intense staining (B and C).

### Metabolomic profiling reveals distinct sarcoma subtype–specific metabolic signatures

To assess whether these molecular differences translated to metabolic phenotypes, we performed ^1^H NMR metabolomics on fresh tumor tissue from UPS (n=3), LMS (n=3), and LPS (n=2) patients. Detailed demographic, pathological, surgical and systemic treatment specificities of the 16 patients from whom normal and tumor tissue samples were collected are provided in Supplementary Tables 7 and 8).

Forty metabolites were identified, with ten differing significantly between subtypes (Figure 6a). UPS displayed higher levels of branched-chain amino acids (leucine, isoleucine, valine), phenylalanine, proline, carnosine, and uracil relative to LMS, consistent with increased protein turnover, anabolic growth, and oxidative stress adaptations. In contrast, LMS showed higher myo-inositol, NAD⁺, and adenine nucleotides, reflecting smooth muscle–specific metabolic programs. Although absolute succinate and fumarate levels did not differ significantly, the succinate-to-fumarate ratio was significantly higher in UPS compared with both LMS and LPS (Figure 6b), demonstrating impaired SDH enzymatic activity despite overexpression of SDH subunits.

**Figure 6a).**
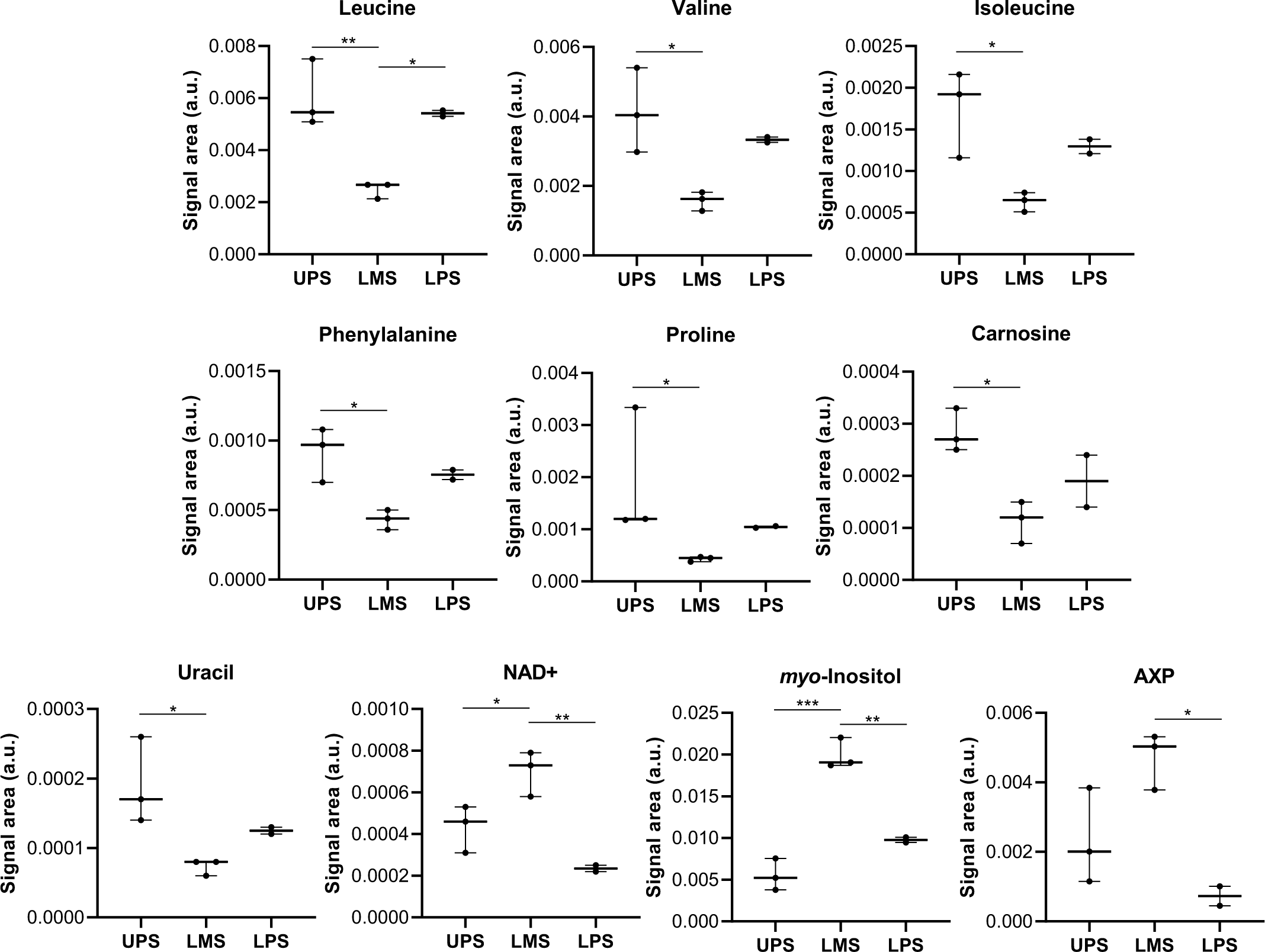
Metabolites whose abundance was significantly different between at least two of the three STS subtypes. *: p-value <0.05; **:p-value <0.01.

**Figure 6b).**
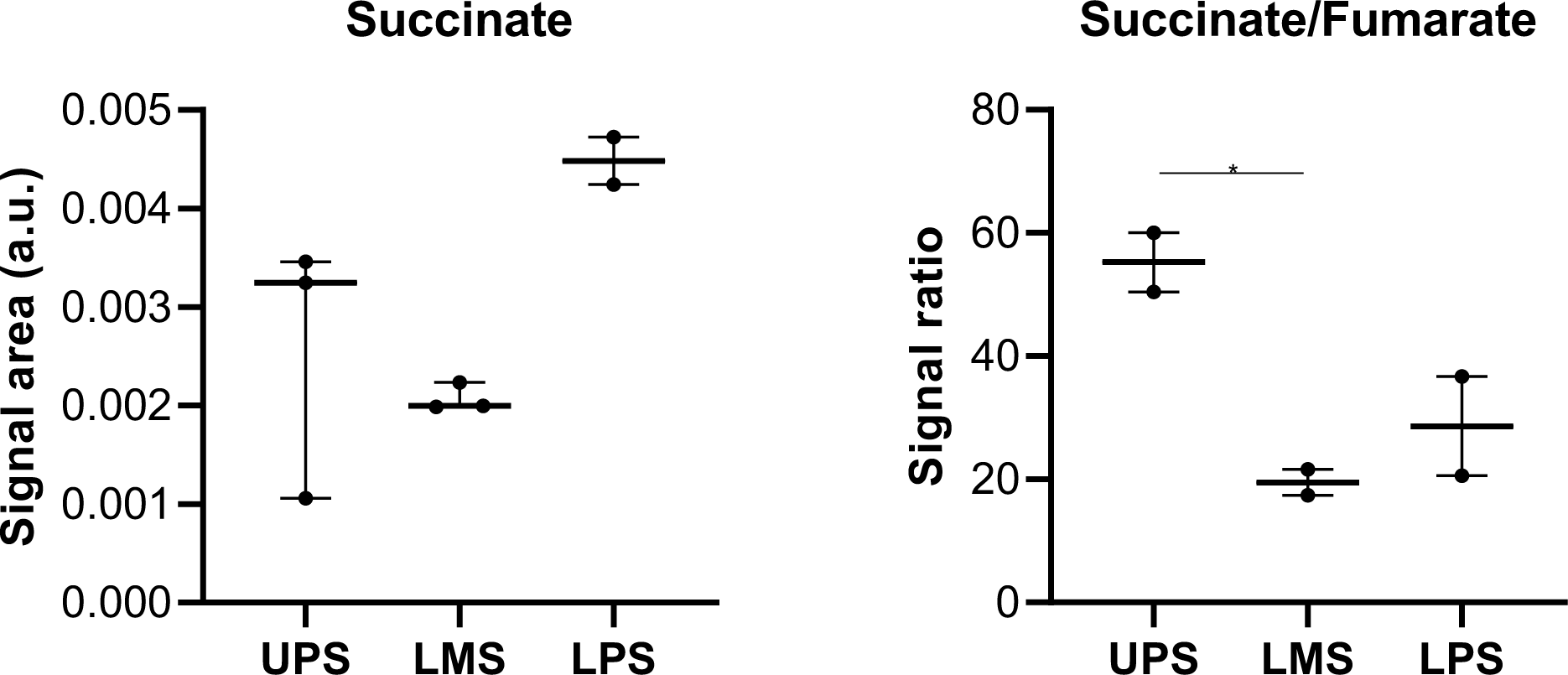
Succinate levels and succinate-to-fumarate ratio in the different STS subtypes. Succinate-to-fumarate ratio is significantly higher in UPS vs. LMS and vs. LPS.

## Discussion

The identification of molecular and biological traits that distinguish UPS from high-grade LMS and DDLPS has not been deeply explored. Isolated single-omics studies, with limited sample sizes, compared UPS and LMS and highlighted a handful of differentially expressed microRNAs that target either UPS and LMS-related genes [12] and SRC as potential discriminators [13], but these small studies did not robustly separate these two histopathological subtypes at the transcriptomic level. Our study is the first to put the scope on a direct comparative analysis of the molecular profiles of UPS, LMS and DDLPS employing a multi-omics approach.

The gene expression profiling of the high-grade STS samples included in the study cohort points towards a closer molecular similarity between UPS and high-grade LMS, a finding that was validated in the TCGA-SARC dataset and that is consistent with prior descriptions [34].

UPS are distinctively characterized by the overexpression of TCA cycle and OXPHOS-related genes, namely *SDHB*, *SDHC* and *SDHD*, a finding that was also validated in the TCGA-SARC dataset. While increased mitochondrial activity and TCA cycle upregulation have been previously reported in UPS [35,36], the overexpression of SDH subunit– encoding genes represents a novel observation.

SDH, positioned at the interface of the TCA cycle and the electron transport chain (ETC), is typically portrayed by a panoply of loss-of-function mutations in sarcomas which promote succinate accumulation, epigenetic dysregulation, and oncogenesis [37, 38, 39]. Overexpression of SDH subunits, in contrast, has been rarely described in cancer, though elevated *SDHA* expression levels have been observed in metastatic uveal melanoma [39], advanced ovarian cancer [40], and multiple myeloma [41], where they are linked to enhanced OXPHOS capacity, increased expression of proteins that shuttle electrons between different complexes of the ETC, augmented levels of key TCA enzymes, enhanced ATP yield, and amplified metabolic plasticity [39,40]. While *SDHA* encodes a flavoprotein, *SDHB* encodes the iron-sulfur protein that functions as the SDH catalytic core [40]. Given its role as the catalytic core of the SDH complex, *SDHB* overexpression may be linked to its dysfunction in UPS.

The correlation between high *SDHB* expression and poor survival in UPS in the study cohort further underscores the clinical relevance of this metabolic phenotype. Survival associations could not be validated using TCGA-SARC, possibly due to either the small size of the UPS samples pool of each cohort and different coverage of the gene sets employed for DNAseq in each cohort, or potentially to the putative survival impact of distinct immunological profiles of UPS populations of each cohort, likely driven by higher relative indel burden and overexpression of antigen processing and presentation pathways of UPS samples of the TCGA-SARC dataset.

IHC evaluation confirmed higher SDHB protein abundance in UPS and LMS compared to DDLPS. The proposed three-tier scoring system for SDHB staining provides a newly developed tool to categorize SDH expression levels in sarcoma tissues, complementing transcriptomic findings and offering a potential biomarker for future studies. Nevertheless, it is important to note that intra-tumoral heterogeneity and cell morphological patterns may influence the evaluation of granular staining intensity, potentially without any correlation with underlying genomic and/or transcriptomic alterations. This is particularly relevant for UPS, which may show areas with different morphologies mixing epithelioid, rhabdoid, pleomorphic and spindle cells. SDH granules are in the cytoplasm. Epithelioid, rhabdoid and pleomorphic cells tend to have a more voluminous cytoplasm and, therefore, effectively more granules that are more easily detectable. Hence, staining tends to appear more intense and diffused in regions rich in these cells. Spindle cells, on the other hand, have a scarce and ill-defined cytoplasm, typically showing fewer granularities. This way, staining tends to appear less intense in regions rich in spindle cells.

Metabolomic profiling further revealed distinctive metabolic features of UPS, including an enrichment in amino acids, uracil, and carnosine. Increased amino acid levels may reflect enhanced protein turnover, anabolic growth, or altered transamination, consistent with aggressive tumor behavior [42]. Higher uracil levels suggest augmented nucleotide turnover or dysregulated pyrimidine metabolism [43], in line with enhanced metabolic plasticity and proliferative signaling, while increased levels of carnosine, a dipeptide with antioxidant and pH-buffering properties [44], may indicate oxidative and metabolic stress adaptation. Carnosine accumulation could also contribute to immune evasion by modulating the expression of immune-modulatory proteins, protecting the neoplastic tissue from T cell-mediated immune surveillance [45]. Together, these findings seem to locate critical metabolic adaptations (with clinical impact) of aggressive and progressing UPS in the mitochondria, potentially at the ETC.

The higher succinate-to-fumarate ratio observed in UPS, despite SDHB overexpression, suggests functional impairment of SDH enzymatic activity, potentially due to post-translational modifications, SDH subunits misassemble, or TCA cycle flux imbalance. Future work in this direction will identify the post-translational modifications that could be responsible for this observation and the implications of this phenotype on UPS microenvironmental composition and on the nature of antitumor (UPS) immune response.

Indeed, succinate accumulation was shown to increase antigen presentation by the transcriptional and epigenetic activation of MHC-APP-related genes in melanoma, potentially enhancing tumor immunogenicity and indirectly modifying the microenvironmental immune composition [46]. The clarification of the interplay between SDH dysfunction, succinate accumulation and immunogenicity specifically in UPS is thus crucial, since the microenvironmental immune composition of UPS was shown to have prognostic value (denser infiltration by CD8+T lymphocytes and monocytes is associated with significantly better survival outcomes [47,48]), and the intracellular succinate levels of both tumor cells and microenvironmental CD8+ T cells are known to be linked to different profiles of anti-tumor immunogenicity [49].

This study identified a paradoxical phenotype of SDH subunit overexpression with functional impairment in UPS, defining a molecular and metabolic subtype associated with poor prognosis. These findings suggest novel avenues for understanding UPS pathogenesis and for developing therapies targeting mitochondrial metabolism and its interaction with the immune microenvironment. Future studies should clarify the mechanisms underlying SDH dysfunction in UPS and explore the therapeutic potential of modulating SDH activity and succinate levels to improve clinical outcomes.

## Data Availability

All data produced in the present study are available upon reasonable request to the authors.

## Conflicts of Interest Statement

**Miguel Esperança-Martins (MEM)**

<u>Research Grants</u>: Hoffmann-LaRoche, Foundation Medicine Inc., PharmaMar;

<u>Invited Speaker:</u> Bayer;

<u>Advisory Boards:</u> Gilead.

**Richard S.P. Huang (RH)**

Richard Huang is an employee of Foundation Medicine Inc, a wholly owned subsidiary of Roche and receives stocks from Roche

**Isabel Fernandes (IF)**

<u>Research Grants:</u> MSD; PharmaMar.

**Brian Van Tine (BVT)**

Brian A. Van Tine reports research grants from Polaris Pharmaceuticals and personal fees from consulting from Aadi Bioscience, Actua Capital Partners LLC, Advenchen Laboratories, Boxer Capital LLC, Crisper Therapeutics, Deciphera Pharmaceuticals Inc., Daiichi Sankyo Company, EcoR1 Capital LLC, Galapagos, Hinge Bio, Putnam, Sonata Therapeutics, and Salarius Pharmaceuticals Inc.; Honoraria for educational talks from Beijing Biostar Pharmaceuticals, Co., LTD., Oncology Education, and Total Health Conference; participation on a data/safety advisory board for Daiichi Sankyo, Bayer US Medical Affairs, PTC Therapeutics, Aadi Biosciences, Boehringer Ingelheim, Regeneron Pharmaceuticals, Curis, Syneos Health, Deciphera Pharmaceuticals, and Curtis; nonpaid service as a board member with Polaris; travel support from Adaptimmune LLC; licensing/patents with Accuronix Therapeutics.

**Luis Costa (LC)**

<u>Research Grants:</u> MSD; Eli Lilly; Amgen, Roche, Janssen;

<u>Invited Speaker:</u> Hoffmann-LaRoche, Gilead; AstraZeneca; Eli Lilly; MSD; BMS, Astellas.

<u>Advisory Boards:</u> Roche; AstraZeneca; Bayer; Pfizer; Gilead, Novartis; Servier.

## Declarations

### Ethics Approval and Consent to Participate

The present study was performed in accordance with the ethical standards of Helsinki Declaration II and was approved by the Institution Review Boards of both CAML and IPOLFG.

### Consent for Publication

Not Applicable.

### Authors’ Contributions

#### Authors Initials

Miguel Esperança-Martins (MEM) (First and Corresponding Author)

Hugo Vasques (HV)

Manuel Sokolov Ravasqueira (MSR)

Filipa Santos (FS)

Filipa Fonseca (FF)

António Syder Queiroz (ASQ)

João Boavida (JB)

Daniel Martins Jordão (DMJ)

Joaquim Soares do Brito (JSB)

Patrícia Corredeira (PC)

Marta Martins (MM)

Ângela Afonso (AA)

Jorge Antonio López (JAL)

Richard S.P.Huang (RH)

Cecília Melo-Alvim (CMA)

Isabel Fernandes (IF)

Dolores López-Presa (DLP)

Maria Manuel Lemos (MML)

Brian Van Tine (BVT)

Alliny Bastos (AB)

Sandra Casimiro (SC)

Nuno Abecasis (NA)

Luís Costa (LC)

Emanuel Gonçalves (EG)

Iola Duarte (ID)

Sérgio Dias (SD)

#### Contributions

<u>Contribution of each of the authors</u>

Conceptualization: MEM, HV, MSR, FS, IF, MML, BVT, AB, LC, EG, ID, SD.

Methodology: MEM, HV, MSR, FS, MML, SC, EG, ID, SD.

Investigation: MEM, HV, MSR, FS, FF, ASQ, JB, DMJ, JSB, PC, MM, AA, CMA, DLP, MML, SC, EG, ID, SD.

Visualization: MEM, MSR, FS, FF, PC, MML, SC, EG, ID, SD.

Funding acquisition: MEM, HV, IF.

Project administration: MEM, HV, IF, SD.

Supervision: HV, JAL, RH, IF, MML, BVT, AB, SC, NA, LC, EG, ID, SD.

Writing – original draft: MEM, HV, MSR, FS, FF, ASQ, MML, SC, EG, ID, SD.

Writing – review & editing: MEM, MSR.

## Acknowledgements

This research was sponsored by F.Hoffmann-LaRoche AG and by Foundation Medicine Inc under the RNA LDT Research Programme and also by PharmaMar. We would like to thank F.Hoffmann-LaRoche AG and Foundation Medicine Inc for providing the FoundationOne®CDx and FoundationOne®RNA assays, and for all the technical support, specifically during the transfer of the sequencing data via the safe platform. We would also like to thank PharmaMar for sponsoring the acquisition of laboratory reagents and materials.

We would like to thank Sarah Yacoub (Foundation Medicine Inc) for all the precious help in the logistical operationalization of the transfer of all of the different batches of samples between iMM (Lisbon, Lisbon, Portugal) and the Foundation Medicine Headquarters (Cambridge, Massachusetts, United States of America) and for her valuable assistance on sending the results of both DNA and RNAseq results via a safe platform.

We would also like to thank Rachel Beth Keller-Evans (Foundation Medicine Inc) for her crucial support in the analysis of the data that has resulted from the employment of the FoundationOne®RNA assay, namely for her help in the analysis of the detected fusions. We would also like to show our deepest gratitude to the IPOLFG tumor biobank staff (that were responsible for the original retrieval and organization both of the FFPE samples and also of some of the fresh tissue samples that were used in this study), the IPOLFG pathology department staff (who developed, in collaboration with the IPOLFG pathologists, the protocol for SDHB immunostaining, who assisted both the two pathologists that scored SDHB immunoreactivity and also the pathologist that reviewed each FFPE sample), the iMM biobank staff (that were responsible for the collection, storage and organization of the majority of the fresh tissue samples that were used in this study, and that were also responsible for the preparation and shipment of fresh tissue samples to CICECO (Aveiro, Portugal) for the ^1^H NMR evaluation), the iMM Comparative Pathology unit team (that have sectioned the FFPE blocks), the iMM Translational Oncobiology Lab staff (that have helped in the preparation and shipment of the different batches of samples and that have also played a key role in the development of the protocol for SDHB immunostaining), the iMM Technology Transfer Office staff, and the CICECO staff (who have played a crucial role in the logistical operationalization of the ^1^H NMR evaluation).

We would finally like to thank Li Cao for designing the graphical abstract.

