## Supplementary Figures for "Succinate dehydrogenase B (SDHB) overexpression with enzymatic dysfunction defines a distinct subtype of undifferentiated pleomorphic sarcoma"

### Slide 1
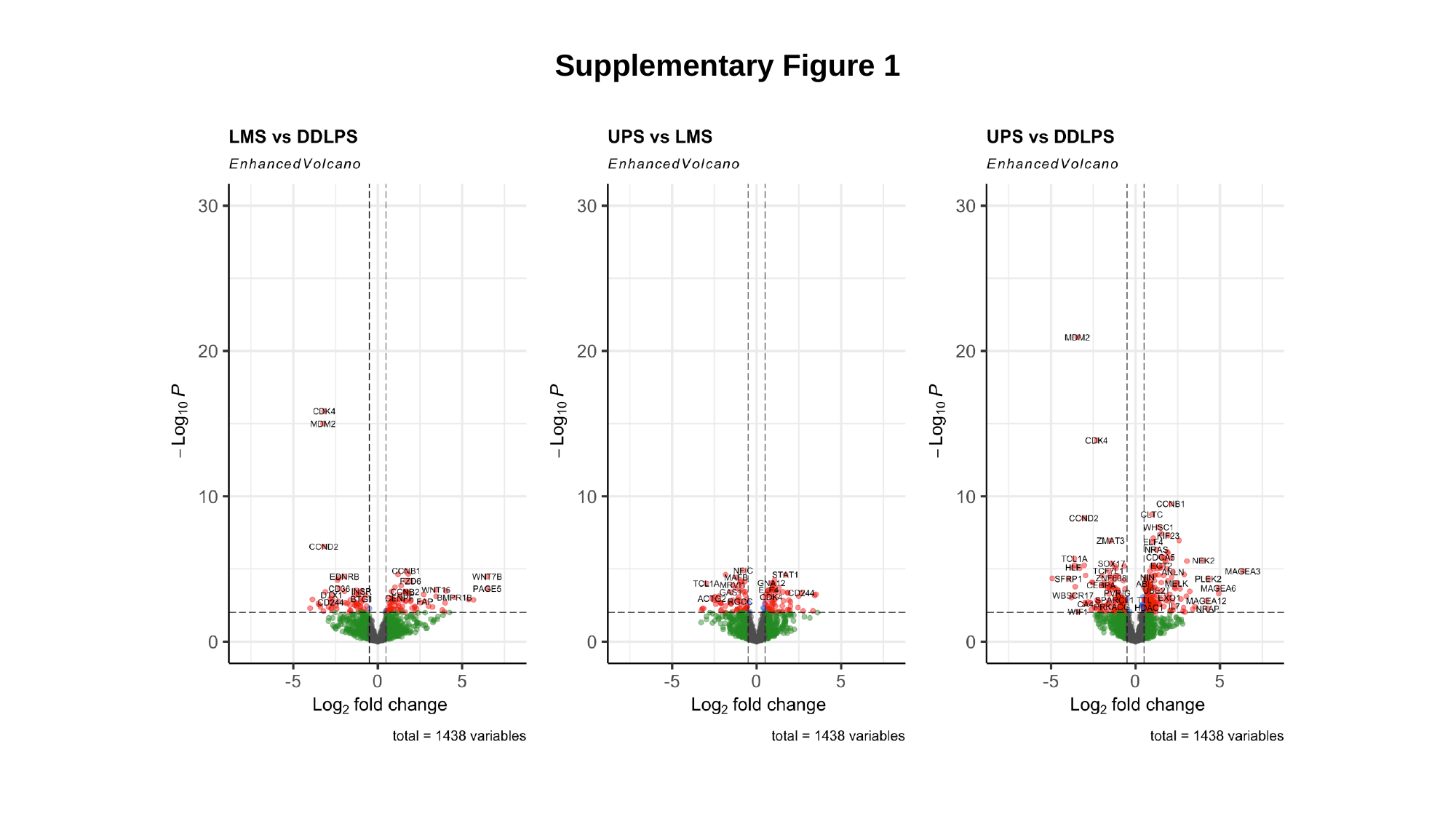

Supplementary Figure 1

### Slide 2
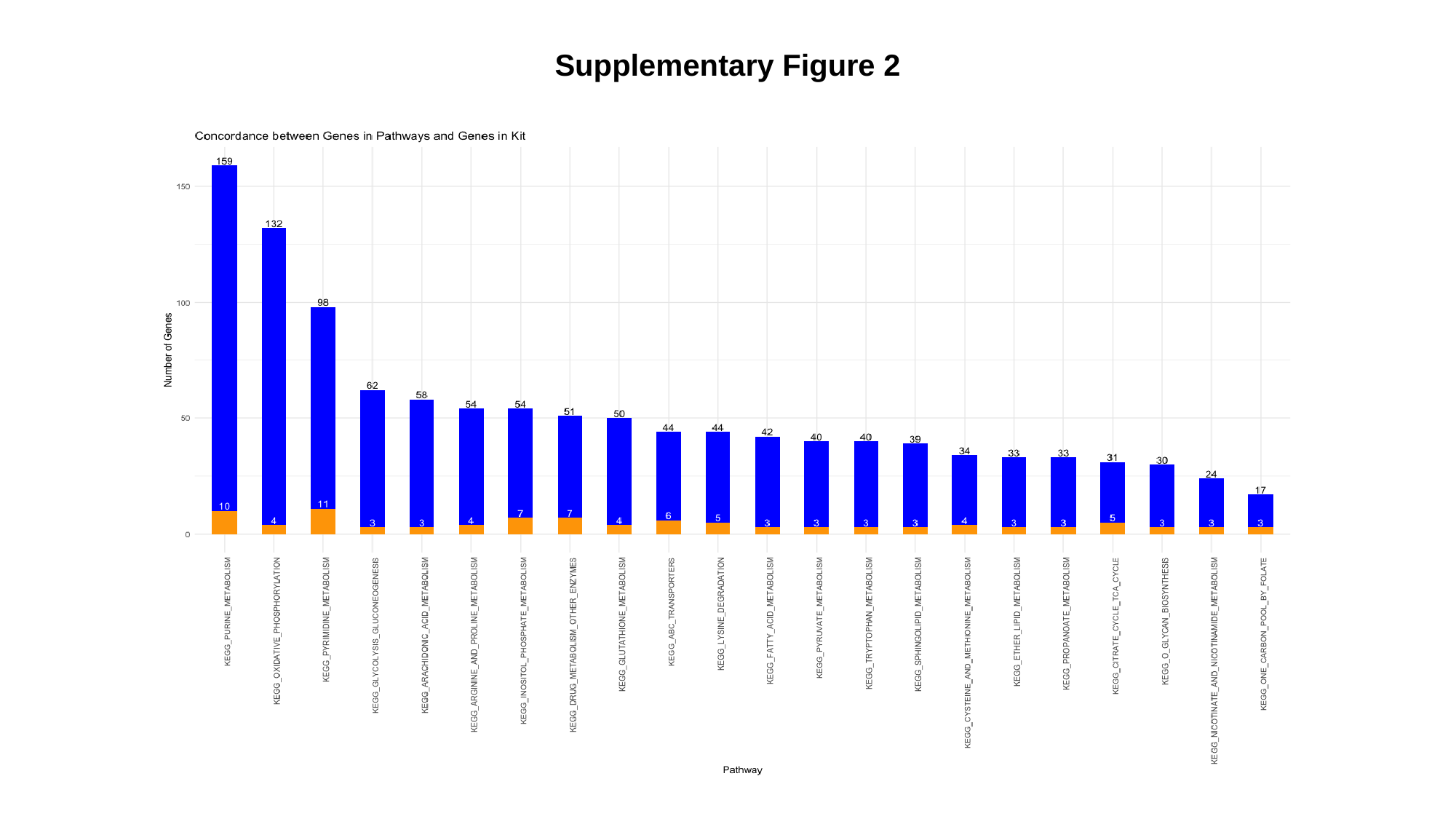

Supplementary Figure 2

### Slide 3
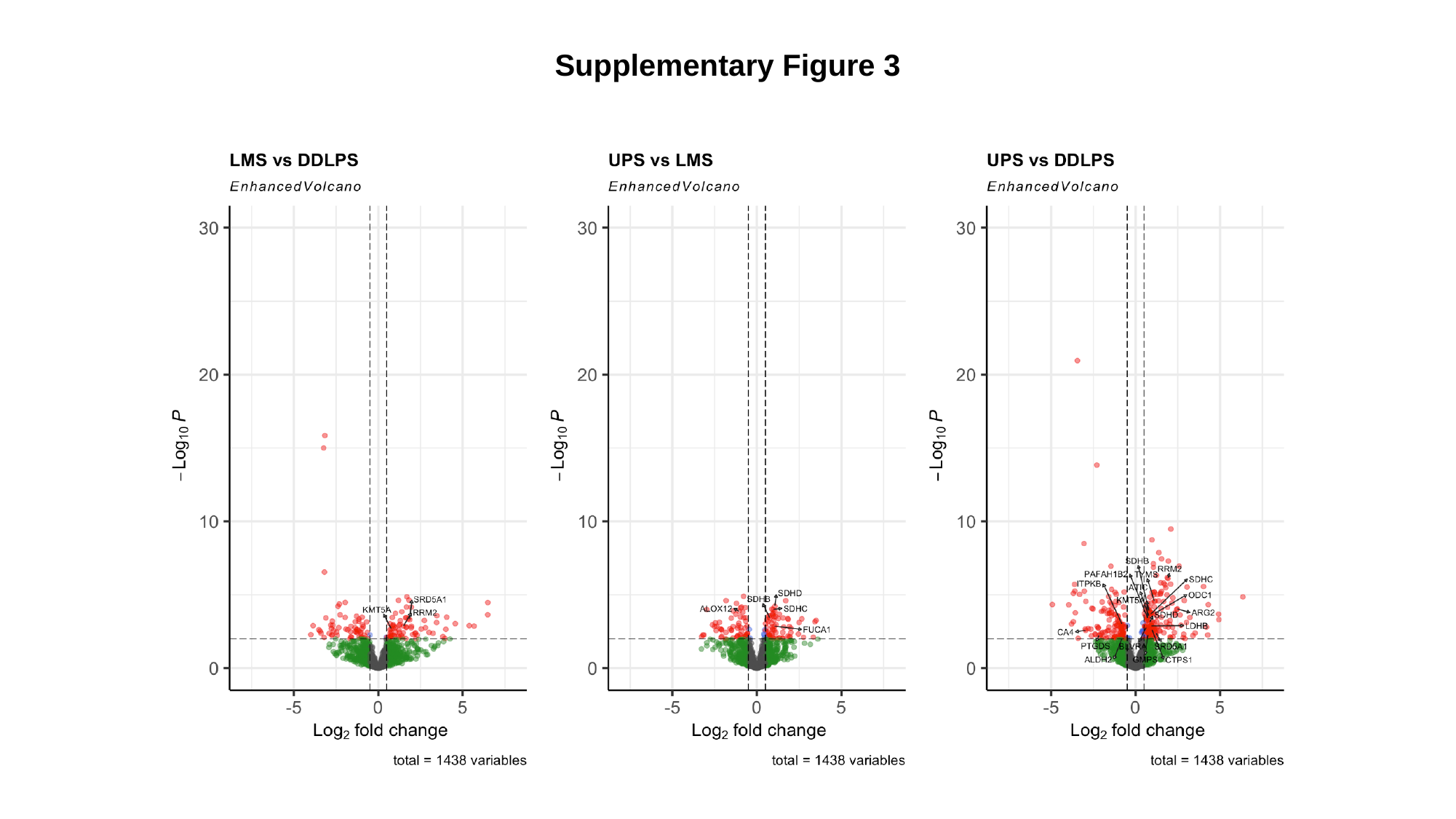

Supplementary Figure 3

### Slide 4
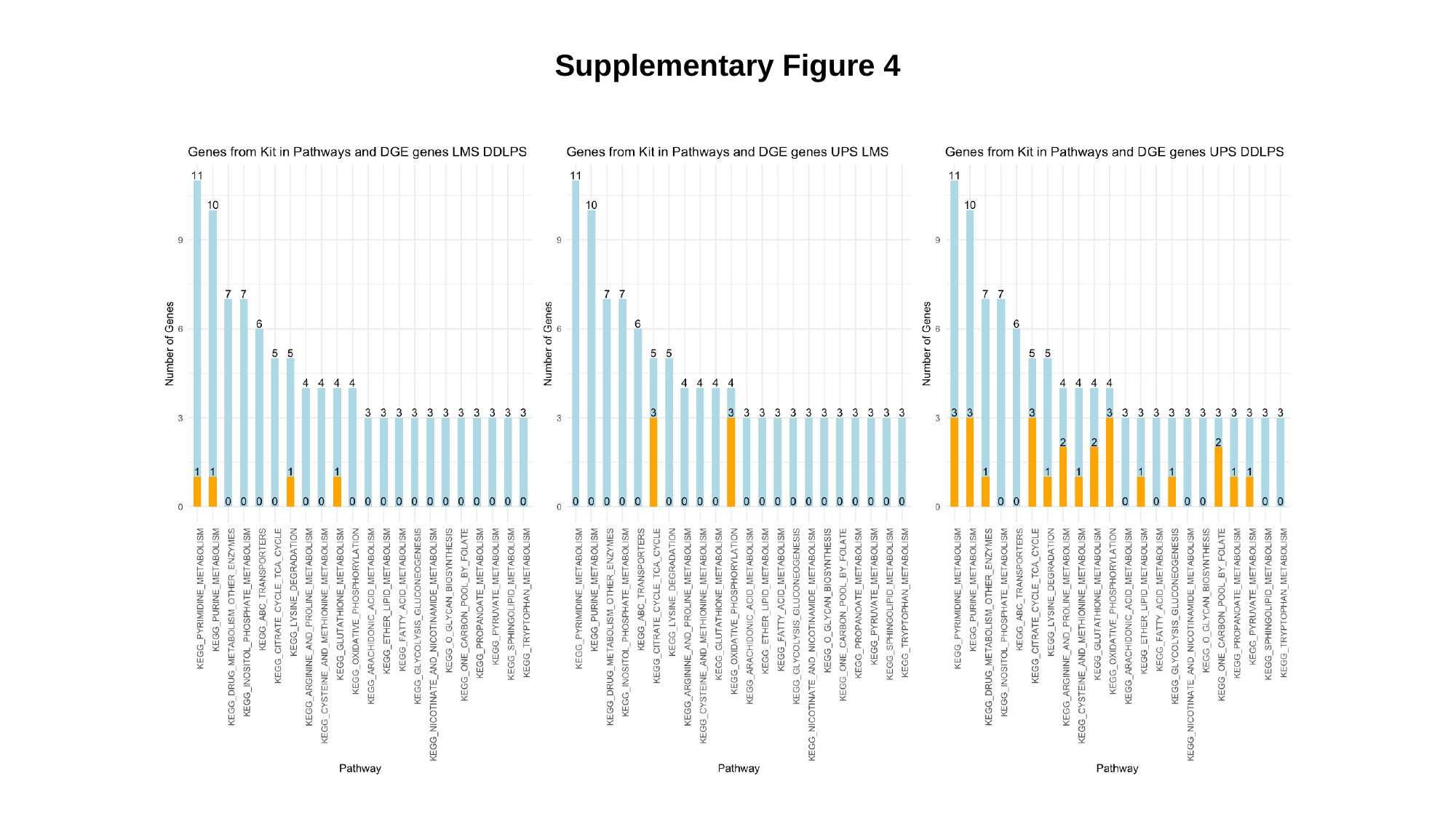

Supplementary Figure 4

### Slide 5
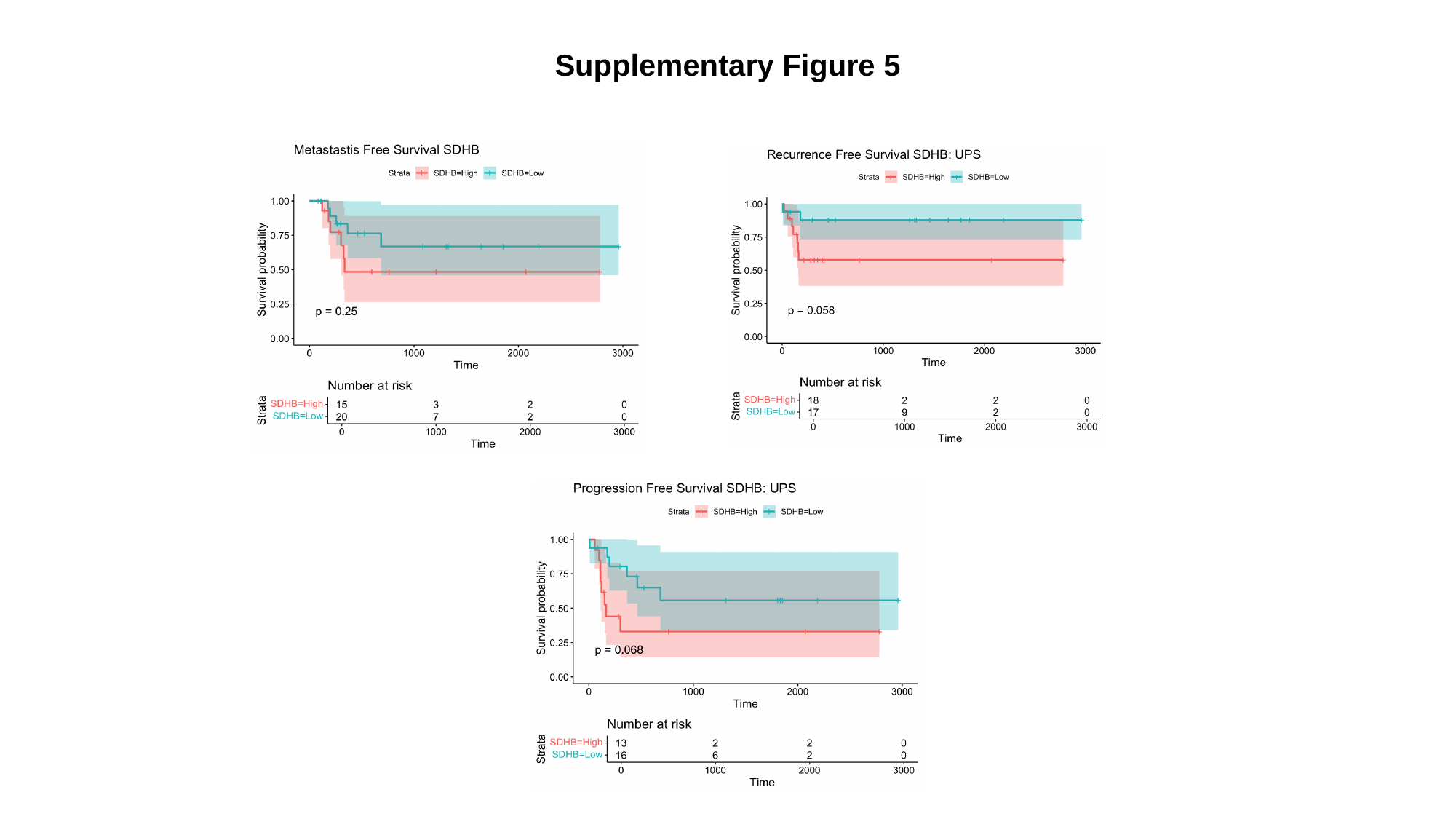

Supplementary Figure 5

### Slide 6
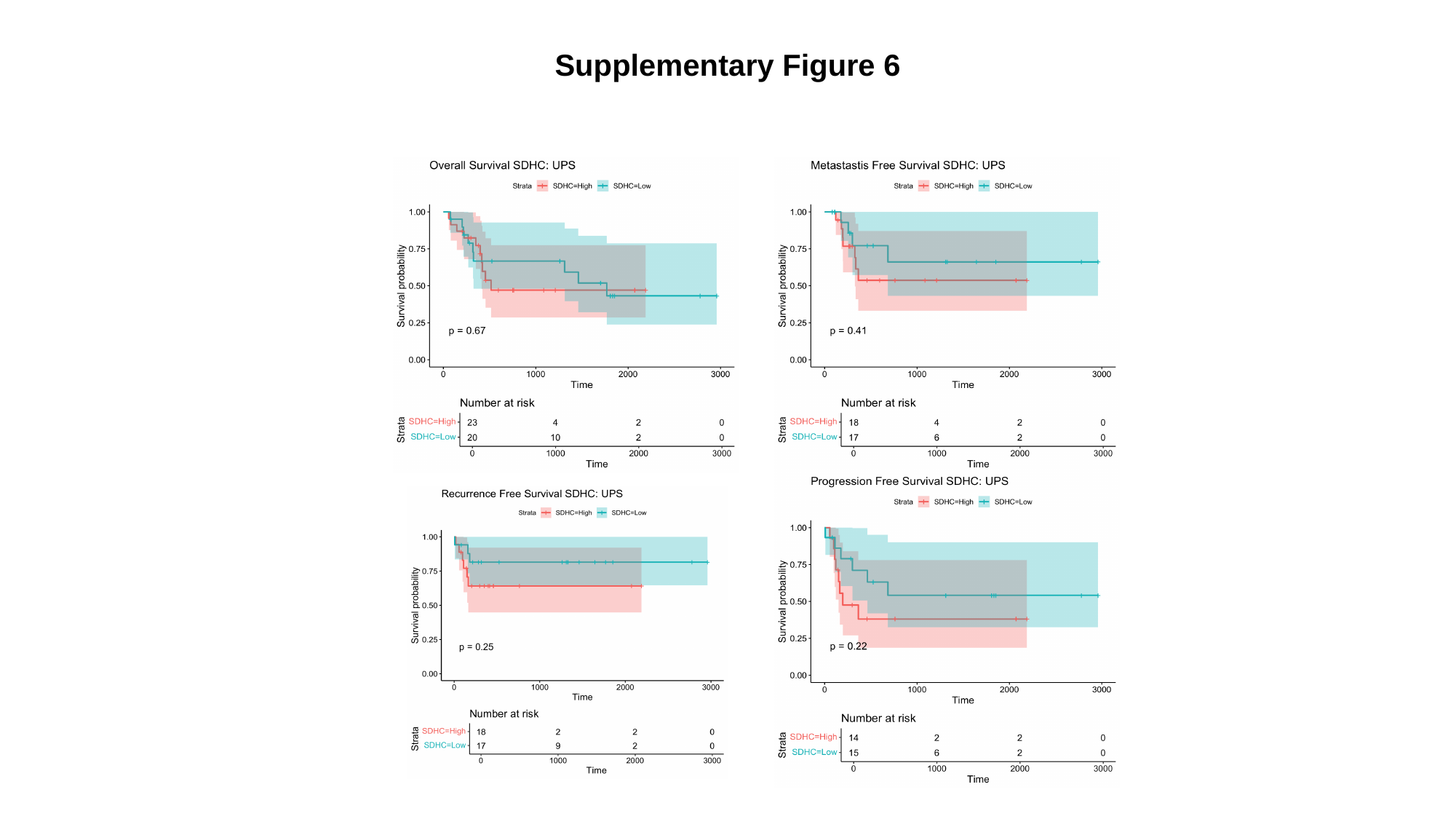

Supplementary Figure 6

### Slide 7
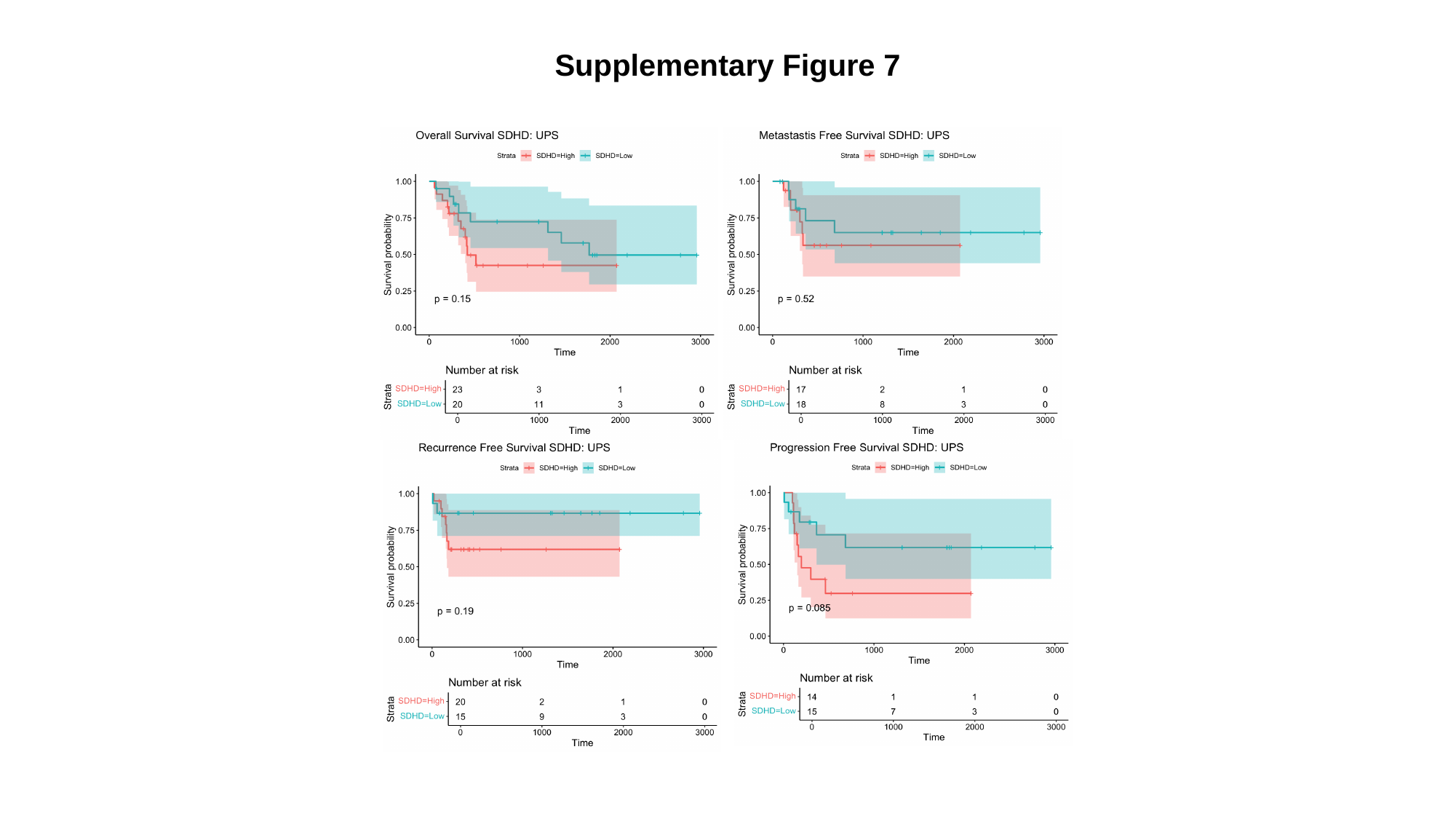

Supplementary Figure 7

### Slide 8
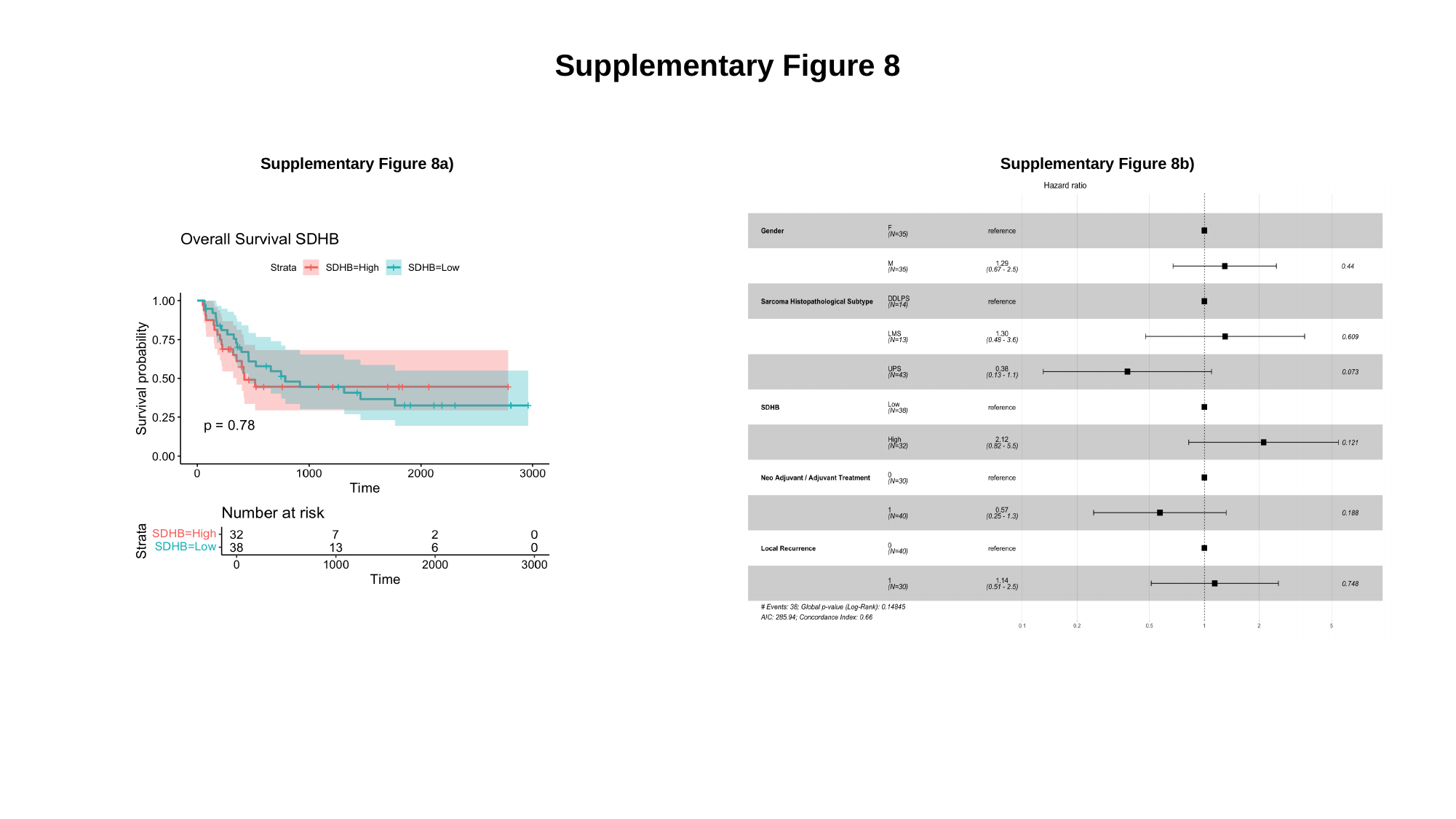

Supplementary Figure 8
Supplementary Figure 8a)
Supplementary Figure 8b)

### Slide 9
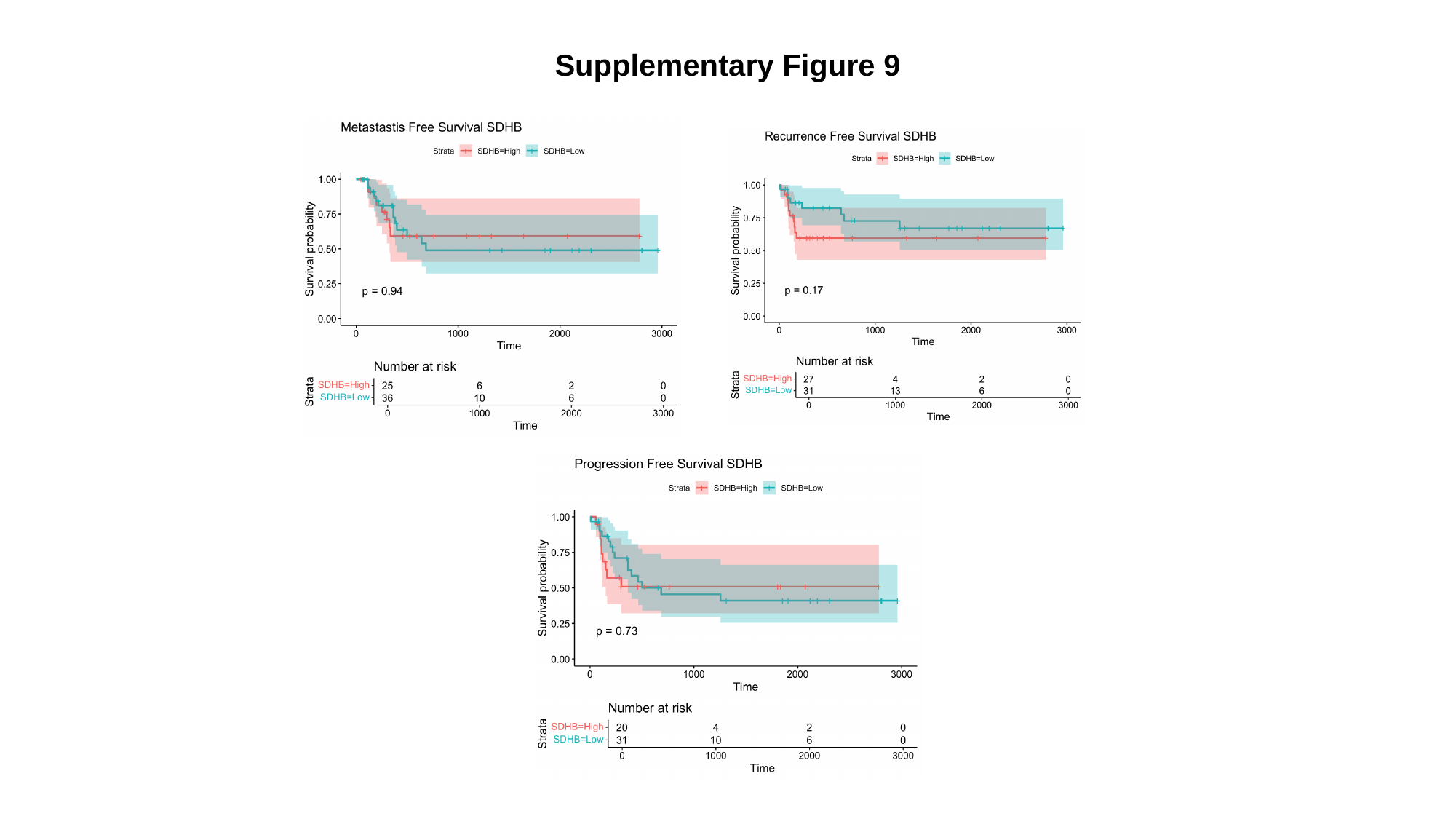

Supplementary Figure 9

### Slide 10
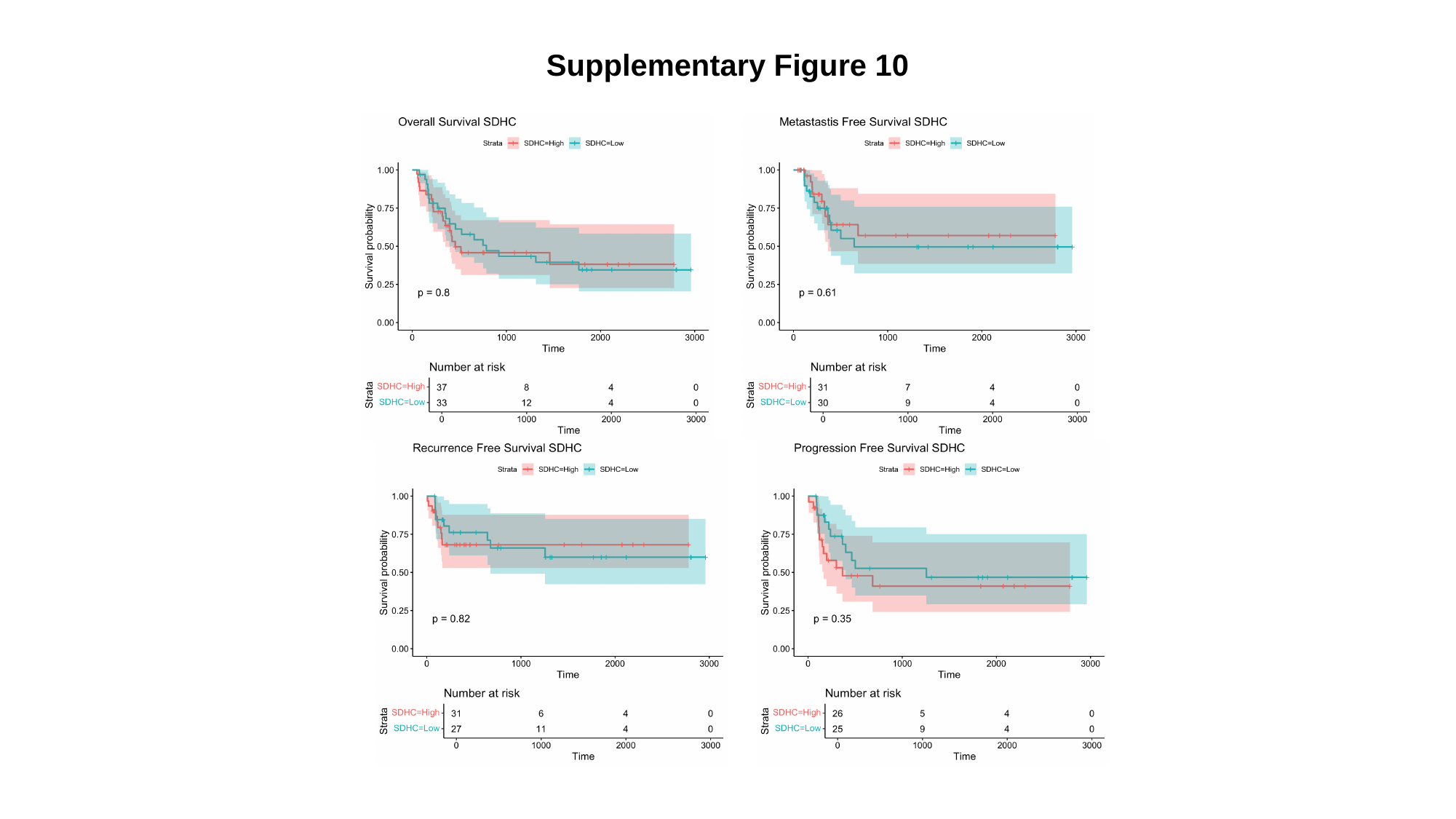

Supplementary Figure 10

### Slide 11
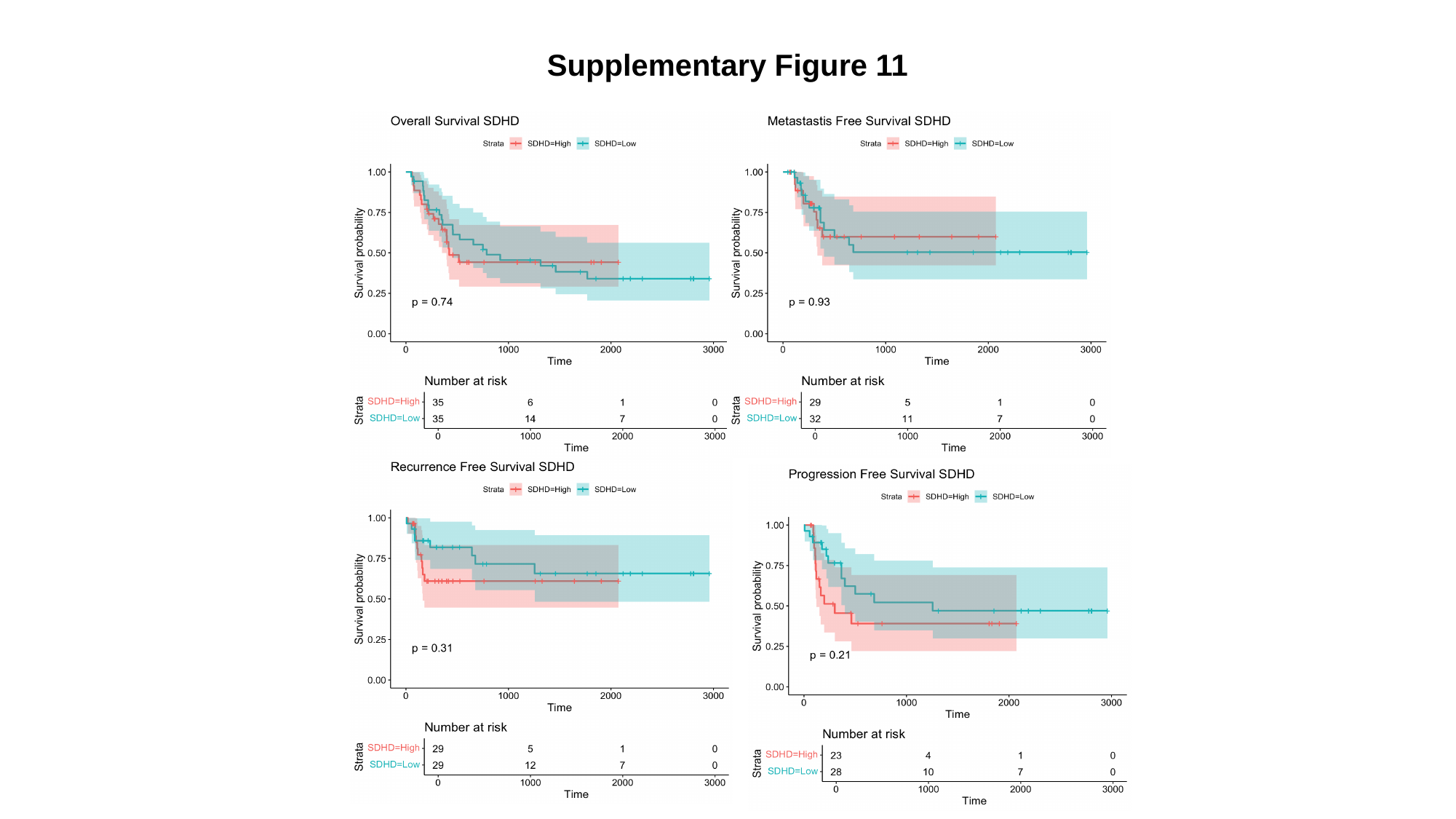

Supplementary Figure 11

### Slide 12
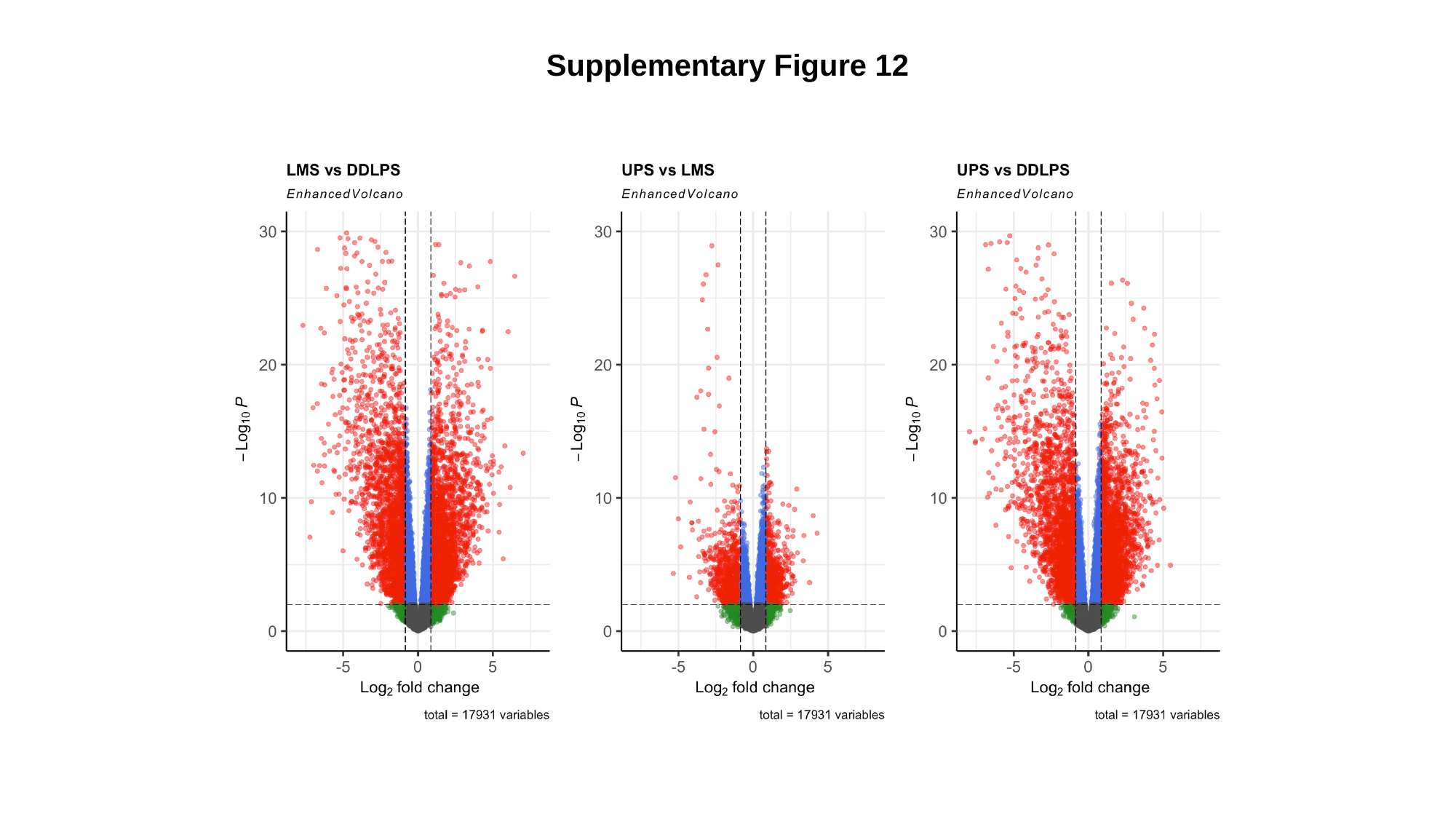

Supplementary Figure 12

### Slide 13
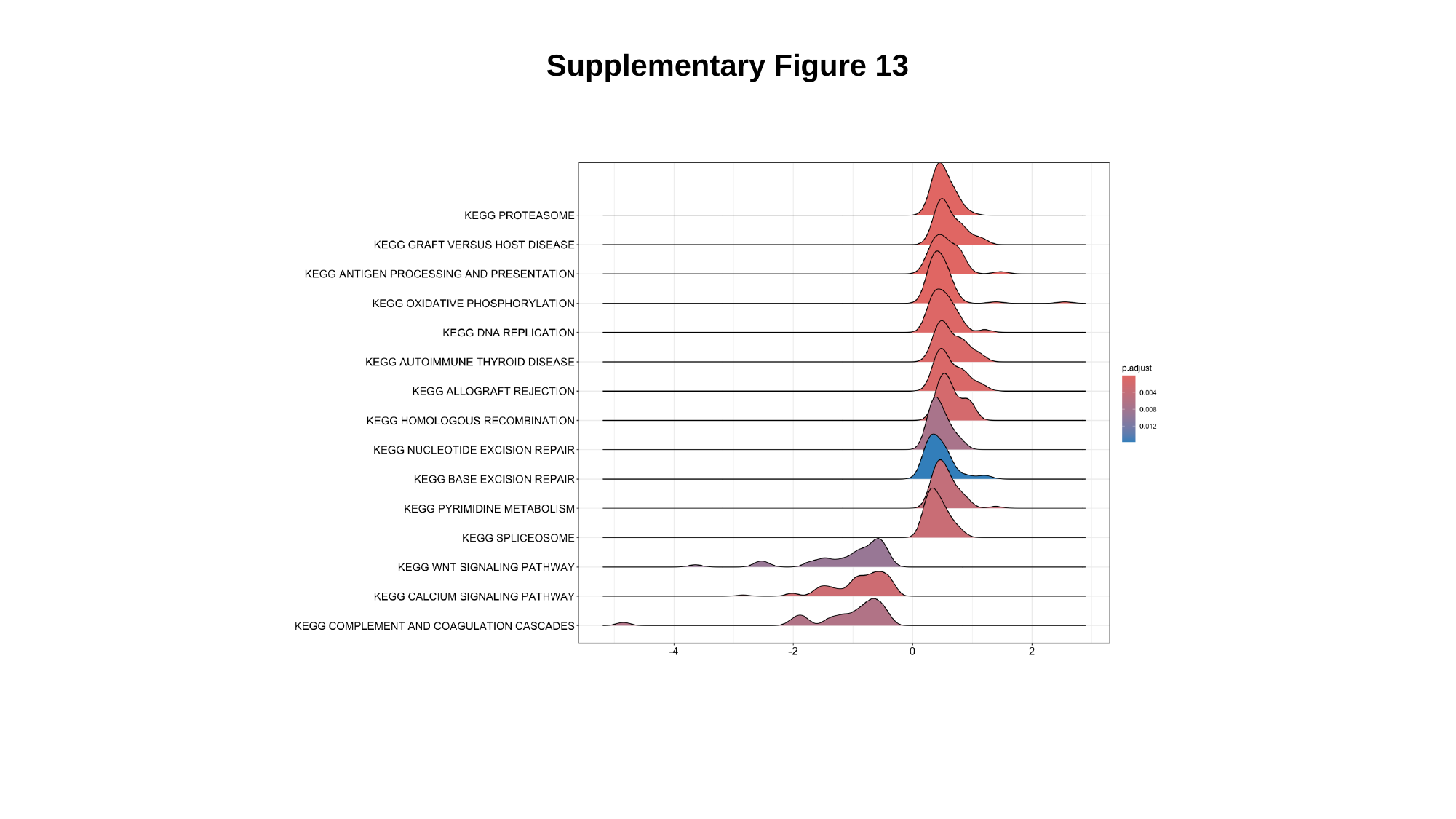

Supplementary Figure 13

### Slide 14
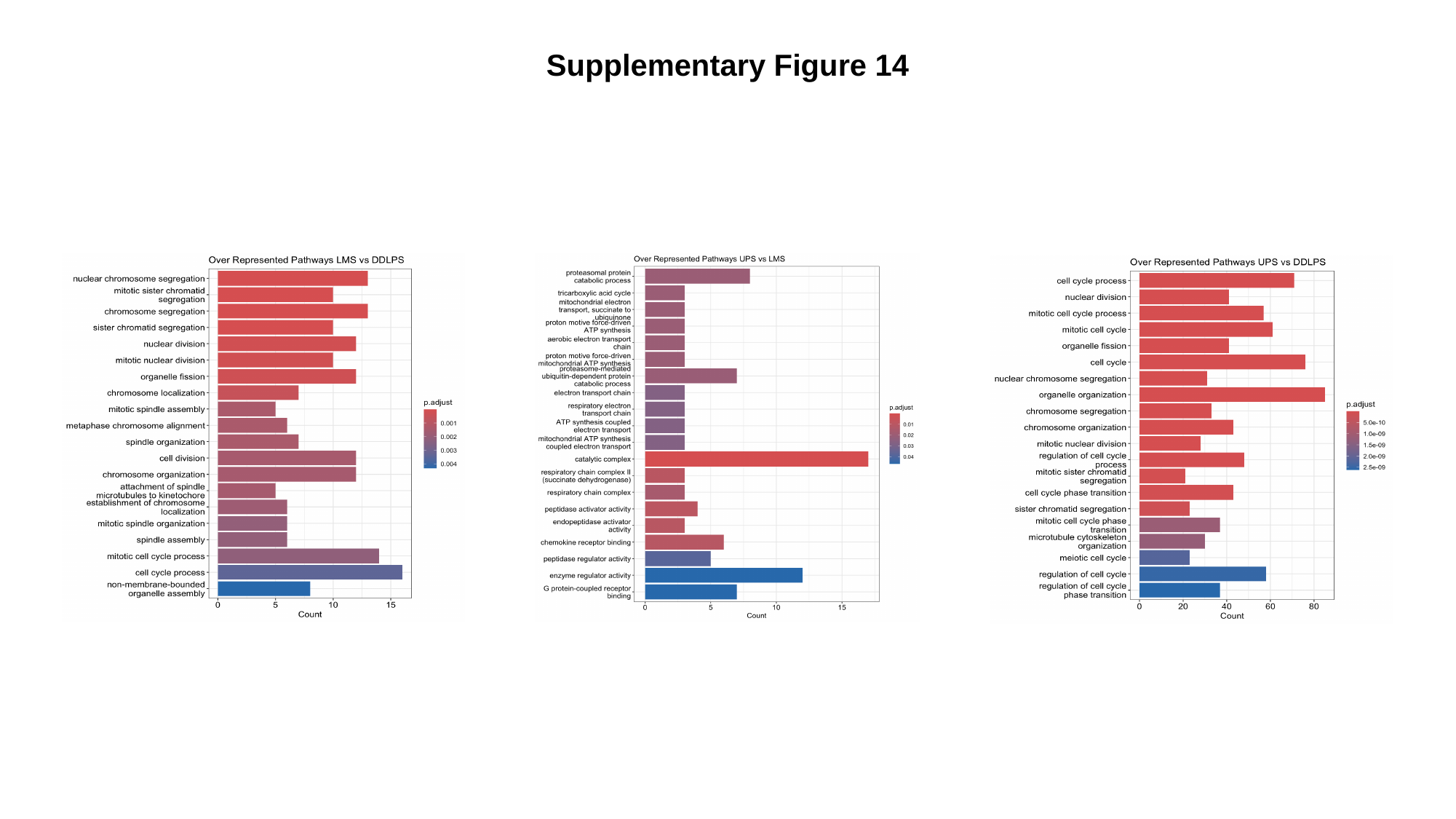

Supplementary Figure 14

### Slide 15
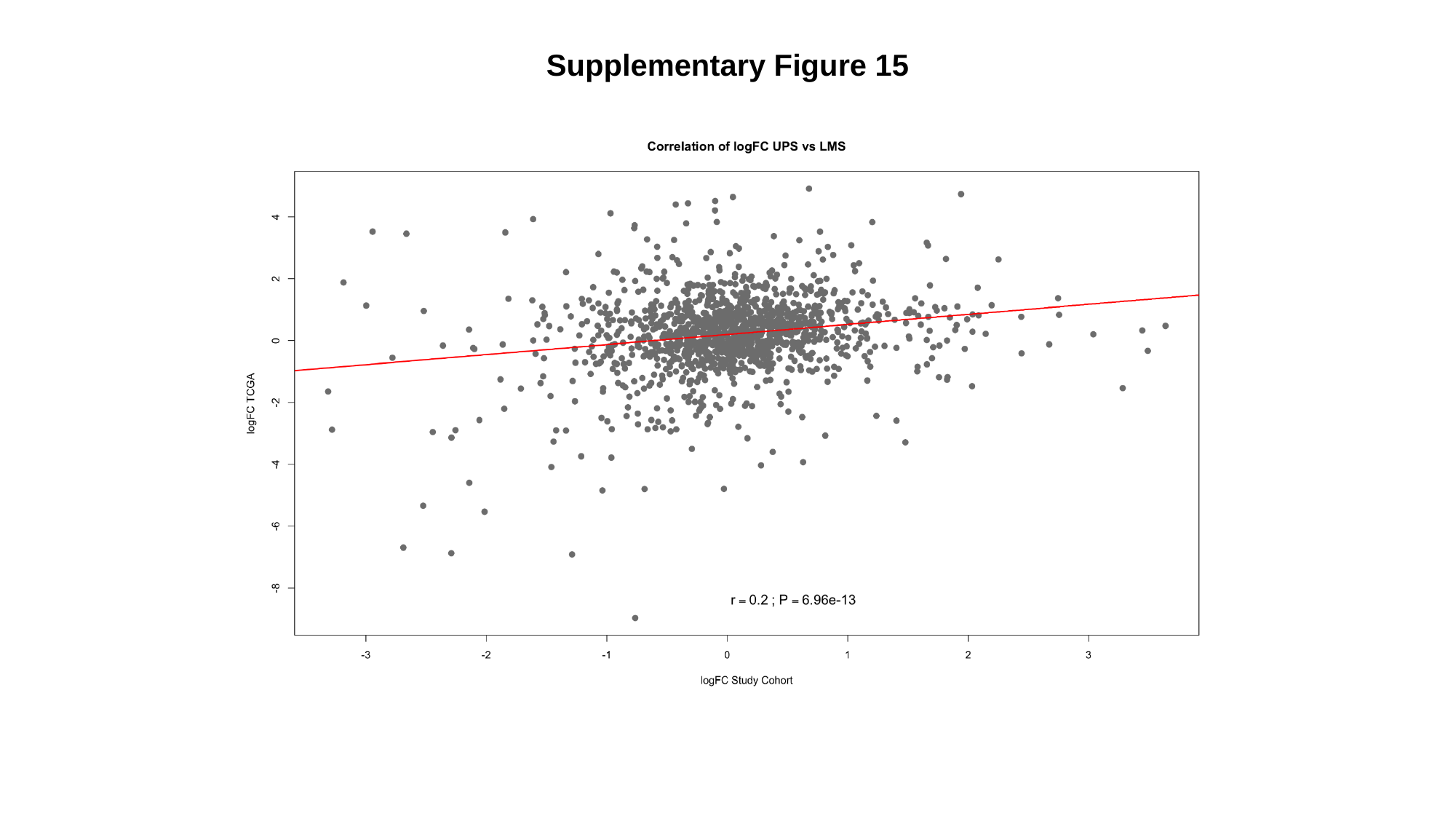

Supplementary Figure 15

### Slide 16
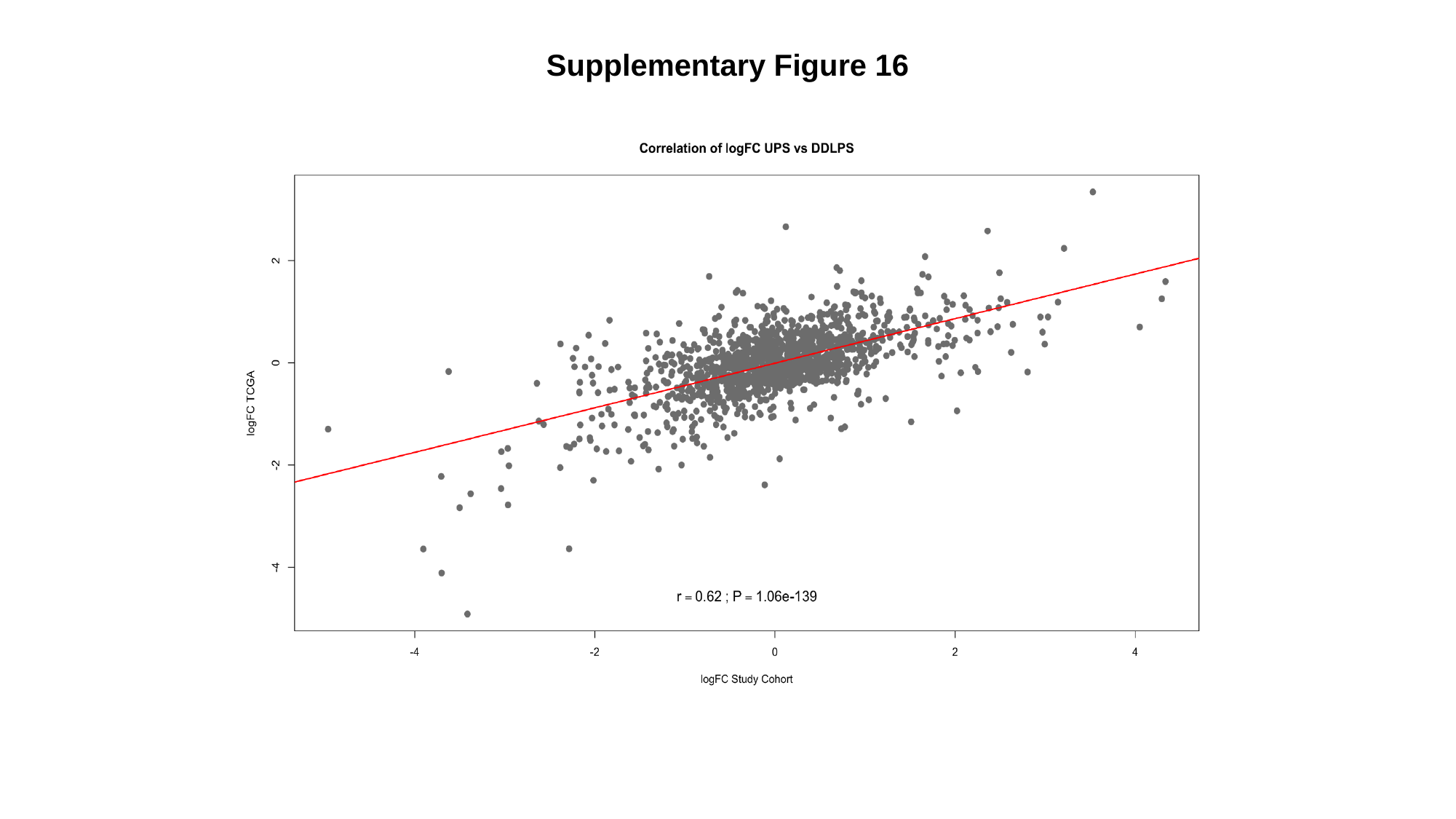

Supplementary Figure 16

### Slide 17
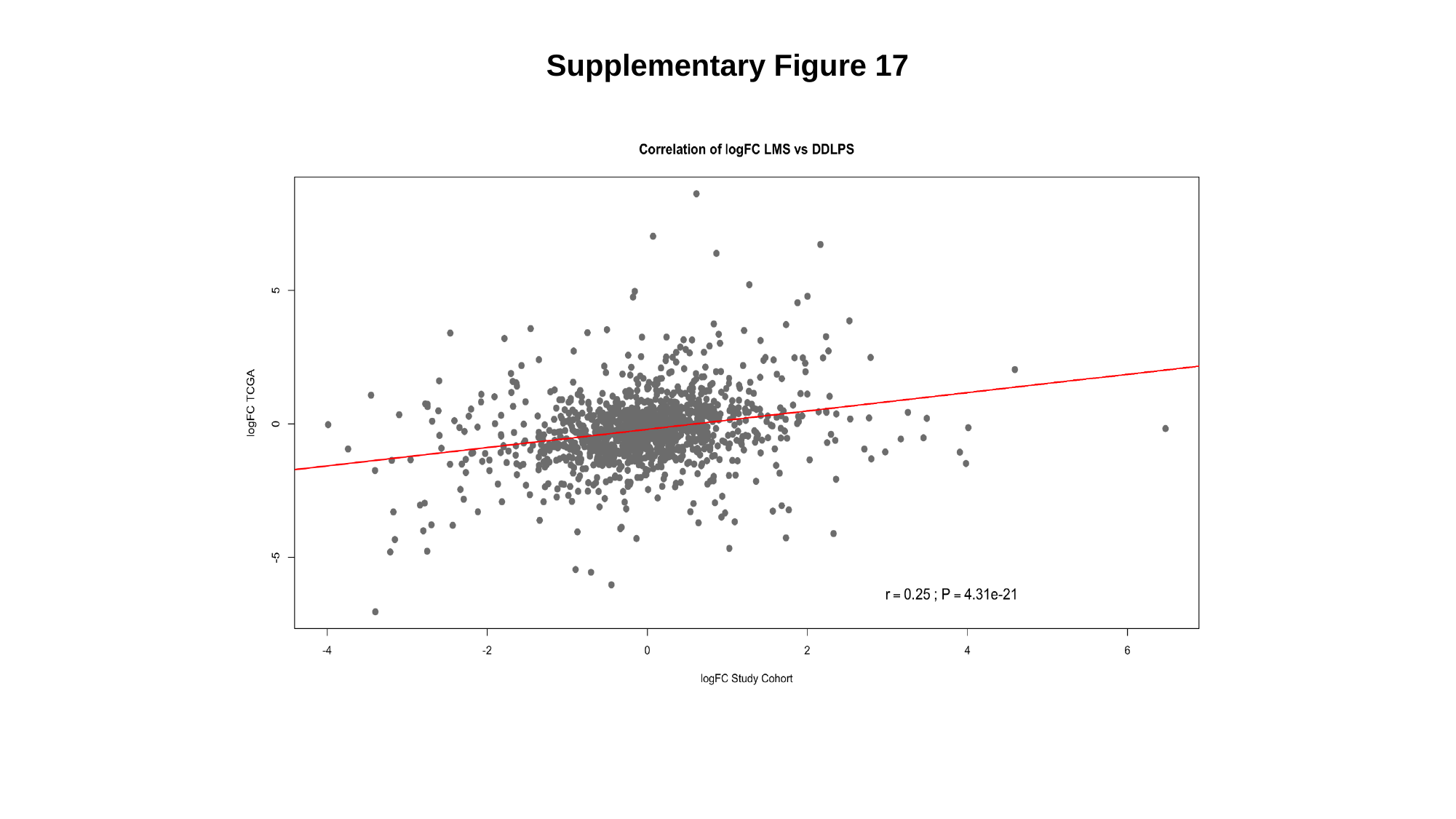

Supplementary Figure 17

### Slide 18
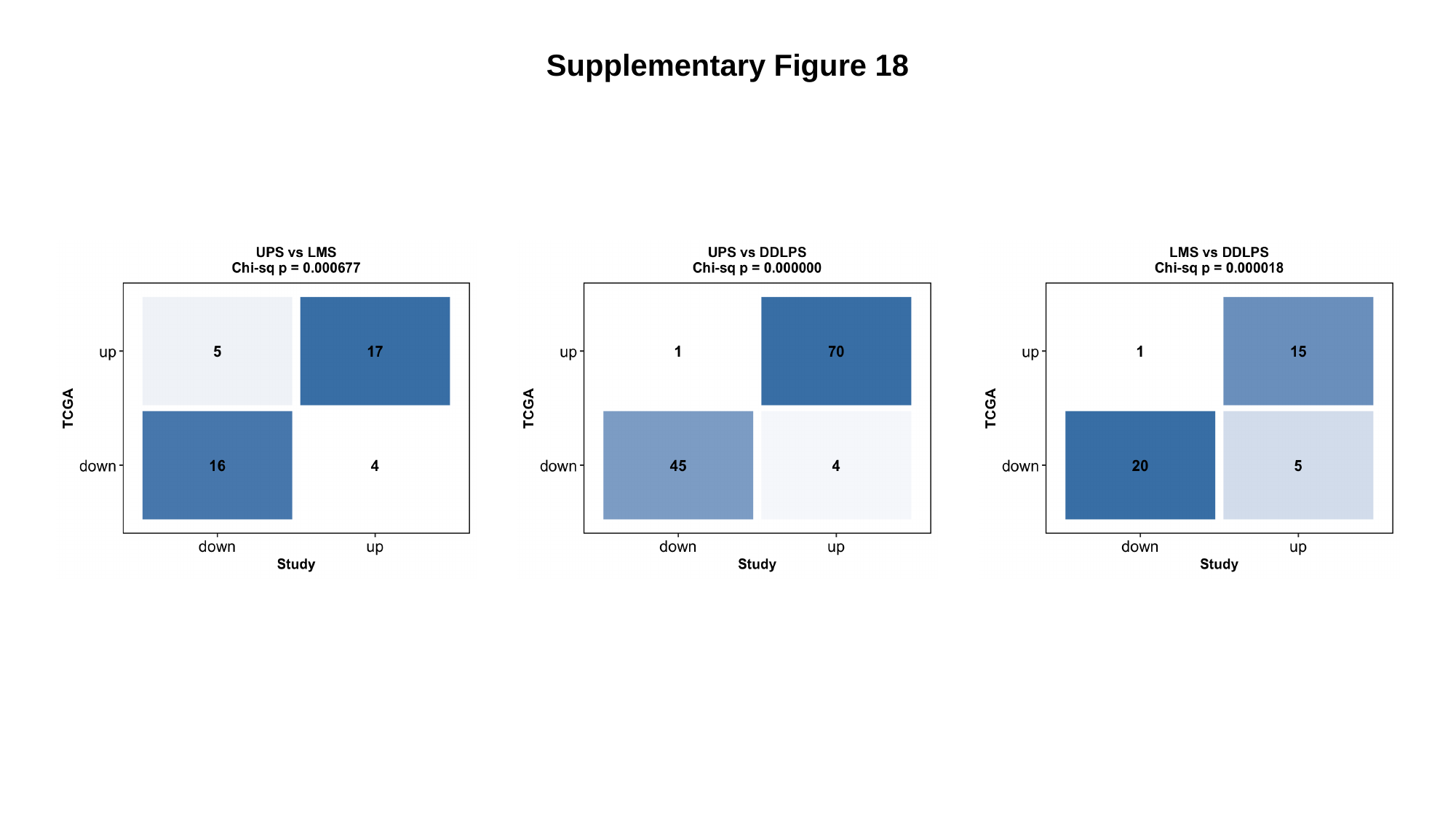

Supplementary Figure 18

### Slide 19
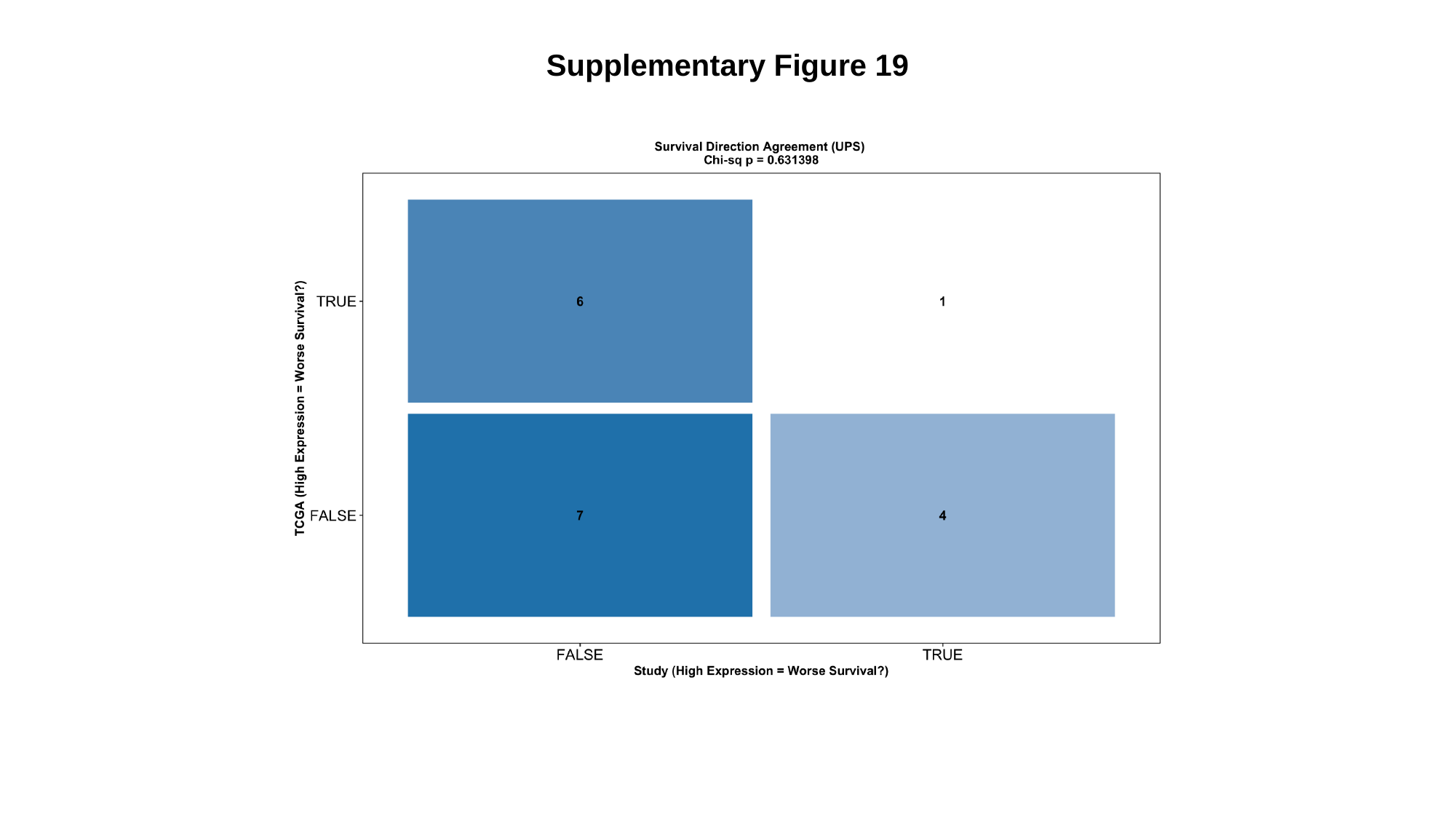

Supplementary Figure 19

### Slide 20
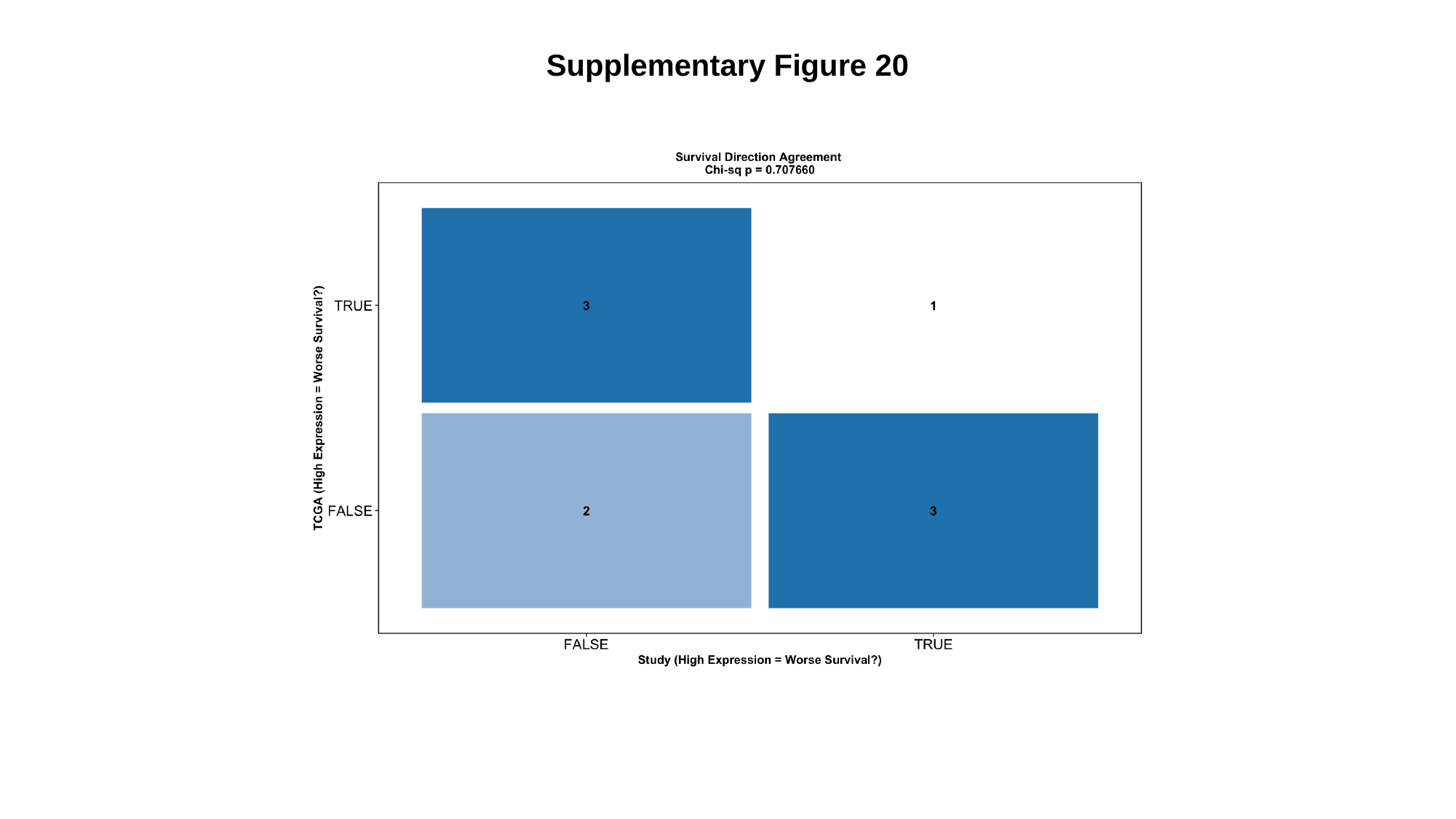

Supplementary Figure 20

### Slide 21
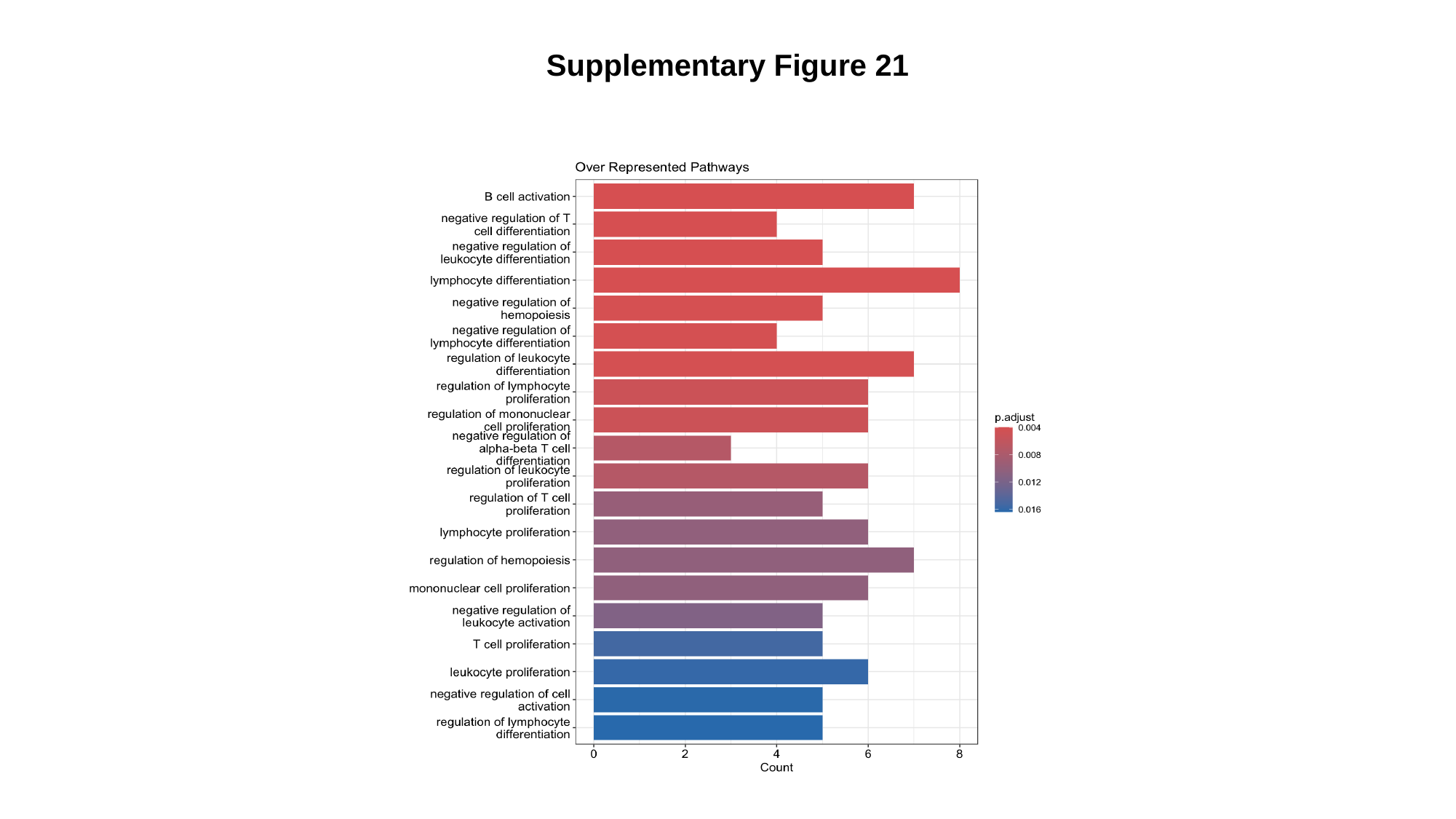

Supplementary Figure 21

### Slide 22
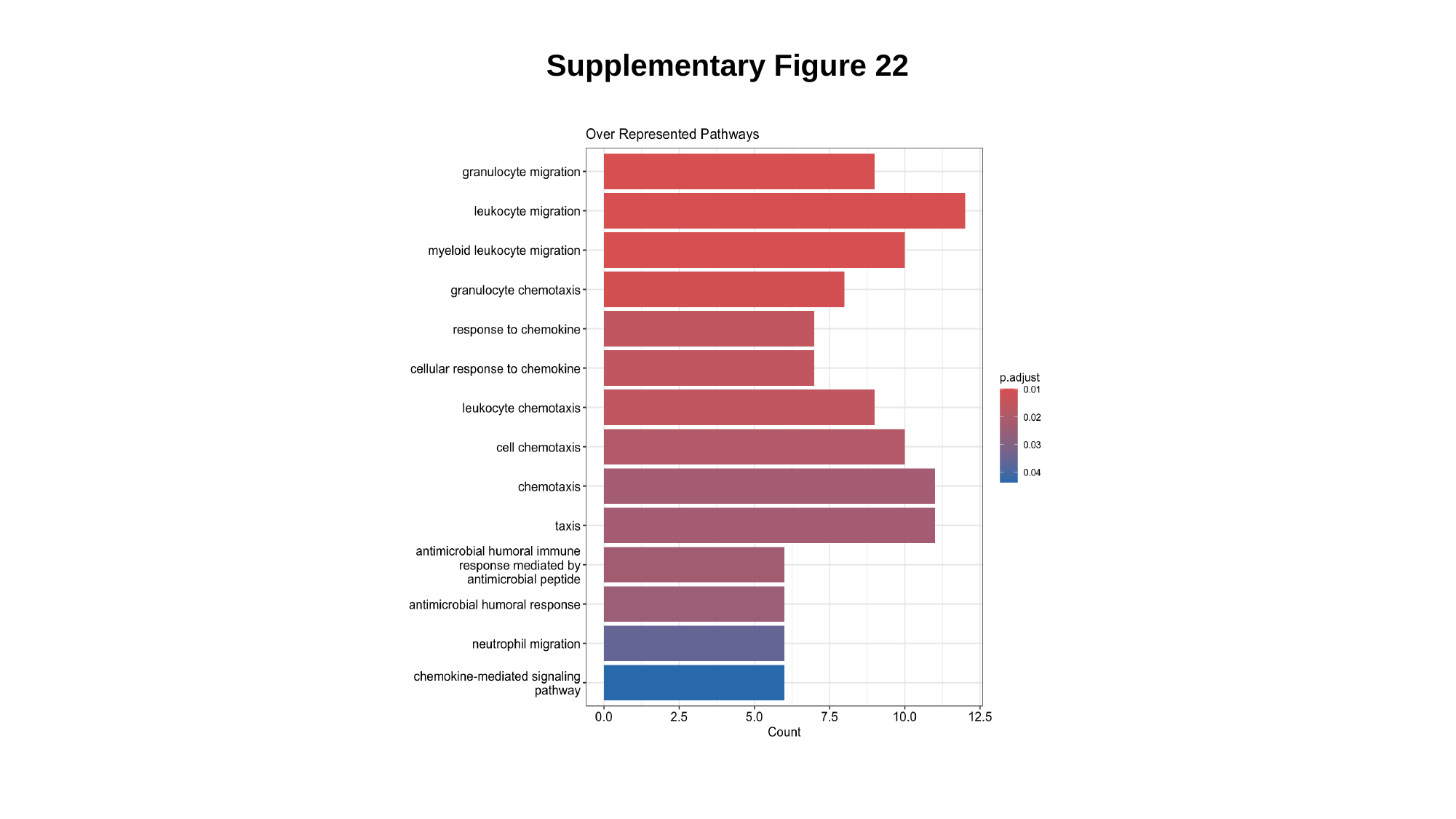

Supplementary Figure 22

### Slide 23
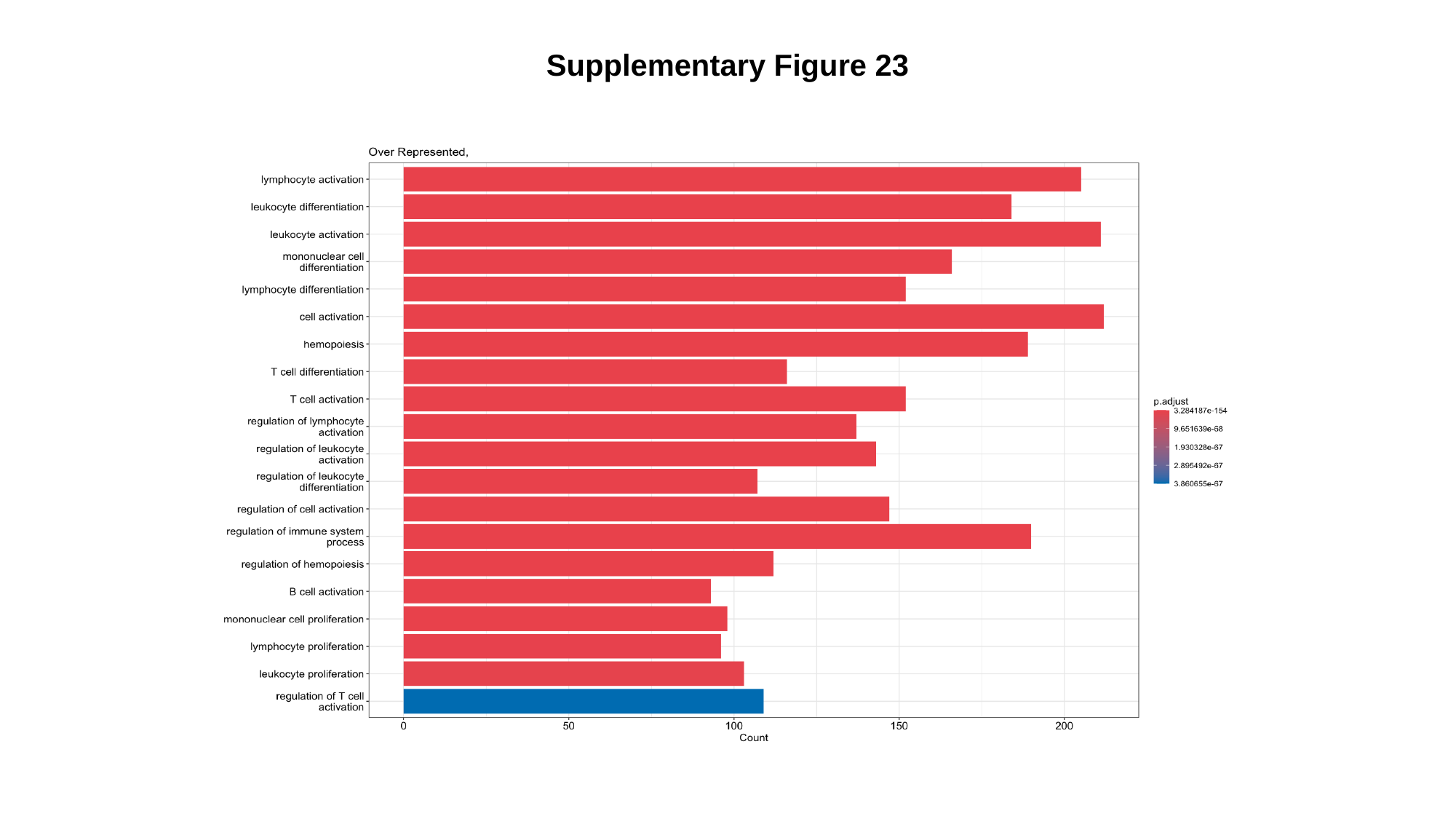

Supplementary Figure 23

### Slide 24
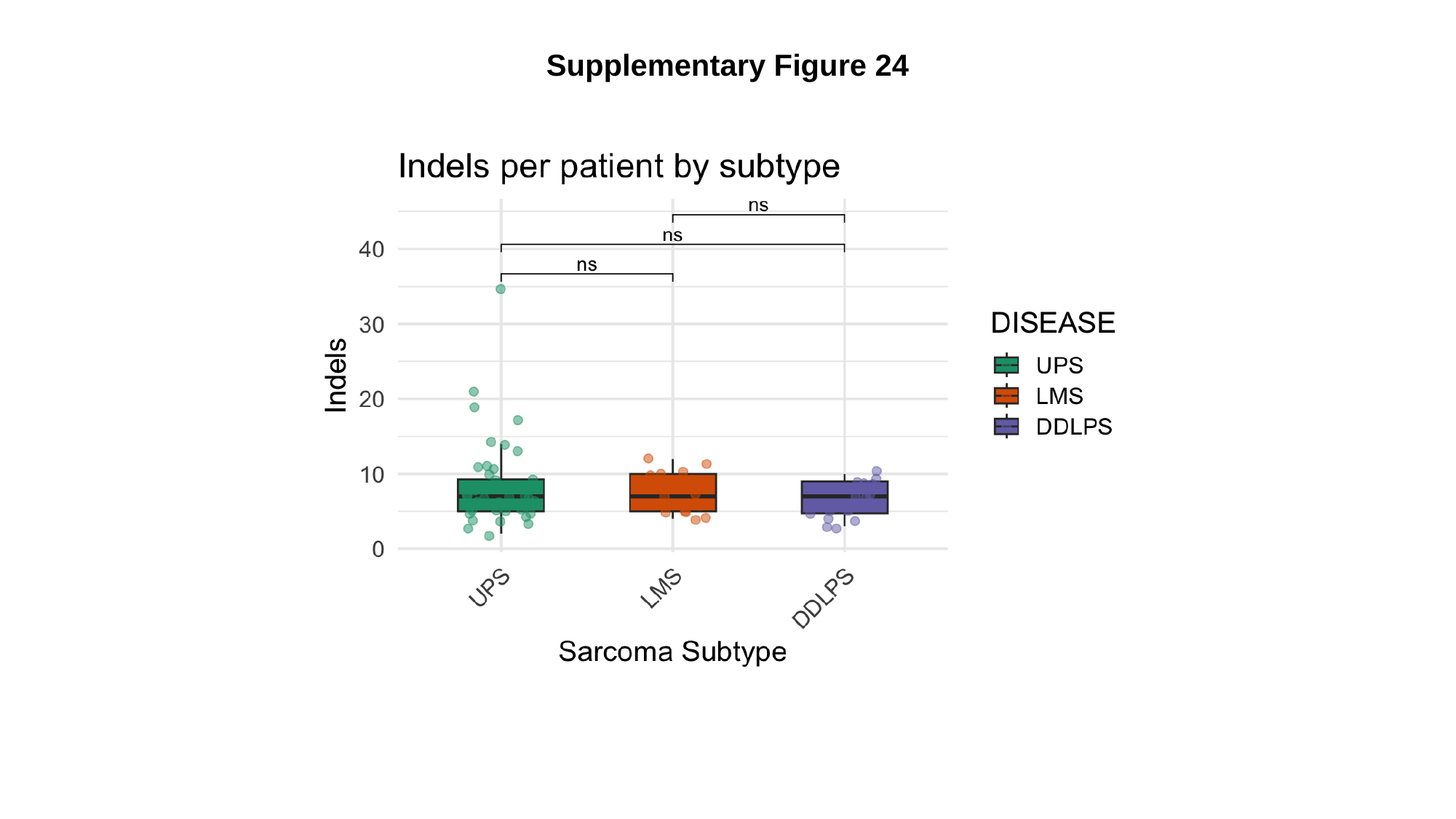

Supplementary Figure 24

### Slide 25
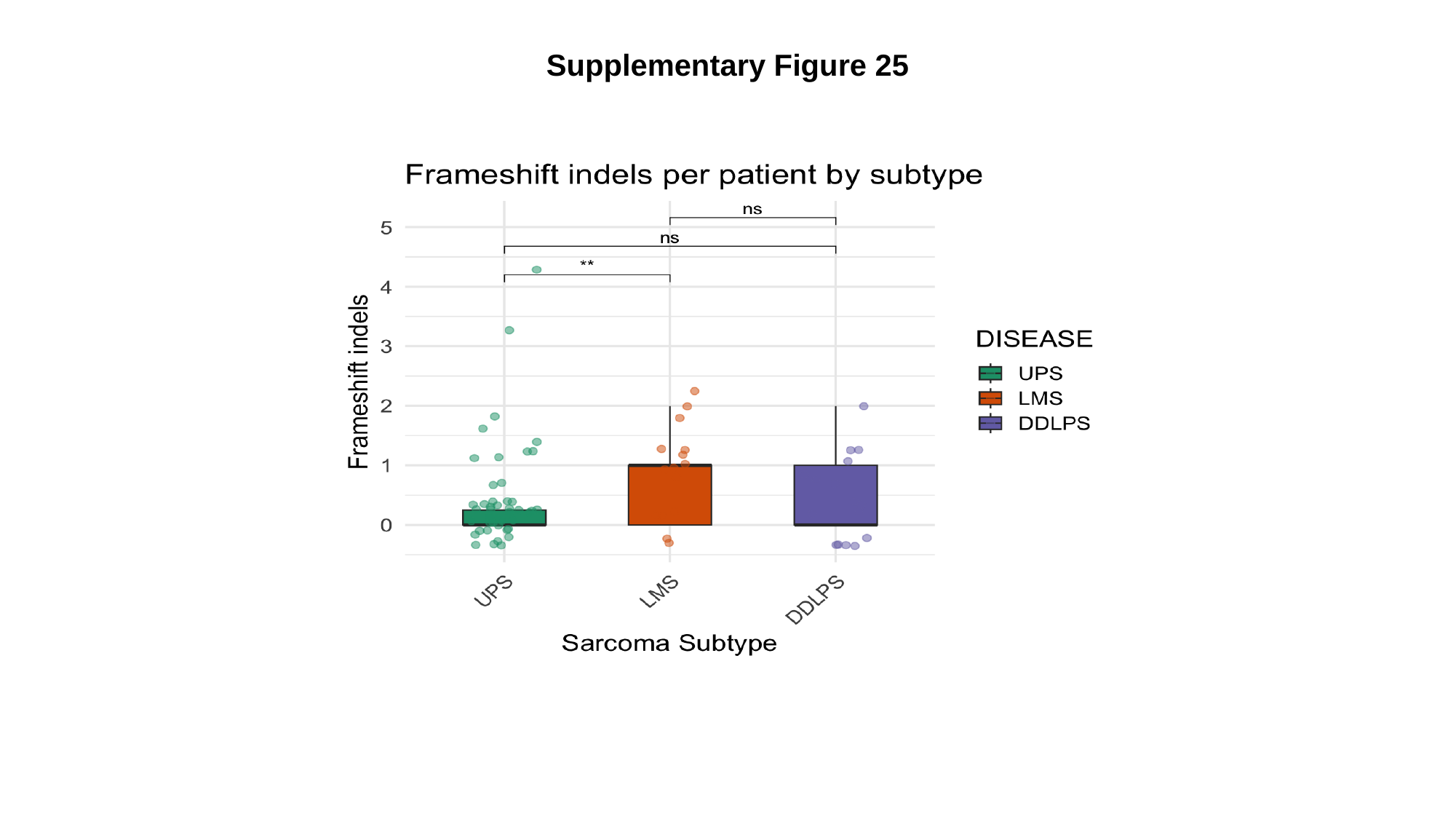

Supplementary Figure 25

### Slide 26
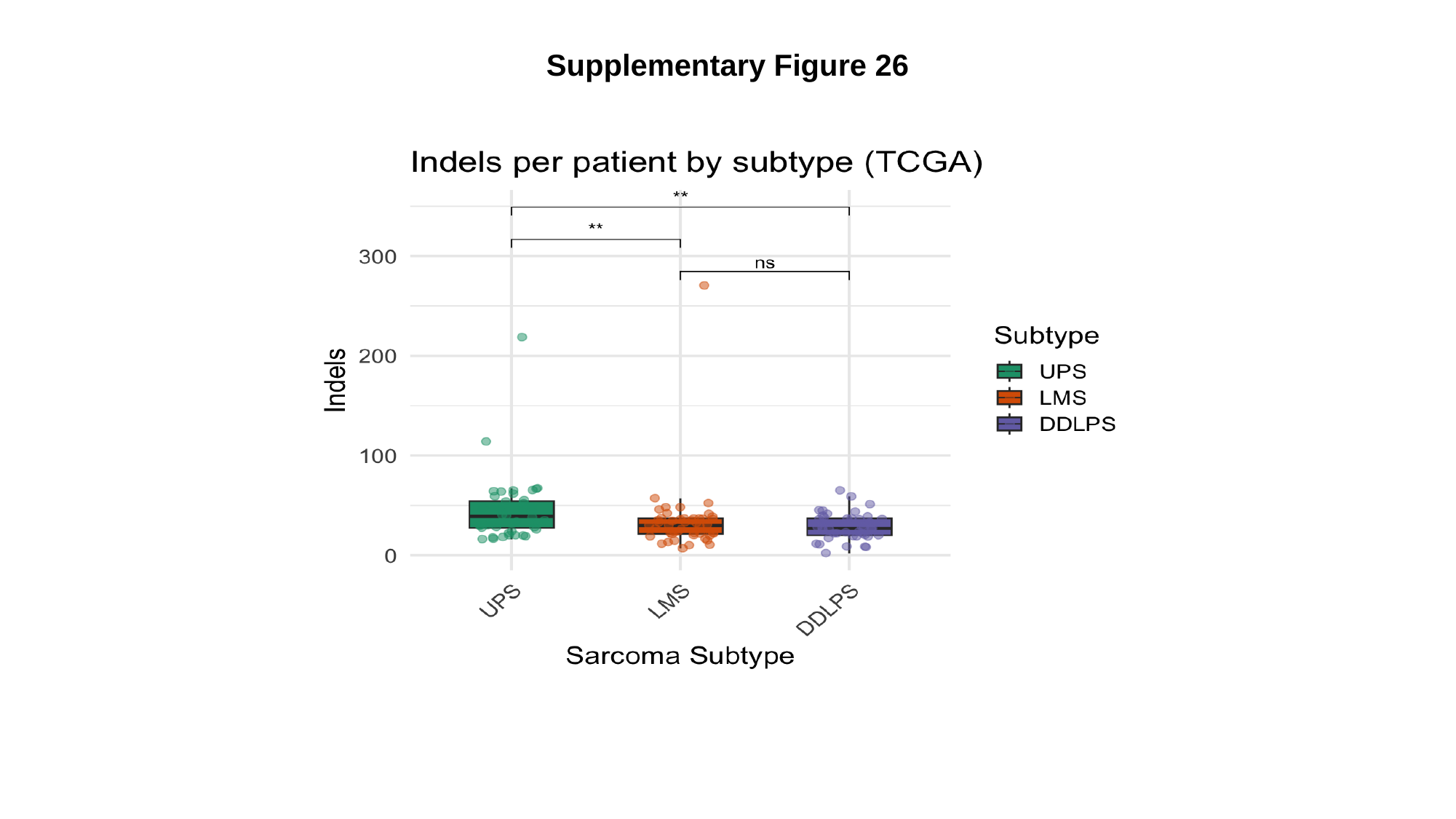

Supplementary Figure 26

### Slide 27
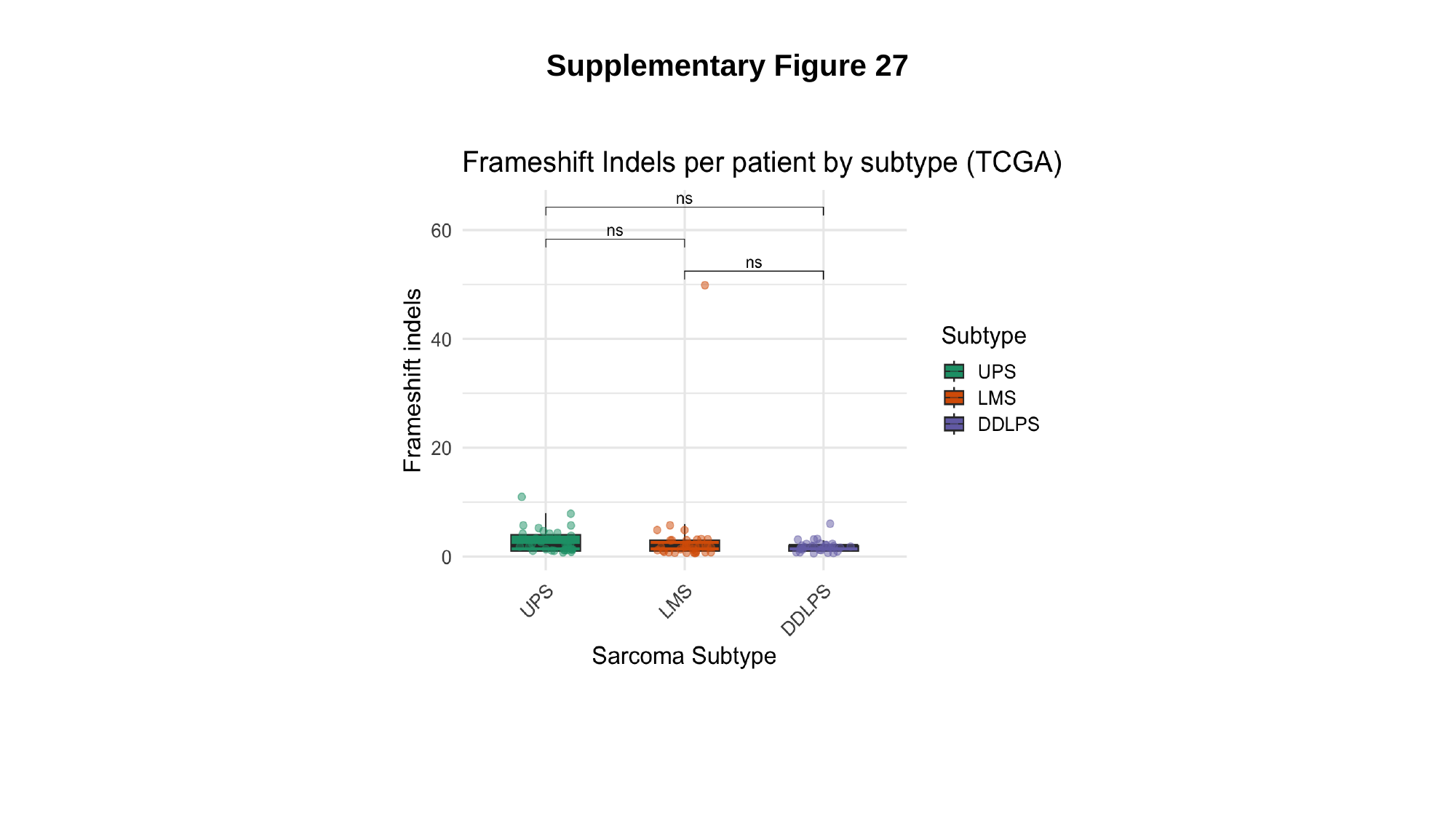

Supplementary Figure 27
