## Supplementary Tables and Figures for "Succinate dehydrogenase B (SDHB) overexpression with enzymatic dysfunction defines a distinct subtype of undifferentiated pleomorphic sarcoma"

### Supplementary Materials

### Results

Characterization of the cohort of STS patients whose samples were used for the exploratory metabolomic characterization using $^1$H NMR

This group of patients was composed by 16 patients with soft tissue or bone sarcomas. The median age at diagnosis was 74 years (interquartile range [IQR]: 33.4), and the majority were female (n = 11, 68.8%). Primary tumor localization was predominantly in the lower limbs (n = 7, 43.8%), followed by the upper limbs (n = 3, 18.8%) and the retroperitoneum (n = 3, 18.8%).

This set of fresh tissue samples featured a diverse range of histological subtypes, including: 4 UPS, 3 LMS, 2 LPS—1 DDLPS and 1 MLPS -, 1 DFSP, 1 SFT, 1 SS, 1 CS, 1 CCSLGT, 1 low-grade ESS, and 1 ADM.

Regarding tumor grade, the majority of tumors in the fresh tissue samples set were high-grade (G3) lesions (n = 9, 56.3%), followed by intermediate-grade (G2) tumors (n = 5, 31.3%) and low-grade (G1) tumors (n = 2, 12.5%).

Most tissue samples were obtained from primary tumor sites (n = 13, 81.3%), whereas three samples (18.7%) were derived from patients with metastatic disease at the time of sample collection.

All patients underwent surgical treatment. Surgical procedures were performed at both centers in all cases, with curative intent in 13 patients (81.2%) and palliative intent in 3 patients (18.8%). Among the patients treated with curative intent, 2 (15.4%) had been previously operated on at other institution, 4 (30.8%) had received neoadjuvant therapy, and all presented with resectable disease. The surgical margin status was R0 or R1 in all curatively treated cases. Adjuvant therapy was administered in 5 patients (38.5%), primarily radiotherapy (n = 4, 80.0%).

Oncological outcomes were evaluated in the 13 patients who underwent curative surgery, with a median follow-up duration of 24 months (IQR: 8.3). Of these, 2 patients (15.4%) had previously undergone surgical resection at another institution. The rate of local recurrence in this subgroup was 53.8% (n = 7), with a median time to recurrence of 6 months (IQR: 10.8). Similarly, the rate of distant metastasis was 53.8% (n = 7), with a median time to metastasis of 10 months (IQR: 9.0). The disease-free survival (DFS) rate was 30.8% (n = 4), while the overall survival (OS) rate was 69.2% (n = 9).

### Supplementary Tables

#### Supplementary Table 1 - Differentially expressed genes between UPS and high-grade LMS

| Gene | logFC | AveExpr | t | P.Value | adj.P.Val | B |
|---|---|---|---|---|---|---|
| **NFIC** | -0.789584423237439 | 9.85583721746982 | -4.6660585018346 | 1.27135446837404e-05 | 0.0120270391011854 | 3.04831067541231 |
| **STAT1** | 1.70934989082454 | 7.67779987726033 | 4.48488591618392 | 2.50911803223618e-05 | 0.0120270391011854 | 2.45239696777146 |
| **PPP1R13L** | -1.82984844077045 | 6.42612631774074 | -4.48740428565049 | 2.48574777197426e-05 | 0.0120270391011854 | 2.44230457271796 |
| **MAFB** | -1.20505318351186 | 7.96666758464597 | -4.3697395307354 | 3.83786404151358e-05 | 0.013082367784934 | 2.06379964871452 |
| **SDHD** | 1.059339669051 | 6.21377881179977 | 4.24624566850602 | 6.01514716959258e-05 | 0.013082367784934 | 1.64860486804697 |
| **BCAR1** | -0.916175634126613 | 7.09177556659966 | -4.21702103692396 | 6.68335435202448e-05 | 0.013082367784934 | 1.56626603886904 |
| **SP2** | -0.69359254201985 | 7.8139211508565 | -4.18824794918991 | 7.41088290066368e-05 | 0.013082367784934 | 1.46673965215019 |
| **FOXO4** | -0.962971458356956 | 6.79906087671984 | -4.17452813287969 | 7.78411833354592e-05 | 0.013082367784934 | 1.4295063347586 |
| **GNA12** | 0.884818280232242 | 6.24666348018837 | 4.12487634688578 | 9.2921968862282e-05 | 0.013082367784934 | 1.26517173990552 |
| **SDHC** | 0.96469603609239 | 7.92909864628879 | 4.10801208008077 | 9.86566121906347e-05 | 0.013082367784934 | 1.205319930679 |
| **MRVI1** | -1.40317465136939 | 6.90771964013597 | -4.03766343913373 | 0.000126468462537322 | 0.0134968252328725 | 0.99455365309176 |
| **DCAF12** | 1.04845431977809 | 6.48046987078603 | 4.03478162622433 | 0.000127755177330249 | 0.0134968252328725 | 0.986581195842436 |
| **ALOX12** | -1.05560818070521 | 6.79124530293581 | -4.0267637439263 | 0.000131401636481374 | 0.0134968252328725 | 0.96118142024031 |
| **CDK2** | 0.957753438981249 | 5.59209412999343 | 3.9339151282697 | 0.000181610719333151 | 0.0159918024729759 | 0.654646742801542 |
| **MALAT1** | -0.838050666754233 | 12.4567926229887 | -3.98169342015621 | 0.000153833792908748 | 0.0147475329468519 | 0.601171495377745 |
| **PSME1** | 0.916464511001152 | 7.63706827543938 | 3.89069144573472 | 0.000210830365549874 | 0.0159918024729759 | 0.523839166374308 |
| **ZEB1** | -1.25395266293234 | 5.07097883876647 | -3.89004964839513 | 0.000211296416541406 | 0.0159918024729759 | 0.488550038378418 |
| **PARP12** | 1.32167593196179 | 4.23223254928642 | 3.90113258224261 | 0.000203385222509571 | 0.0159918024729759 | 0.409523701816112 |
| **FBXW11** | 0.947713948809528 | 5.1460530867747 | 3.84600063469755 | 0.00024574975306906 | 0.0176694072456654 | 0.36736542687013 |
| **ELF4** | 0.723924242102099 | 8.85448493792498 | 3.82070855166854 | 0.00026789550627175 | 0.0183444637151799 | 0.271177837795681 |

| | | | | | | |
|---|---|---|---|---|---|---|
| **PSENEN** | 1.01102953566758 | 6.75695886193859 | 3.77620882743149 | 0.000311564142928 61 | 0.0184612552389121 | 0.192455209103178 |
| **PRPF8** | 1.09618469051861 | 7.27140011149265 | 3.77784495165756 | 0.000309844733984757 | 0.0184612552389121 | 0.185931968072728 |
| **IL4R** | -0.986530497173707 | 6.07832256813762 | -3.76742334291816 | 0.000320953672442839 | 0.0184612552389121 | 0.173224747830697 |
| **HDAC5** | -0.676570828491214 | 7.57194189812347 | -3.72785299537956 | 0.000366694020566 59 | 0.0188262309940631 | 0.0275723245293769 |
| **TPM3** | 0.694216188836872 | 11.3755175626795 | 3.77214774518086 | 0.000315871390119234 | 0.0184612552389121 | -0.00362359774743659 |
| **ZC3H13** | -0.787798795080736 | 9.142888222286 | -3.72786429478596 | 0.000366680113322768 | 0.0188262309940631 | -0.0274837140767881 |
| **CDC6** | 1.82431862649599 | 6.26393514089737 | 3.68804204713572 | 0.000418942553414477 | 0.0188262309940631 | -0.0608173889452077 |
| **GAS1** | -1.52418631375995 | 8.51699241897329 | -3.70734927716933 | 0.000392775286557459 | 0.0188262309940631 | -0.0628476546399499 |
| **SDHB** | 0.764410407423375 | 9.02510705733615 | 3.71232230256748 | 0.000386291805152101 | 0.0188262309940631 | -0.0693821590224903 |
| **IRF1** | 1.27297577574181 | 7.23333501388461 | 3.69167012540596 | 0.000413902031566874 | 0.0188262309940631 | -0.0720496622093973 |
| **ATAD2** | 1.54602842456617 | 4.00626755112027 | 3.71617429709067 | 0.000381340050188926 | 0.0188262309940631 | -0.130004799319943 |
| **CASP1** | 1.85420409956581 | 4.12127720538774 | 3.65454228399873 | 0.000468327259386067 | 0.019807488205799 | -0.276708110242507 |
| **APH1A** | 0.581090741306348 | 8.25216939630856 | 3.62723705738726 | 0.000512626868176715 | 0.021061641041089 | -0.293786118555732 |
| **DDX21** | 0.994664813116282 | 5.70929447492488 | 3.60066669496652 | 0.000559536211464 23 | 0.0218976507729986 | -0.311205882892391 |
| **TCL1A** | -2.97087493747756 | -1.79053088752711 | -4.10399013066376 | 0.000100073745225503 | 0.013082367784934 | -0.313873212014557 |
| **CNN1** | -2.22374270952542 | 5.50285040591746 | -3.50920766275827 | 0.000754108360327525 | 0.0264489712719752 | -0.5690223365955 7 |
| **LPP** | -0.655660519492356 | 10.4622602159627 | -3.56133166060312 | 0.000636525052651795 | 0.0240874480450863 | -0.598332375619616 |
| **PVRIG** | -1.06694254408491 | 5.8573433177247 | -3.46838867979067 | 0.000860250108755257 | 0.0274897701420013 | -0.681663797165902 |
| **CD244** | 2.65930632921841 | 1.71320393218464 | 3.67535404065576 | 0.000437033550352471 | 0.0190440680426319 | -0.683594133088697 |
| **JAK1** | 0.65614998541497 | 12.1126984491325 | 3.54465046704332 | 0.000672119935570723 | 0.0243704299529685 | -0.748521223813842 |
| **CDK4** | 0.87345473634057 | 11.0481320094943 | 3.4748031488797 | 0.000842683477142154 | 0.0274897701420013 | -0.888439993732616 |

| | | | | | | |
|---|---|---|---|---|---|---|
| **ACTG2** | -2.6359879599009 | 4.68324949873384 | -3.3872922275652 | 0.00111437323395613 | 0.0333847648006025 | -0.923635082785917 |
| **LY6E** | 1.13708408653432 | 8.03043097194872 | 3.38942957096463 | 0.00110685092309864 | 0.0333847648006025 | -0.976196471230746 |
| **CXCL11** | 2.49141070213015 | 2.2690675813549 | 3.50109567787524 | 0.000774161770433262 | 0.0265058244257865 | -0.97647470619912 |
| **ARHGAP15** | 3.43187615413257 | 1.51736308621025 | 3.54202198108126 | 0.000677897912460875 | 0.0243704299529685 | -1.01342520317702 |
| **C5** | 3.5078087410777 | 0.598295495425664 | 3.59855700781313 | 0.00056343051363 07 | 0.0218976507729986 | -1.02012080246691 |
| **CLTC** | 0.511164864309173 | 12.3423770107446 | 3.44804869951008 | 0.000918270080139964 | 0.0287059212008971 | -1.05184162054194 |
| **FUCA1** | 1.03798838745945 | 5.59456238700002 | 3.32613196733803 | 0.00135118957201456 | 0.0378602976043414 | -1.06536251235048 |
| **NOS1AP** | -2.45732542633202 | 1.9530615040011 | -3.48254423356801 | 0.000821935371830395 | 0.0274870480160955 | -1.07288547900803 |
| **CXCL10** | 1.91976595871566 | 5.19974482256587 | 3.30707438057998 | 0.00143416558204807 | 0.0378729290261139 | -1.11552401419339 |
| **MEGF9** | -0.578896185674568 | 8.82547273090993 | -3.33098686349743 | 0.00133078489236414 | 0.0378602976043414 | -1.17736835702136 |
| **KDR** | 1.58360578160831 | 5.26574865641108 | 3.27655099448382 | 0.00157710768051356 | 0.0397873832382194 | -1.19468191913993 |
| **NXPH3** | -1.41398602160162 | 3.75280976665206 | -3.30387703484765 | 0.00144854735496263 | 0.0378729290261139 | -1.20982035859615 |
| **SKP1** | 0.892900007857872 | 6.91753203598304 | 3.28036250496056 | 0.0015585542491817 | 0.0397873832382194 | -1.23593015327085 |
| **RGCC** | -0.956792626419003 | 5.96359606347053 | -3.25184259355131 | 0.00170250101123396 | 0.0414948551551598 | -1.27133192345584 |
| **DHH** | -1.10162471330505 | 2.44936375831611 | -3.34741048917319 | 0.00126388611787112 | 0.0370911885203809 | -1.29166857229547 |
| **CCL8** | 2.00832830371516 | 3.73866146947965 | 3.22463307685507 | 0.00185137419629623 | 0.042939937004419 | -1.40097944845372 |
| **NKD1** | -2.16578714451691 | 4.52967115004595 | -3.1827254890963 | 0.00210470113462438 | 0.0465624651013824 | -1.44828305697936 |
| **PSMB8** | 0.92145054852312 | 6.55759138665133 | 3.18634477040592 | 0.00208160555928537 | 0.0465624651013824 | -1.47173801110651 |
| **SS18** | 0.56681038858023 | 10.9421447558634 | 3.26687784383266 | 0.00162512890897367 | 0.0402919891569679 | -1.48156499708501 |
| **APOBR** | -1.14393285048069 | 9.82998442272165 | -3.22641851570226 | 0.0018412431489761 | 0.042939937004419 | -1.52851940827563 |

| | | | | | | |
|---|---|---|---|---|---|---|
| **WNT16** | -2.50233022533956 | 0.970347960667151 | -3.30654005429618 | 0.00143655962509844 | 0.0378729290261139 | -1.5823718649353 8 |
| **YY1** | -0.458134303470864 | 7.62317518370726 | -3.16005079335066 | 0.00225493397349069 | 0.0483969411026809 | -1.5916594752995 9 |
| **NEAT1** | -0.695691659265238 | 12.0076595692596 | -3.2335635231698 | 0.00180121795092807 | 0.042939937004419 | -1.6468655919427 5 |
| **CXCR5** | -1.58756790938665 | 1.1947580572389 | -3.19652340335262 | 0.00201791717828504 | 0.046059760355141 | -1.7795925552787 2 |
| **CDX2** | -2.38894959864585 | -3.35859924892717 | -3.32193193884522 | 0.00136907891197897 | 0.0378602976043414 | -1.9811986828531 5 |
| **IHH** | -2.10299166365811 | -1.11174763770739 | -3.16345595024607 | 0.00223175098824757 | 0.0483969411026809 | -2.0443520825844 2 |

**Supplementary Table 2** - Differentially expressed genes between UPS and DDLPS

| Gene | logFC | AveExpr | t | P.Value | adj.P.Val | B |
|---|---|---|---|---|---|---|
| **MDM2** | -3.43687632132414 | 8.29318909997119 | -13.3168093110995 | 1.12693473209516e-21 | 1.62053214475284e-18 | 38.2628546199005 |
| **CDK4** | -2.29283312838111 | 11.0481320094943 | -9.47684030878701 | 1.44554429982583e-14 | 1.03934635157477e-11 | 22.8024759412912 |
| **CCNB1** | 2.09232664455016 | 6.16107552618726 | 7.2092996753437 | 3.32441952102088e-10 | 1.59350509040934e-07 | 12.965833102946 |
| **CLTC** | 0.973536472632007 | 12.3423770107446 | 6.82335493341451 | 1.78981982825169e-09 | 6.43440228256481e-07 | 11.3022428777154 |
| **CCND2** | -3.04904442754487 | 7.51636847893976 | -6.68786625725138 | 3.21774833852623e-09 | 9.25424422160144e-07 | 10.8631051249955 |
| **WHSC1** | 1.38230927351389 | 9.96797152236539 | 6.35518001830884 | 1.34246513058173e-08 | 3.21744142962755e-06 | 9.44405660524049 |
| **ANXA2** | 1.54081426376126 | 7.57648312659572 | 6.12511576914354 | 3.56391389773567e-08 | 7.3212974070627e-06 | 8.56552375023907 |
| **KIF23** | 1.94489456591771 | 7.0699488629083 | 6.04385982191768 | 5.01865976039145e-08 | 9.02104091930363e-06 | 8.2437856021799 |
| **TPM3** | 1.05259703273507 | 11.3755175626795 | 5.94433704921053 | 7.6177234464948e-08 | 1.21714292400661e-05 | 7.69873500066388 |
| **ZMAT3** | -1.45812405562422 | 7.00743750286118 | -5.85012571717193 | 1.12847714610337e-07 | 1.4752273964515e-05 | 7.47414114692098 |
| **AURKA** | 2.58473371922525 | 5.46159656917431 | 5.85402622526248 | 1.11031123429752e-07 | 1.4752273964515e-05 | 7.46109664210979 |
| **ELF4** | 1.05657748103714 | 8.85448493792498 | 5.8044031389142 | 1.36456217441891e-07 | 1.63520033901199e-05 | 7.25005007755985 |
| **NRAS** | 1.24171341270049 | 9.24542481988066 | 5.50243151215441 | 4.72146984296236e-07 | 5.22267202629221e-05 | 6.04131786334106 |
| **CCNB2** | 1.87281831899367 | 4.96628794243036 | 5.41219494499644 | 6.8089341572695e-07 | 6.99374808439539e-05 | 5.75005899623829 |
| **RRM2** | 1.92464976834381 | 6.53079159814559 | 5.38093905166572 | 7.72529026100944e-07 | 7.40597826355439e-05 | 5.66092591562009 |
| **CDCA5** | 1.49176657234836 | 7.46693861182796 | 5.21328175867733 | 1.51309148234315e-06 | 0.000134856390425326 | 4.99891193804802 |
| **TOP2A** | 1.67180795332205 | 6.17158009105779 | 5.20015578260586 | 1.59426887150941e-06 | 0.000134856390425326 | 4.98724534661351 |
| **CDC20** | 2.07806572687036 | 5.74632963522538 | 5.16205985049827 | 1.85480324786655e-06 | 0.000146621550350762 | 4.85095173774474 |
| **CDKN3** | 1.74263160168204 | 6.37308000991613 | 5.13290071671133 | 2.0819722334271e-06 | 0.000149693803583409 | 4.73157540181996 |
| **KIF2C** | 3.0481937988861 | 5.53678481188073 | 5.04590274877661 | 2.93395306192605e-06 | 0.000191773841047712 | 4.42490200665526 |
| **TPX2** | 1.79117202523813 | 5.5062293775195 | 5.01660822301251 | 3.29130257799959e-06 | 0.000205777961181018 | 4.31835878723997 |
| **SOX17** | -1.41243873435967 | 5.49637862514277 | -4.94999386850339 | 4.26963223512162e-06 | 0.000255822131421037 | 4.07749122849891 |
| **ECT2** | 1.53522096923241 | 5.88834794509555 | 4.8686523225608 | 5.8542778342112e-06 | 0.000301929435689813 | 3.77859754344779 |

| | | | | | | |
|---|---|---|---|---|---|---|
| **ZBTB16** | -3.02969522450831 | 7.55188795965377 | -4.87651963200134 | 5.67884588081095e-06 | 0.000301929435689813 | 3.74286482301715 |
| **CDK2** | 1.11862682100119 | 5.59209412999343 | 4.84487130458789 | 6.4173250603003e-06 | 0.000301929435689813 | 3.69959253059801 |
| **NEK2** | 4.02729494171626 | 0.493595759371475 | 5.06509597756555 | 2.72070229525967e-06 | 0.00018630332859921 | 3.67232764554546 |
| **UBA7** | -1.2141914687894 | 6.41468546013424 | -4.83474309322513 | 6.67285341902151e-06 | 0.000301929435689813 | 3.63677095944523 |
| **SHISA5** | -0.66930640115519 | 8.80976332935458 | -4.84326036413137 | 6.45732012045044e-06 | 0.000301929435689813 | 3.5637039565271 |
| **DDX10** | 0.983516149902752 | 8.59542403277383 | 4.83295924997945 | 6.71887478586511e-06 | 0.000301929435689813 | 3.5379682151666 |
| **TCL1A** | -3.60769750911783 | -1.79053088752711 | -5.15108792754164 | 1.93728056791689e-06 | 0.000146621550350762 | 3.53772985043737 |
| **PRKCA** | 1.10090652009618 | 9.68726634838434 | 4.8394045725602 | 6.5540381910949e-06 | 0.000301929435689813 | 3.49767979845207 |
| **BIRC5** | 1.48648005239151 | 7.04331007026114 | 4.79554632420369 | 7.75858261634734e-06 | 0.000338086115221439 | 3.46786664095592 |
| **HLF** | -3.68052703036482 | 3.2243431277507 | -4.78759373921531 | 7.99899142414855e-06 | 0.000338310284350753 | 3.33122121520384 |
| **PTTG1** | 1.29200631473635 | 5.06175958842315 | 4.7360067155337 | 9.74432130418084e-06 | 0.000365568902310812 | 3.3161025407366 |
| **MAML3** | -1.13913797793737 | 7.79005774710433 | -4.75377675503408 | 9.10489143247562e-06 | 0.000361446903234324 | 3.28707632276831 |
| **UBE2C** | 1.85637443983706 | 5.10165107462133 | 4.72513379071813 | 1.0156909908887e-05 | 0.000365568902310812 | 3.27851506274708 |
| **BRCA1** | 1.46394547249812 | 9.65789139531925 | 4.75772508936888 | 8.96846647461913e-06 | 0.000361446903234324 | 3.19995262906164 |
| **ABL2** | 1.08776685336014 | 9.34131965458619 | 4.72482647716685 | 1.01688150851408e-05 | 0.000365568902310812 | 3.09935851674455 |
| **TCF7L1** | -1.59380066544909 | 7.94820885333099 | -4.65467696910967 | 1.32734546651691e-05 | 0.000465542141671054 | 2.92396571863269 |
| **RAC1** | 0.850385024825114 | 7.49529250357203 | 4.62902588729791 | 1.46246803422395e-05 | 0.000489076519352104 | 2.85161464493739 |
| **ANLN** | 2.21358141867246 | 6.64769665784129 | 4.60177019883416 | 1.62068121542973e-05 | 0.000529668088133625 | 2.79391433437805 |
| **CEACAM5** | -3.59123866796717 | -1.94566212971336 | -4.88258918292219 | 5.54701707898487e-06 | 0.000301929435689813 | 2.74084219158653 |
| **MAGEA3** | 6.35383561113722 | 1.58931441429007 | 4.64081873071703 | 1.39876348615455e-05 | 0.000478909974545295 | 2.56225785254754 |
| **NUF2** | 2.88900834655358 | 3.39777460745 | 4.49307712400218 | 2.43387493342437e-05 | 0.000777758256503166 | 2.39204728244211 |
| **CENPF** | 1.36054437279211 | 6.74228526996646 | 4.47769725146477 | 2.57700630663617e-05 | 0.00080559458020496 | 2.35452436067173 |
| **MEST** | -2.94502925489403 | 4.85135156753427 | -4.45261916886218 | 2.82806363577383e-05 | 0.000813985470365727 | 2.34361833347357 |
| **FOXM1** | 1.15845573899596 | 6.59134509552175 | 4.46617496489004 | 2.68955081474972e-05 | 0.000813985470365727 | 2.32284397208683 |
| **CEP55** | 1.66270193698569 | 4.14237818936096 | 4.44705713875397 | 2.88687475581725e-05 | 0.000813985470365727 | 2.30487555154276 |

| | | | | | | |
|---|---|---|---|---|---|---|
| **GJA4** | -1.16456594059534 | 6.53568928266332 | -4.44933075052264 | 2.86269249534848e-05 | 0.000813985470365727 | 2.26767532518626 |
| **RBP7** | -1.97808651177442 | 5.16052157050578 | -4.42907373910943 | 3.08524771341003e-05 | 0.000837091738091249 | 2.26177475404237 |
| **TYMS** | 1.1159138073786 | 7.98575294821864 | 4.45372554662049 | 2.81650439274219e-05 | 0.000813985470365727 | 2.21199352344049 |
| **CDX2** | -3.30856083563814 | -3.35859924892717 | -4.74822636928026 | 9.30009417223227e-06 | 0.000361446903234324 | 2.14338529729037 |
| **ORC6** | 2.14759682313583 | 3.86778865088096 | 4.38039526024092 | 3.69073968365519e-05 | 0.000931102397385292 | 2.07054924388497 |
| **TP53** | -1.50950490106517 | 9.2489821895617 | -4.42993431803163 | 3.07546107603376e-05 | 0.000837091738091249 | 2.05339893759585 |
| **BTG1** | -0.989759559639765 | 8.11983632953776 | -4.4091416280592 | 3.32054624201259e-05 | 0.000884249165928538 | 2.05020410542932 |
| **ZNF608** | -1.43373770312096 | 5.77728143507703 | -4.34086526931 | 4.26562185133743e-05 | 0.00105758003831435 | 1.9413361339008 |
| **SFRP1** | -3.9552061137872 | 4.83147470612725 | -4.31047292049684 | 4.7655806660449e-05 | 0.00112342704881518 | 1.87063651245789 |
| **FLNA** | 0.940010155872242 | 10.4533373946454 | 4.39033865825046 | 3.55838368059424e-05 | 0.00093035558776264 | 1.83775048260095 |
| **NIN** | 0.716249474555887 | 11.2502589788451 | 4.38227280191176 | 3.66538829067259e-05 | 0.000931102397385292 | 1.75575376160915 |
| **COL18A1** | -1.22817418215843 | 7.0525427017944 | -4.27986313134791 | 5.32615087510613e-05 | 0.00122739193885622 | 1.65817558683894 |
| **CDCA7L** | 1.89856705765824 | 5.32001522067576 | 4.2068988181914 | 6.93106999653896e-05 | 0.00151013312954894 | 1.51728373654644 |
| **CBL** | 0.608893897751194 | 9.43495760638122 | 4.27722664240994 | 5.37730821612949e-05 | 0.00122739193885622 | 1.51171984033877 |
| **STIL** | 1.11582966196778 | 6.82276876037446 | 4.21967263272838 | 6.61988832703788e-05 | 0.00147557628772544 | 1.46807537411121 |
| **PLEK2** | 4.30534255145317 | 0.588127128773754 | 4.31758661104576 | 4.64371469805235e-05 | 0.00111294362263321 | 1.46728695218775 |
| **SSPO** | -2.58268261592189 | 5.11161085466355 | -4.15905138396299 | 8.22689697096523e-05 | 0.00173974674180118 | 1.36907503451292 |
| **MEGF9** | -0.704205396839651 | 8.82547273090993 | -4.21758315974122 | 6.66985109194394e-05 | 0.00147557628772544 | 1.3501561337423 |
| **DTX1** | -2.51873105530207 | 3.78425152972448 | -4.13701300461795 | 8.89949608753857e-05 | 0.00177742713526117 | 1.2924880789786 |
| **CNTNAP2** | -4.92483878148736 | -1.22668120237324 | -4.32227941445264 | 4.56497632011444e-05 | 0.00111261626242789 | 1.28549499345275 |
| **EMCN** | -1.85684173124978 | 3.47133801066378 | -4.14030123234929 | 8.79588267720773e-05 | 0.00177742713526117 | 1.28451873012508 |
| **CDCA8** | 1.53216418124137 | 6.34857896475686 | 4.14435574541503 | 8.66972204510507e-05 | 0.00177742713526117 | 1.24802658906493 |
| **MELK** | 2.42940078477617 | 4.39987719884064 | 4.10981159011044 | 9.80288270538686e-05 | 0.00188346292284841 | 1.22301263834308 |

| | | | | | | |
|---|---|---|---|---|---|---|
| **ARG2** | 2.47698271081156 | 2.79907484131873 | 4.131480153422 | 9.07649654614423e-05 | 0.00178794548402129 | 1.19787712568063 |
| **TGFB1** | 0.818767204649225 | 9.69031825923995 | 4.1828122426857 | 7.55664547853159e-05 | 0.00162185913404902 | 1.17269919860625 |
| **CX3CL1** | -1.57380694808997 | 4.37737454936791 | -4.07010719426765 | 0.000112815430602868 | 0.00207985370778107 | 1.09766532731182 |
| **SPRY4** | -1.42264084573196 | 7.76366624910483 | -4.10922379871957 | 9.82334625964053e-05 | 0.00188346292284841 | 1.04918776452387 |
| **PRMT5** | 0.721423699617493 | 10.2386843571671 | 4.14937640950808 | 8.51591254083612e-05 | 0.00177476554111918 | 1.0234595362917 |
| **ABI1** | 0.589889341546361 | 7.65939442898955 | 4.09042601714651 | 0.00010499891630219 | 0.00196088885250063 | 0.99219842811368 |
| **CEBPA** | -2.0184382685675 | 3.8665713809092 | -4.01689367402131 | 0.000136027856713635 | 0.00238546412139277 | 0.9239105179441 |
| **IFI16** | 0.841496339137113 | 7.17012536446893 | 4.05138611411388 | 0.000120510274846529 | 0.00219359209151023 | 0.889221094311413 |
| **TFG** | 0.743943991578584 | 9.92361777276792 | 4.0967653502981 | 0.000102668737174537 | 0.00194260057969717 | 0.867452448392681 |
| **CD36** | -1.90046998940801 | 7.07763823172717 | -4.03816844804607 | 0.000126244264584566 | 0.00226924065590757 | 0.851145349718682 |
| **BUB1B** | 2.31538481278129 | 4.34223097799566 | 3.95010436355237 | 0.000171699720503224 | 0.00280589688332397 | 0.724039745585332 |
| **EDNRB** | -1.40448357215831 | 5.80590005369478 | -3.96823601950368 | 0.000161215363296115 | 0.00275985348118825 | 0.715543200434919 |
| **SDHB** | 0.797064893128686 | 9.02510705733615 | 4.02959038615312 | 0.000130104840409372 | 0.00230976247541577 | 0.707065540609236 |
| **CD79B** | -1.54640903120178 | 3.04660188769957 | -3.95008717616175 | 0.000171709962261829 | 0.00280589688332397 | 0.679798779632503 |
| **HMGA2** | -3.56393816851271 | 6.73720236397679 | -3.95709128503346 | 0.000167584457376815 | 0.00280216801985883 | 0.609489021568163 |
| **BAG1** | 0.942190042594286 | 7.03604331038156 | 3.9628461115673 | 0.000164266090229685 | 0.00277899573823868 | 0.608663076206334 |
| **PTPRB** | -1.6580993658271 | 4.47299925907608 | -3.91048149081594 | 0.00019693310236014 | 0.00300885351606468 | 0.600441846968578 |
| **FGFR1OP** | 0.829094615908327 | 8.79237545295724 | 3.99140464992971 | 0.000148709670231858 | 0.00257643982883628 | 0.598350163638586 |
| **ODC1** | 1.13485461567255 | 7.21899801246465 | 3.93327949915497 | 0.000182010826669937 | 0.00290812854168189 | 0.502555085591766 |
| **RRAGC** | 1.01787562533637 | 6.68984637110609 | 3.9077803995404 | 0.00019877683172889 | 0.00300885351606468 | 0.454554995704194 |
| **PBK** | 2.34533142498986 | 3.3217000496932 | 3.86687345572315 | 0.000228802722350053 | 0.00329018314739377 | 0.451150116292498 |
| **ANXA1** | 0.934227085358573 | 8.69789629853966 | 3.9425698229985 | 0.000176246060577564 | 0.00284766106865772 | 0.445846190931237 |
| **COL15A1** | -1.56573358078258 | 8.22882430859266 | -3.92218814415196 | 0.000189129938392268 | 0.00292706231996158 | 0.410911737049359 |
| **FAP** | 2.63757428823055 | 5.44389281084811 | 3.85772117773592 | 0.000236091243608216 | 0.0033613783000853 | 0.392721213238443 |

| | | | | | | |
|---|---|---|---|---|---|---|
| **SOCS1** | -1.20957620446082 | 7.3906536326869 | -3.89380131730105 | 0.000208585987525586 | 0.00309223350579168 | 0.366186653154318 |
| **CYLD** | 0.824769246507302 | 9.36599616675823 | 3.92687832284302 | 0.000186087527439602 | 0.00292706231996158 | 0.345988226481082 |
| **SDHC** | 0.879171414761639 | 7.92909864628879 | 3.8863153161425 | 0.000214027784985974 | 0.00310880762434172 | 0.31284134022158 |
| **P2RY8** | -1.41270844848827 | 3.61080421098569 | -3.81404134461128 | 0.000274043597732109 | 0.00375309231941689 | 0.306241929174495 |
| **DDIT4** | -1.26848959112528 | 8.49303839063792 | -3.88704120884144 | 0.000213494219336489 | 0.00310880762434172 | 0.28073239966534 |
| **MAGEA6** | 4.90830035040555 | 0.319771394873463 | 3.89469406506388 | 0.000207945933683727 | 0.00309223350579168 | 0.27579782194208 |
| **RHOA** | 0.522956699073175 | 10.5294436249095 | 3.92192450623122 | 0.000189302361443969 | 0.00292706231996158 | 0.249198930504259 |
| **UBE2T** | 1.26428081139244 | 4.7759738516116 | 3.78814341441324 | 0.000299228189539234 | 0.00402140314539643 | 0.216911265592535 |
| **CCNA2** | 1.52766779571648 | 4.61642658112547 | 3.75626526938839 | 0.000333268544247932 | 0.0043966987764085 | 0.127399739315665 |
| **APH1A** | 0.588710029913238 | 8.25216939630856 | 3.81702042734559 | 0.000271279990673127 | 0.00375309231941689 | 0.072866709925294 |
| **NFIC** | -0.626949285785596 | 9.85583721746982 | -3.85083604992552 | 0.000241720122672831 | 0.00340777976866207 | 0.0655100267123885 |
| **LRRC15** | 3.21449572673313 | 6.36894823259401 | 3.73864330473226 | 0.00035363988712877 | 0.00456001840809545 | -0.0522317684755 |
| **SNW1** | 0.71607516063691 | 6.51772116224886 | 3.73736629715405 | 0.000355161378099228 | 0.00456001840809545 | -0.0684484794273992 |
| **ATIC** | 0.79052172524664 | 9.40988703891627 | 3.79416868333644 | 0.00029317880595044 | 0.00397727474487483 | -0.0834913309982079 |
| **NOTCH4** | -1.04605242677878 | 10.5180359050849 | -3.81669783340554 | 0.000271577960971047 | 0.00375309231941689 | -0.0897737843431097 |
| **PAFAH1B2** | 0.711371477125049 | 7.83157130647687 | 3.73820483328403 | 0.000354161602150683 | 0.00456001840809545 | -0.150237287608659 |
| **BCL2** | -1.03412182275148 | 10.8232205867547 | -3.78267003737772 | 0.000304826614061652 | 0.00405870991685792 | -0.22016032170892 |
| **KMT5A** | 0.715843869676683 | 6.97457438442478 | 3.6829398742777 | 0.000426130010108615 | 0.00542278720828485 | -0.27047189982518 |
| **PVRIG** | -1.06581121150692 | 5.8573433177247 | -3.65101915954171 | 0.000473831409111564 | 0.00597692602019674 | -0.274744759251115 |
| **TDRD7** | 2.04223885251096 | 3.07426554457914 | 3.59499331725836 | 0.000570067289524227 | 0.00694709120623592 | -0.33715340729512 |
| **PPARG** | -1.56487204255917 | 6.69288535824364 | -3.6208406565452 | 0.000523565456897678 | 0.00643493270956292 | -0.43946972555853 |
| **ARID3A** | -1.0374864730222 | 4.48967375537047 | -3.56019887924892 | 0.000638884514459888 | 0.00759269365118446 | -0.444924244890665 |

| | | | | | | |
|---|---|---|---|---|---|---|
| **PAGE5** | 4.92537854161681 | -0.914344196772872 | 3.64155724777841 | 0.000488919748460731 | 0.00611362259379592 | -0.463954143497072 |
| **WBSCR17** | -3.68399822855932 | 3.55138461170312 | -3.54085551347122 | 0.000680477110732919 | 0.00795549662791819 | -0.479387324630422 |
| **MCM4** | 1.24684455842201 | 5.48081375041509 | 3.55459815533236 | 0.000650672590841826 | 0.00766940316090612 | -0.529371328771038 |
| **CHEK1** | 1.82706636049403 | 4.41465354627502 | 3.52171978930035 | 0.000724138960835521 | 0.00833049460545183 | -0.551986866592586 |
| **BUB1** | 1.1022246454636 | 4.83284892492767 | 3.50494524351866 | 0.000764583249664654 | 0.0086562558298236 | -0.624278229047891 |
| **LAMB1** | -0.950671596006534 | 8.1525155608191 | -3.58062573387698 | 0.000597584761892011 | 0.00717167172791234 | -0.654407999578964 |
| **ATM** | 0.703019238920122 | 10.6486538320452 | 3.62750603926713 | 0.000512171669556789 | 0.00634916259329882 | -0.694160003377232 |
| **SDHD** | 0.831059231930197 | 6.21377881179977 | 3.50333042631485 | 0.000768587513037279 | 0.0086562558298236 | -0.749252578543153 |
| **WT1** | -3.79259073372004 | 2.77319785943346 | -3.43199868106804 | 0.000966641980764635 | 0.0103733669279071 | -0.790456265594206 |
| **EXO1** | 2.00279045347618 | 3.93328904255895 | 3.42652197963517 | 0.00098369123167372 | 0.0104252050750226 | -0.802191389845009 |
| **CTSV** | 3.01706855863633 | -1.26588062403566 | 3.50255590772806 | 0.000770515122543408 | 0.0086562558298236 | -0.827127693226366 |
| **PSMB8** | 0.957522222412327 | 6.55759138665133 | 3.46973382516442 | 0.000856537857654673 | 0.00940230106341541 | -0.879560518427239 |
| **RBX1** | 0.668416650745774 | 6.93118955778652 | 3.47814215303098 | 0.000833674066573849 | 0.00926049963678907 | -0.884886176439246 |
| **EPS15** | 0.638653119364228 | 11.359797274409 | 3.58017376290258 | 0.000598470519714521 | 0.00717167172791234 | -0.89385111361774 |
| **TPM1** | 0.984038645729397 | 9.88373094770697 | 3.53222966518331 | 0.000699840544464348 | 0.008115892765643 | -0.930600283773695 |
| **RAD51** | 1.48821284725954 | 3.97773722597828 | 3.3784427348525 | 0.00114603452456597 | 0.0117974911662533 | -0.935987908699992 |
| **CREB1** | 0.444782461515555 | 7.7000475296914 | 3.47683874289277 | 0.000837180078430166 | 0.00926049963678907 | -0.93807438849456 |
| **STEAP1** | 2.68974886741476 | 3.42838428831222 | 3.36832267888667 | 0.001183280005808 | 0.0119827932982528 | -0.952494274094819 |
| **ITPKB** | -0.802228956353141 | 6.44507019634005 | -3.4257961040893 | 0.000985972107234407 | 0.0104252050750226 | -0.998092043761567 |
| **FZD7** | -1.18702904209053 | 7.4705948003603 | -3.44853417807802 | 0.000916843182958361 | 0.00991293606837686 | -1.00782836221529 |

| | | | | | | |
|---|---|---|---|---|---|---|
| **CDKN1A** | -0.934945504323371 | 8.32124008617721 | -3.46366061469204 | 0.000873419375451717 | 0.0095149777416634 | -1.01638100639175 |
| **FOXO3** | -0.736842174992096 | 6.44339480572688 | -3.41326150747668 | 0.00102615483724575 | 0.0107708807004335 | -1.03435341167893 |
| **MRE11A** | 0.940064811067709 | 4.83310862762722 | 3.34371012978278 | 0.00127867673212883 | 0.012594090005488 | -1.08108591401449 |
| **ATAD2** | 1.3039039217806 | 4.00626755112027 | 3.30456884525532 | 0.00144542412661026 | 0.0134098057681649 | -1.13811953581787 |
| **FANCD2** | 1.7801577370913 | 2.92143842026188 | 3.29843804237616 | 0.00147332389743861 | 0.0135307841291667 | -1.1387554121179 |
| **CEACAM8** | -2.27134927482478 | 1.48590001137227 | -3.3128165477166 | 0.00140867342163965 | 0.0133267919757751 | -1.15161955519393 |
| **MOCOS** | 1.94536880612763 | 3.42585577046377 | 3.27525453679991 | 0.00158346560321777 | 0.0140184791307432 | -1.20020557113246 |
| **FOXO1** | -1.11347908851806 | 6.64283308267774 | -3.36122910747456 | 0.00121006343550875 | 0.0120838279184832 | -1.20236225887212 |
| **SPARCL1** | -1.22518051943253 | 5.55364086163782 | -3.313995711957 | 0.00140349154491279 | 0.0133267919757751 | -1.22843962661701 |
| **COL4A2** | -0.866119440104926 | 8.52492035720122 | -3.3932915902232 | 0.00109337991072496 | 0.0113933355914674 | -1.23827710703531 |
| **NOS1AP** | -2.19990250795027 | 1.9530615040011 | -3.25276805288032 | 0.00169764049786503 | 0.0142760645376018 | -1.28094542856102 |
| **MAGEA12** | 4.19685564643552 | 0.032061366504364 | 3.2771058123032 | 0.00157439414130236 | 0.0140184791307432 | -1.29874992263145 |
| **ACVRL1** | -1.07168675302581 | 8.78779244185615 | -3.37512575350301 | 0.00115811878802923 | 0.011811168916213 | -1.3119177014528 |
| **PAK2** | 0.559817040112623 | 6.84128892990554 | 3.32222195768121 | 0.00136783650053966 | 0.0131569352537161 | -1.330660594277 |
| **COL4A1** | -0.807975072661858 | 9.27987636861398 | -3.37774307827808 | 0.00114857354887028 | 0.0117974911662533 | -1.34407831321242 |
| **CDC45** | 2.1871105422263 | 5.60932646360077 | 3.2682264619683 | 0.00161835251300352 | 0.0140192223716811 | -1.36188662322779 |
| **LMO4** | 0.873246160661167 | 7.50042981347622 | 3.32703505301055 | 0.0013473718484602 | 0.0130913562032822 | -1.36274586563694 |
| **IRF4** | -1.58061895387024 | 5.76755311380368 | -3.27328973498627 | 0.00159314700446633 | 0.0140184791307432 | -1.36419787057604 |
| **BRCA2** | 0.937117212439533 | 9.28425829436263 | 3.36541543757023 | 0.0011941887338027 | 0.0120086950993587 | -1.38040708026672 |
| **BLVRA** | 0.638914347582419 | 6.05783765375503 | 3.27043350576998 | 0.00160731995593755 | 0.0140184791307432 | -1.40308321109276 |
| **DLL4** | -0.873153667266717 | 7.37247977650118 | -3.3080905604161 | 0.00142962294122794 | 0.0134098057681649 | -1.40883290549627 |

| | | | | | | |
|---|---|---|---|---|---|---|
| **RGCC** | -0.911504431991517 | 5.96359606347053 | -3.26317931382896 | 0.00164384976414327 | 0.0140206362467929 | -1.41324451125457 |
| **EZH2** | 0.985660158493691 | 6.95285864037927 | 3.29553891805329 | 0.00148669255383055 | 0.0135307841291667 | -1.41539933602414 |
| **SOX11** | -2.20510768821388 | 5.11979055678222 | -3.22622991216077 | 0.00184231087673395 | 0.0153135435881123 | -1.42961184682809 |
| **DCAF12** | 0.810155336626987 | 6.48046987078603 | 3.27019312824774 | 0.00160851812000878 | 0.0140184791307432 | -1.44659737108155 |
| **TNFRSF1A** | -0.486813209341081 | 9.91110533381927 | -3.3578370685507 | 0.00122307167915091 | 0.012129497066338 | -1.45077855123054 |
| **LDHB** | 0.922866420522018 | 9.16110216127409 | 3.33663863507401 | 0.00130739617700697 | 0.0127893585206533 | -1.4540517566005 |
| **XPO1** | 0.675890131985466 | 7.60993875438212 | 3.29705634511225 | 0.00147968113475795 | 0.0135307841291667 | -1.45528107545087 |
| **TTL** | 0.530141875853412 | 8.6450216623707 | 3.32115319400616 | 0.0013724202281345 | 0.0131569352537161 | -1.45683210559198 |
| **ADGRB3** | -2.97972534272869 | 1.69824989142076 | -3.17772970545985 | 0.00213697429997517 | 0.0170720502409127 | -1.47401980737066 |
| **CCNE1** | 1.08191425006901 | 7.70209747114747 | 3.27078422279867 | 0.00160557330581068 | 0.0140184791307432 | -1.53570271486108 |
| **SH3PXD2A** | -0.604242357373304 | 9.2055584514404 | -3.30622764499862 | 0.00143796111074073 | 0.0134098057681649 | -1.54542020946693 |
| **KIAA0040** | -2.65256111995845 | -0.153657894180022 | -3.17003652148105 | 0.00218757876746546 | 0.0173797694343389 | -1.5556042793872 |
| **PDGFB** | -0.674981390629425 | 8.45789187372517 | -3.27826324456384 | 0.00156874739673047 | 0.0140184791307432 | -1.56516114286699 |
| **AURKB** | 1.12715826230456 | 5.53036410678676 | 3.18848455230011 | 0.00206806275058316 | 0.0168015493521954 | -1.57314082083674 |
| **CDC25C** | 1.21693568355828 | 5.39249018650722 | 3.1829250530207 | 0.00210342144964365 | 0.0169806013688017 | -1.57404032449007 |
| **EPOR** | -0.837605277896282 | 6.64882776081772 | -3.2288676644236 | 0.00182743048962036 | 0.0152781688608958 | -1.57775324448136 |
| **FEV** | -2.79944509062023 | -2.9604208005247 | -3.2093708785957 | 0.00194013133460969 | 0.0158912307969956 | -1.59530505810076 |
| **MAPK1** | 0.565417252451175 | 7.23318050063985 | 3.21939101161283 | 0.00188142153653423 | 0.0155487595950357 | -1.65007900250511 |
| **APLN** | -1.14232535993445 | 4.97828342163412 | -3.13724196732109 | 0.00241611483496183 | 0.0184807081525272 | -1.65629578082064 |
| **ROBO4** | -0.859930886937599 | 9.16384971857725 | -3.26241061195986 | 0.00164776601231432 | 0.0140206362467929 | -1.66736449169246 |

| | | | | | | |
|---|---|---|---|---|---|---|
| **MCM6** | 1.40253383882538 | 4.51219909480524 | 3.11500426372814 | 0.00258355559609948 | 0.0194510625507385 | -1.67373005483544 |
| **CITED4** | -1.65314875449 02 | 2.1799695501392 | -3.08793224196676 | 0.00280198916480821 | 0.0203498000959303 | -1.6780699189582 |
| **CRNDE** | 1.10805866396247 | 4.71157592530515 | 3.10530505731138 | 0.00265992249032207 | 0.0197163326859956 | -1.71618277403781 |
| **BCL6** | -0.832158057835464 | 9.96889971039958 | -3.26447332393079 | 0.00163727698142662 | 0.0140206362467929 | -1.72455536995364 |
| **CA4** | -2.95643238483437 | -1.31046850071621 | -3.10944830945414 | 0.00262704670765479 | 0.0196486226375845 | -1.73628565240617 |
| **INSR** | -0.672325765254801 | 9.99306056756542 | -3.25997217057258 | 0.00166024695094334 | 0.0140437359732736 | -1.73921133818343 |
| **ITGAV** | 0.830757054205023 | 7.08415699795047 | 3.18132241161846 | 0.00211371880738213 | 0.0169806013688017 | -1.74541721714404 |
| **PSENEN** | 0.81094634189665 | 6.75695886193859 | 3.16730996830786 | 0.00220578075894112 | 0.0174280919305348 | -1.75778208627149 |
| **CTPS1** | 0.967480331997654 | 6.54881006076923 | 3.16012652254246 | 0.00225441595751903 | 0.0177150281252042 | -1.75866924485791 |
| **TNS1** | -0.825616094556832 | 6.46834973659149 | -3.14467391916101 | 0.00236245137486292 | 0.0182645434250155 | -1.79326137034542 |
| **SESN1** | -1.28370388849483 | 4.79899924115412 | -3.07785550002394 | 0.00288759730144036 | 0.0208661553742273 | -1.79614900185372 |
| **ECSCR** | -0.800579642368319 | 5.50942648433826 | -3.09689535464351 | 0.00272783357521222 | 0.0200133912303835 | -1.81768690977435 |
| **KRAS** | 0.795871348017331 | 7.31193498919579 | 3.15638917453553 | 0.00228011387016577 | 0.0177761902267082 | -1.83023527951455 |
| **CD79A** | -1.93194203747698 | 1.76662188534424 | -3.00839523161013 | 0.00354720749319575 | 0.0244061453359593 | -1.87442765109203 |
| **AFF3** | -1.60663034578257 | 6.35209578092677 | -3.10817319490858 | 0.00263712390059375 | 0.0196486226375845 | -1.8805533108497 |
| **IL7** | 2.29444439348232 | 2.36493528308722 | 2.99637396115076 | 0.0036746610767293 | 0.0246923487305455 | -1.89480546526656 |
| **PALB2** | 0.633087008786023 | 7.62713683196386 | 3.13971369259775 | 0.00239814241554693 | 0.0184413304468261 | -1.8966943662804 |
| **ITGB1** | 0.756513307459482 | 8.46280016407432 | 3.15540549757083 | 0.00228692294293534 | 0.0177761902267082 | -1.91048779367398 |
| **MAPKAPK2** | 0.640711311829005 | 7.63402349110721 | 3.12811720682324 | 0.00248355690506066 | 0.0188960572988213 | -1.92894232640239 |
| **RAD51B** | 0.859440929960964 | 4.29689711874123 | 3.00617446721735 | 0.0035704396292169 | 0.0244490104133996 | -1.93724752856511 |

| | | | | | | |
|---|---|---|---|---|---|---|
| **NDC80** | 1.67809796089948 | 3.6084823352177 | 2.98488441799906 | 0.00380043240373208 | 0.025184432242243 | -1.94759557926142 |
| **CDC6** | 1.44395578597339 | 6.26393514089737 | 3.06883501360638 | 0.00296629034285234 | 0.0212215199652819 | -1.97702146451408 |
| **WNT7B** | 3.51872473119136 | 1.06533289740239 | 2.96850806365224 | 0.00398660084164382 | 0.0259308049086591 | -1.98150106925754 |
| **PRKACG** | -1.38754134721895 | -0.103805657040987 | -2.97187224048224 | 0.00394768096927666 | 0.0258034783355447 | -2.0000688059311 |
| **SRD5A1** | 1.21104207822223 | 4.4828535134421 | 2.98613646986296 | 0.00378653558486373 | 0.025184432242243 | -2.00344853084469 |
| **PICALM** | 0.576275367730164 | 11.7354884362364 | 3.20855895743843 | 0.00194496287918722 | 0.0158912307969956 | -2.02141732635754 |
| **NFE2L2** | 0.977873728406751 | 5.91553680282495 | 3.03605588755402 | 0.00326935260752163 | 0.0228219856777481 | -2.0247303934294 |
| **SRSF3** | 0.455402191076088 | 6.7750176509881 | 3.06955122539668 | 0.00295997007158016 | 0.0212215199652819 | -2.02516054187619 |
| **IRF7** | -0.708019595551023 | 6.08992988593943 | -3.04000351338882 | 0.00323138827660033 | 0.0227781193223102 | -2.03438168650463 |
| **PLAG1** | -2.39739410693176 | 2.96205707363763 | -2.91873516062149 | 0.00460548853638632 | 0.0286696645685001 | -2.0847656243488 |
| **ISG20** | -0.912286244533581 | 4.87652462090335 | -2.96708028552146 | 0.00400322579257463 | 0.0259308049086591 | -2.0895756566244 |
| **KDM1A** | 0.605825995925193 | 6.54768124845609 | 3.03699644987733 | 0.00326026980927273 | 0.0228219856777481 | -2.09111541341507 |
| **PDIA3** | 0.599478913743603 | 8.53114464026027 | 3.08965110097006 | 0.00278762445317281 | 0.020348243470368 | -2.09633053833146 |
| **AKT3** | 1.11969994445742 | 10.1067870226873 | 3.11728933981082 | 0.0025658639350 58 | 0.0194195386242811 | -2.14808420311452 |
| **WNT2** | 2.85094535033056 | 4.04999333239555 | 2.90681797919946 | 0.0047662840753316 | 0.0294159506451795 | -2.16648263587176 |
| **HDAC1** | 0.814563115246188 | 4.68008610313991 | 2.9194094517224 | 0.00459654193089168 | 0.0286696645685001 | -2.19007583469095 |
| **NUP98** | 0.411595762735956 | 10.0567816697456 | 3.10047346919577 | 0.00269874409681184 | 0.0199015077498227 | -2.19059848012473 |
| **ZNF703** | -0.751054468721194 | 6.61571275872583 | -3.00013405842977 | 0.00363434608459461 | 0.0246518380643729 | -2.19527730084442 |
| **TMPRSS2** | -2.06015261843754 | 1.2339137065014 | -2.86749835536325 | 0.00533426566551155 | 0.0320953259672848 | -2.20349169788029 |
| **NRAP** | 4.27983013340913 | -0.0558643808349152 | 2.86390255962215 | 0.00538920288233745 | 0.0321563225925363 | -2.23310119699666 |

| | | | | | | |
|---|---|---|---|---|---|---|
| **BRIP1** | 0.924397035123963 | 7.80002928805852 | 3.0144820423069 | 0.00348424822923413 | 0.0240882161232629 | -2.24669264107619 |
| **FMOD** | -1.78831753566505 | 7.90805435829958 | -3.01705806285318 | 0.00345791654681393 | 0.0240216618083016 | -2.24696955765673 |
| **GINS2** | 0.947225289582278 | 4.99693762386322 | 2.89561123750788 | 0.00492220426777224 | 0.030119701008751 | -2.28271303776919 |
| **MAGEC1** | 3.39120296711718 | 1.59515937081083 | 2.82754536512314 | 0.00597466621303905 | 0.0346434274772184 | -2.28712456559328 |
| **RAD54L** | 1.22780611130244 | 4.40009483515272 | 2.86445452289851 | 0.00538073618769135 | 0.0321563225925363 | -2.29915467611268 |
| **PTGDS** | -2.1606771406803 | 4.20390008415428 | -2.8573972771429 | 0.00548991710365446 | 0.032621904111798 | -2.29920544053497 |
| **DTL** | 0.984866420721693 | 3.96624427922039 | 2.84927614643137 | 0.00561807780060558 | 0.0331098191691427 | -2.30020700982949 |
| **CDHR1** | -2.60325511798437 | 0.689900263800672 | -2.817693220843 | 0.00614308597265482 | 0.0354769382677817 | -2.32019968169261 |
| **SMAD4** | 0.445674929288453 | 9.95080416962075 | 3.04765212644612 | 0.00315899429644027 | 0.0224882861301045 | -2.32662976436769 |
| **DIRC2** | 0.489107801368794 | 8.06330198086246 | 2.98998510740476 | 0.00374411287620496 | 0.0250420200743383 | -2.32943602692619 |
| **MEG3** | -2.36789385182138 | 5.46196644639608 | -2.87602905491847 | 0.00520598845750329 | 0.0317212347537701 | -2.3840678947477 |
| **CEACAM1** | -1.15384856661421 | 3.02377342420284 | -2.79058444747358 | 0.00662924891620382 | 0.037828809291671 | -2.38626417119892 |
| **MCAM** | -0.7395210477849 | 7.82797755800885 | -2.95456934730061 | 0.00415167087851978 | 0.0267717610910827 | -2.40685996444145 |
| **NUP214** | 0.383373071054937 | 10.9773563156576 | 3.04234059018167 | 0.00320910600184426 | 0.0227324848800593 | -2.41997894138887 |
| **TBC1D10C** | -0.838350415562464 | 5.37647127609931 | -2.85054210397207 | 0.00559792015598104 | 0.0331098191691427 | -2.43782877334046 |
| **PSME1** | 0.665212777305008 | 7.63706827543938 | 2.93286547590958 | 0.00442132694111026 | 0.0280082296974298 | -2.45086679065044 |
| **PIK3R1** | -0.868696595101166 | 10.0002309925012 | -3.00214901721743 | 0.00361291123384085 | 0.0246225893566973 | -2.45298377002289 |
| **GMPS** | 0.736134310014126 | 6.50977090872864 | 2.89668685474936 | 0.00490703757161215 | 0.030119701008751 | -2.45363256857338 |
| **MAFB** | -0.779270775803513 | 7.96666758464597 | -2.93338814337186 | 0.00441464728623057 | 0.0280082296974298 | -2.47158252787519 |
| **KIAA0125** | -1.6140311222143 | 2.57546567298117 | -2.74115463483845 | 0.00760746492464152 | 0.0420751329293635 | -2.48418853926849 |

| | | | | | | |
|---|---|---|---|---|---|---|
| **SYCP3** | 2.21650916736716 | -2.17688816400266 | 2.76381681031222 | 0.00714366444167336 | 0.0402846645769658 | -2.48844286713558 |
| **JUN** | -0.719260578422794 | 9.62709941709403 | -2.97634238756601 | 0.00389651036658622 | 0.0257026692988578 | -2.49259168959055 |
| **PLEKHA4** | -0.990449097354285 | 6.35034880804293 | -2.86749337613911 | 0.00533434138121076 | 0.0320953259672848 | -2.5109902613894 |
| **BCAM** | -0.826723536264134 | 5.93073279639023 | -2.84151783685354 | 0.00574307735083536 | 0.0335713220752083 | -2.5270968158759 |
| **RAD51AP1** | 1.15441516852903 | 3.90924645151829 | 2.75053228463808 | 0.00741230285399743 | 0.0411540212511517 | -2.53023148360366 |
| **EGF** | 2.8170765764054 | 1.08348688686692 | 2.71540025860152 | 0.00816785096900644 | 0.0441555251632754 | -2.53323988818836 |
| **NBEA** | -1.90387614394964 | 3.15268652937989 | -2.72380293707688 | 0.00798100779886586 | 0.0438041573082791 | -2.54422364970159 |
| **TTK** | 2.04804269121851 | 4.32760949155221 | 2.75858950874599 | 0.00724828906790212 | 0.0404450416985391 | -2.54743791647965 |
| **SKP1** | 0.748586285593642 | 6.91753203598304 | 2.87146807774748 | 0.00527421466068916 | 0.0320013530889072 | -2.55653167000825 |
| **MALT1** | 0.69962018765289 | 8.73613268595508 | 2.9129802421296 | 0.00468250244055525 | 0.0290234418513726 | -2.58384165122969 |
| **BMF** | -0.701515239147074 | 6.74675352412345 | -2.84746359920386 | 0.00564705494351605 | 0.0331447551378616 | -2.60210765247357 |
| **NRG2** | -2.05222108901016 | 3.05812907595956 | -2.69483692496611 | 0.00864205275860999 | 0.0453550068134349 | -2.60489733433097 |
| **PARP12** | 0.873851382465765 | 4.23223254928642 | 2.7212232563922 | 0.00803795066799996 | 0.0438896505371659 | -2.62643991483794 |
| **ELN** | -2.03587957061992 | 11.6220344356367 | -2.9723951808889 | 0.00394166280208499 | 0.0258034783355447 | -2.66144167345016 |
| **IL11** | 2.92968373994671 | 2.25371806620626 | 2.65485324672041 | 0.00963657677648872 | 0.0485759779235897 | -2.66601211874693 |
| **WIF1** | -3.39894651480548 | -2.28157580928531 | -2.67382472847345 | 0.00915239678422535 | 0.0470040949132716 | -2.66623775347832 |
| **NONO** | 0.349310497353134 | 12.5888330264345 | 2.99831309862434 | 0.00365381856026692 | 0.0246675638012386 | -2.67242434648019 |
| **NKD1** | -1.7404837879964 | 4.52967115004595 | -2.7011839116723 | 0.00849307683365252 | 0.0450665848221119 | -2.7032460343468 |
| **NUTM2A** | -1.26646370021783 | 5.70234210510509 | -2.75818310252351 | 0.00725648175119825 | 0.0404450416985391 | -2.70485622805485 |
| **PDCD1** | -1.28679306561792 | 4.39876905994992 | -2.6867588352693 | 0.00883510402476273 | 0.0458504442959954 | -2.72311243931604 |

| | | | | | | |
|---|---|---|---|---|---|---|
| **SMAD9** | -1.21095834337549 | 6.35871847764882 | -2.77920496061968 | 0.00684364200672664 | 0.0387447134081611 | -2.73309514377699 |
| **HMMR** | 2.08041515857325 | 4.8863526961286 | 2.70517440392545 | 0.00840061492682403 | 0.0450665848221119 | -2.73396219253175 |
| **AXL** | 1.15830071584403 | 12.0164245895428 | 2.95134272430674 | 0.00419077292066712 | 0.0269032654460684 | -2.75112461310955 |
| **C1RL** | -0.668668556222687 | 5.03280036865967 | -2.68951687049056 | 0.00876875489368377 | 0.045686483830135 | -2.7881759582603 |
| **S1PR2** | -0.80202928677725 | 7.72345621188695 | -2.79815734565119 | 0.00649000374418024 | 0.0373305015365247 | -2.80111281632265 |
| **CENPA** | 1.1983479035785 | 4.57197321520107 | 2.65634748848119 | 0.00959761330767465 | 0.0485759779235897 | -2.81225729508276 |
| **DTX3L** | 0.518639590451021 | 5.80507355699621 | 2.71896294780935 | 0.00808814839523572 | 0.0438896505371659 | -2.81290144191127 |
| **HDAC8** | 0.727329017266337 | 5.5359016033961 | 2.70187285319281 | 0.00847704771003686 | 0.0450665848221119 | -2.82002958854072 |
| **PLVAP** | -0.764219459198063 | 6.89985855397016 | -2.76152368321804 | 0.00718939012757177 | 0.0403841523572196 | -2.83113534417644 |
| **LY6E** | 0.902357905661092 | 8.03043097194872 | 2.79239423167253 | 0.00659572505122119 | 0.0377874606520162 | -2.83671866852886 |
| **DEK** | 0.61679671298729 | 12.7601135410959 | 2.93753096099066 | 0.00436203129173355 | 0.0278782266556127 | -2.84816426795701 |
| **LMNA** | 0.520811949613471 | 12.4731661659254 | 2.92889735318889 | 0.00447234465072954 | 0.028207156174338 | -2.84832983890064 |
| **KIF5B** | 0.697788734554228 | 12.3779587087822 | 2.92406160793102 | 0.00453525229726307 | 0.0284790078753899 | -2.85340054766333 |
| **CCL14** | -1.23084133789336 | 5.12485143613191 | -2.66447740492703 | 0.00938811778159096 | 0.0477035808124657 | -2.85811514035724 |
| **SCUBE2** | -1.28908116816095 | 5.43767498852039 | -2.67457339577602 | 0.00913375182511437 | 0.0470040949132716 | -2.87272731812452 |
| **GMNN** | 0.79600058064605 | 6.12346557054396 | 2.69756459302717 | 0.00857773976517239 | 0.0452599262803017 | -2.90459375265552 |
| **TET1** | -1.10299522432242 | 7.35462248858777 | -2.73847670868687 | 0.00766405210252155 | 0.0422256970246206 | -2.92384183322712 |
| **CD47** | 0.585399966068079 | 6.24075448525865 | 2.69287598611449 | 0.00868855896077328 | 0.0454332646748799 | -2.930193423975 |
| **CD248** | -0.904725332642988 | 10.5896123146487 | -2.83518991460079 | 0.0058469186580532 | 0.0340399555881802 | -2.93583186319763 |
| **RAD51D** | 0.480736194773715 | 7.02183243654285 | 2.7193350626358 | 0.00807986471362932 | 0.0438896505371659 | -2.9451656240566 |
| **HDAC4** | -0.650709438624883 | 6.16126587912274 | -2.66882947653576 | 0.0092776885931664 | 0.0473096319041606 | -2.97814403353734 |

| | | | | | | |
|---|---|---|---|---|---|---|
| **DDX21** | 0.692469633831638 | 5.70929447492488 | 2.64253282646538 | 0.0099633612461387 | 0.0495627165466753 | -2.98285513085174 |
| **ALDH2** | -0.796214067895345 | 6.94313203612925 | -2.66993988936125 | 0.00924970290379556 | 0.0473096319041606 | -3.05822834107074 |
| **TBL1XR1** | 0.488949287819501 | 10.9883791694911 | 2.78656999140017 | 0.00670416959369212 | 0.0381051220384556 | -3.09149998597109 |
| **CCND3** | -0.498783360667474 | 8.57935854761159 | -2.70393022119597 | 0.00842934437688724 | 0.0450665848221119 | -3.09844837875919 |
| **HIST1H3B** | 0.859541168739522 | 7.37606746146405 | 2.653913795907 | 0.00966114720872508 | 0.0485759779235897 | -3.13085359212759 |
| **CCL2** | 1.03732961733387 | 7.22184740386323 | 2.64134982229085 | 0.00999526272499015 | 0.0495627165466753 | -3.14942658670761 |
| **SESN2** | -0.615042433373784 | 7.68355534008214 | -2.63715969104088 | 0.0101090033406353 | 0.0499544563705623 | -3.19270221878287 |
| **ELL** | -0.573201185669051 | 9.38636014243196 | -2.68556380883025 | 0.00886399409894765 | 0.0458504442959954 | -3.2142595569534 |
| **MEF2D** | -0.36278214758077 | 10.7544110824689 | -2.70217247712157 | 0.00847008517766472 | 0.0450665848221119 | -3.28330055935208 |
| **SPARC** | -0.482940003741762 | 11.9467979502059 | -2.69693856111216 | 0.0085924616651755 | 0.0452599262803017 | -3.39707872923289 |
| **B2M** | 0.515988315298396 | 12.8040762661172 | 2.64299899923382 | 0.00995081561704323 | 0.0495627165466753 | -3.60006092800625 |
| **COL3A1** | -0.610563572672532 | 12.8716408911363 | -2.64509957134344 | 0.00989446285514674 | 0.0495627165466753 | -3.6002543446345 |

**Supplementary Table 3** - Metabolism-related genes covered by the F1RNA gene set

| | | | | | |
|---|---|---|---|---|---|
| ABCA6 | BLVRA | GALNT10 | LDHB | PLCB4 | SRD5A1 |
| ABCB1 | CA4 | GALNT12 | MTAP | PLPP3 | ST3GAL2 |
| ABCC2 | CD38 | GMPS | MTHFD1L | POLD1 | SULT1A1 |
| ABCC9 | CES1 | GPI | NAT1 | POLE | SUV39H2 |
| ACACA | CES2 | GSTM1 | NOS1 | PSAT1 | TAP1 |
| ACLY | CMPK2 | GUSB | NSD1 | PTEN | TAP2 |
| ACSL3 | CTPS1 | HDC | NT5C3A | PTGDS | TYMS |
| ACSL6 | CTPS2 | HSD11B1 | NT5E | PTGS2 | UGT8 |
| ADCY7 | DNMT1 | IDO1 | ODC1 | RRM2 | UPP1 |
| ALDH2 | DNMT3A | IMPDH1 | PAFAH1B2 | SCD | |
| ALOX12 | DOT1L | INPP1 | PHGDH | SCD5 | |
| ANPEP | ENTPD1 | ITPKB | PIK3CA | SDHA | |
| ARG2 | FUCA1 | KDSR | PIK3CD | SDHB | |
| ATIC | FUT4 | KMT5A | PIK3CG | SDHC | |
| B3GNT5 | FUT8 | KYNU | PLA2G7 | SDHD | |

**Supplementary Table 4** - Distribution of metabolism-related genes (covered by the F1RNA gene set) per metabolic pathway - Provided as a separate Word file.

**Supplementary Table 5** - Differentially expressed genes between UPS and high-grade LMS samples of an independent cohort (TCGA-SARC) - Provided as a separate CSV file.

**Supplementary Table 6** - Differentially expressed genes between UPS and DDLPS samples of an independent cohort (TCGA-SARC) - Provided as a separate CSV file.

**Supplementary Table 7 -** Demographic, pathological and clinical characteristics of the patients whose fresh normal and tumor tissue samples were used for $^{1}H$ NMR

| | **Total** (n=16) |
|---|---|
| **Age at diagnosis**, median [IQR], years | 74 [33.4 |
| **Gender**, n (%) | |
| Male | 5 (31.2) |
| Female | 11 (68.8) |
| **Histologic subtypes**, n (%) | |
| Undifferentiated pleomorphic sarcoma | 4 (25.0) |
| Leiomyosarcoma | 3 (18.8) |
| Liposarcoma | 2 (12.5) |
| Other histology* | 7 (43.7) |
| **Tumor grade**, n (%) | |
| G1 | 2 (12.5) |
| G2 | 5 (31.2) |
| G3 | 9 (56.3) |
| **Location**, n (%) | |
| Upper limb | 3 (18.8) |
| Lower limb | 7 (43.7) |
| Retroperitoneum | 3 (18.8) |
| Abdominal/Pelvic | 2 (12.5) |
| Trunk | 1 (6.2) |
| **Presentation type**, n (%) | |
| Localized | 13 (81.2) |
| Distant metastasis | 3 (18.8) |

*1 DFSP, 1 SFT, 1 synovial sarcoma,1 chondrosarcoma,1 CCSLGT, 1 low-grade endometrial stromal sarcoma, and 1 adamantinoma

**Supplementary Table 8 -** Surgical and systemic treatment specificities of the patients that underwent curative-intent surgery at the participating institutions and whose fresh normal and tumor tissue samples were used for $^1$H NMR

| | **Total** (n=13) |
|---|---|
| **Neoadjuvant treatment,** n (%) | |
| No neoadjuvant treatment | 9 (69.2) |
| Neoadjuvant treatment | 4 (30.8) |
| **Ressectability,** n (%) | |
| Resectable | 13 (100.0) |
| **Ressection margins,** n (%) | |
| R0/R1 | 13 (100.0) |
| **Adjuvant treatment,** n (%) | |
| Radiotherapy | 4 (30.8) |
| Chemotherapy | 1 (7.7) |
| No adjuvant treatment | 8 (61.5) |

### Supplementary Figures

**Supplementary Figure 1 -** Differentially expressed genes between LMS and DDLPS, between UPS and LMS and between UPS and DDLPS (including the whole range of genes included in the F1RNA gene set) (provided in larger size and higher resolution in a separate PPT file)

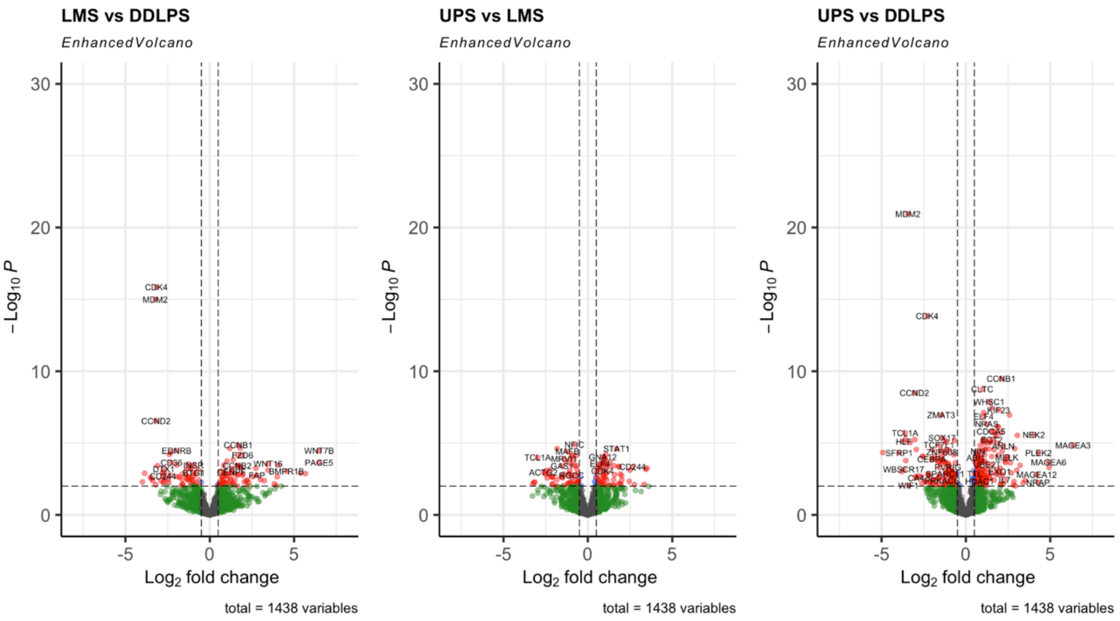

**Supplementary Figure 2** - F1RNA gene set coverage of metabolism-related pathways and genes and distribution of metabolism-related genes per metabolic pathway (provided in larger size and higher resolution in a separate PPT file)

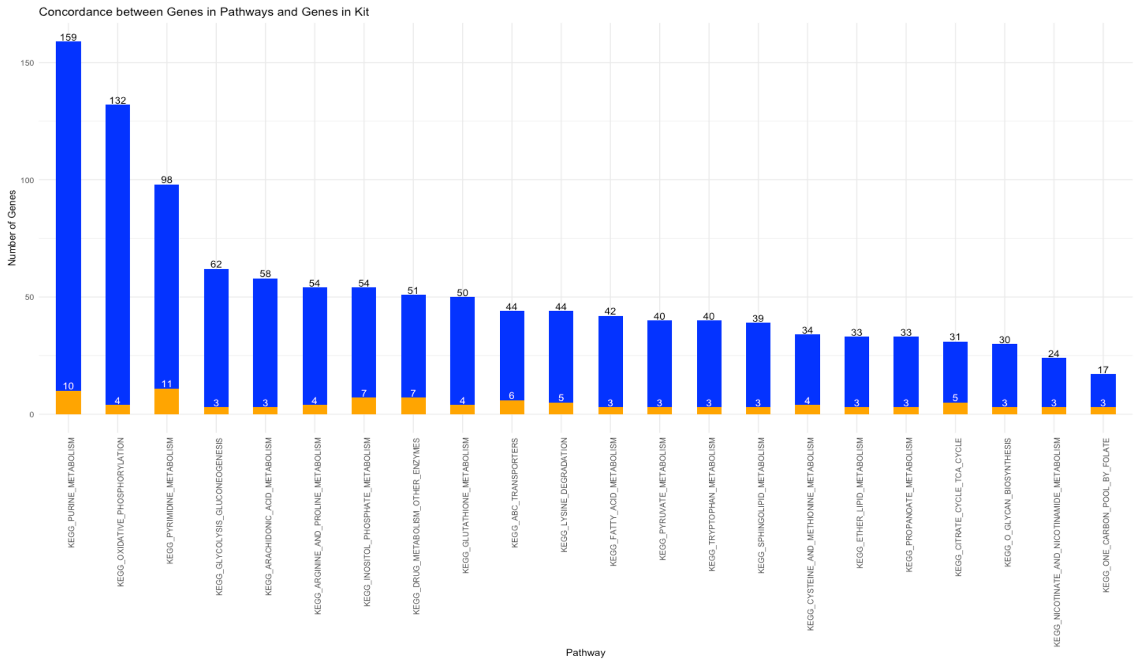

**Supplementary Figure 3** - Differentially expressed metabolism-related genes between LMS and DDLPS, between UPS and LMS and between UPS and DDLPS (provided in larger size and higher resolution in a separate PPT file)

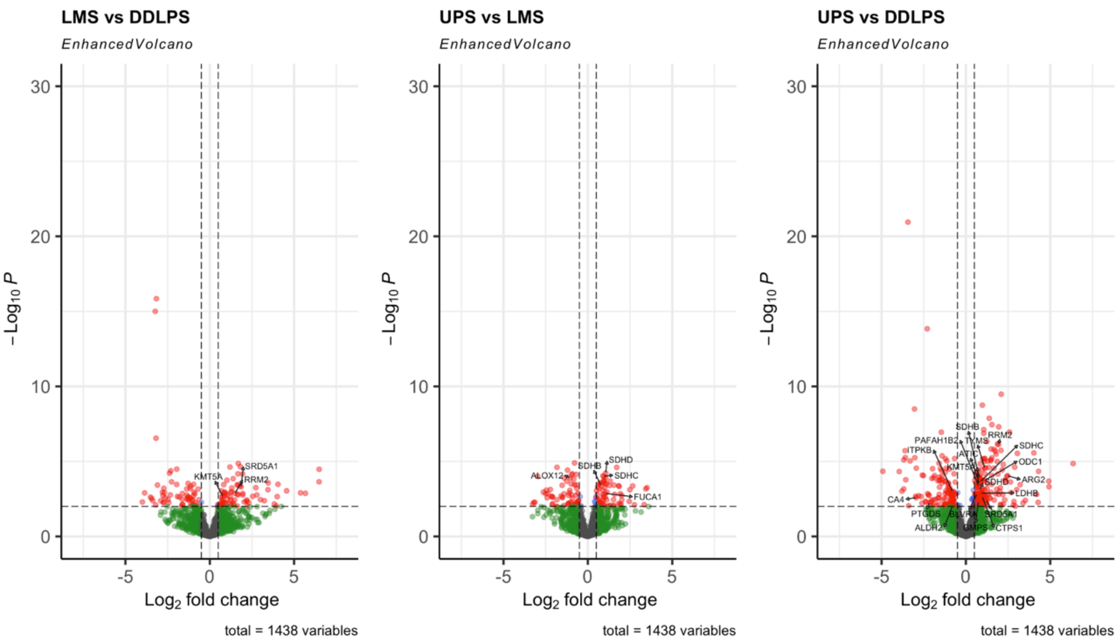

**Supplementary Figure 4** - Distribution of differentially expressed (between LMS and DDLPS, between UPS and LMS and between UPS and DDLPS) metabolism-related genes per represented metabolic pathway (provided in larger size and higher resolution in a separate PPT file)

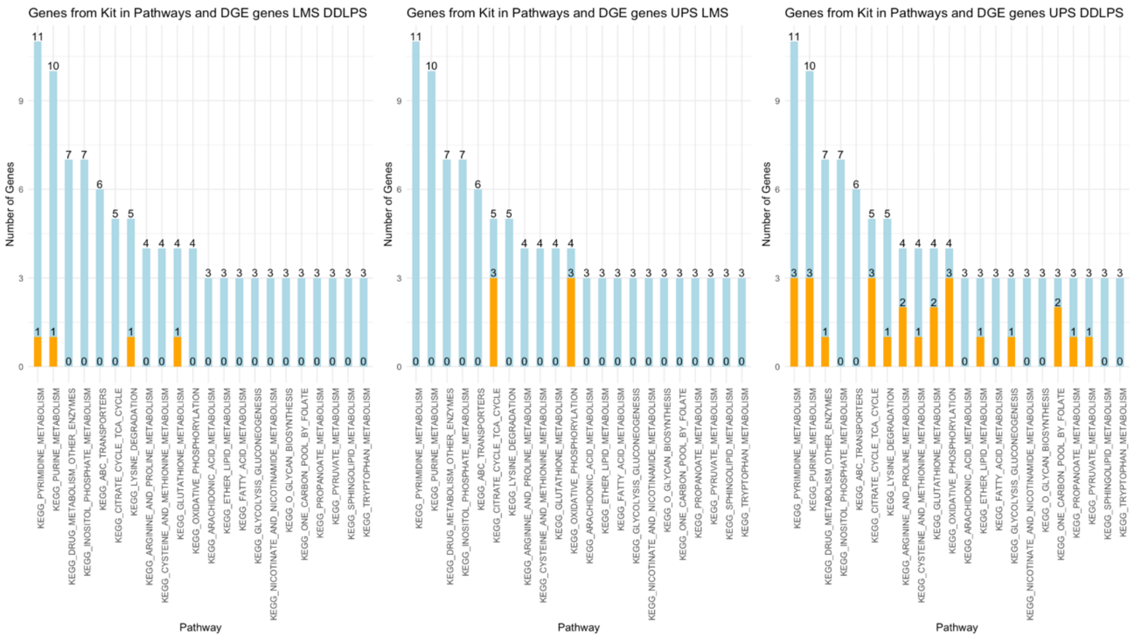

**Supplementary Figure 5** - Correlations, within the UPS population, between the over expression of SDHB and MFS, RFS and PFS.

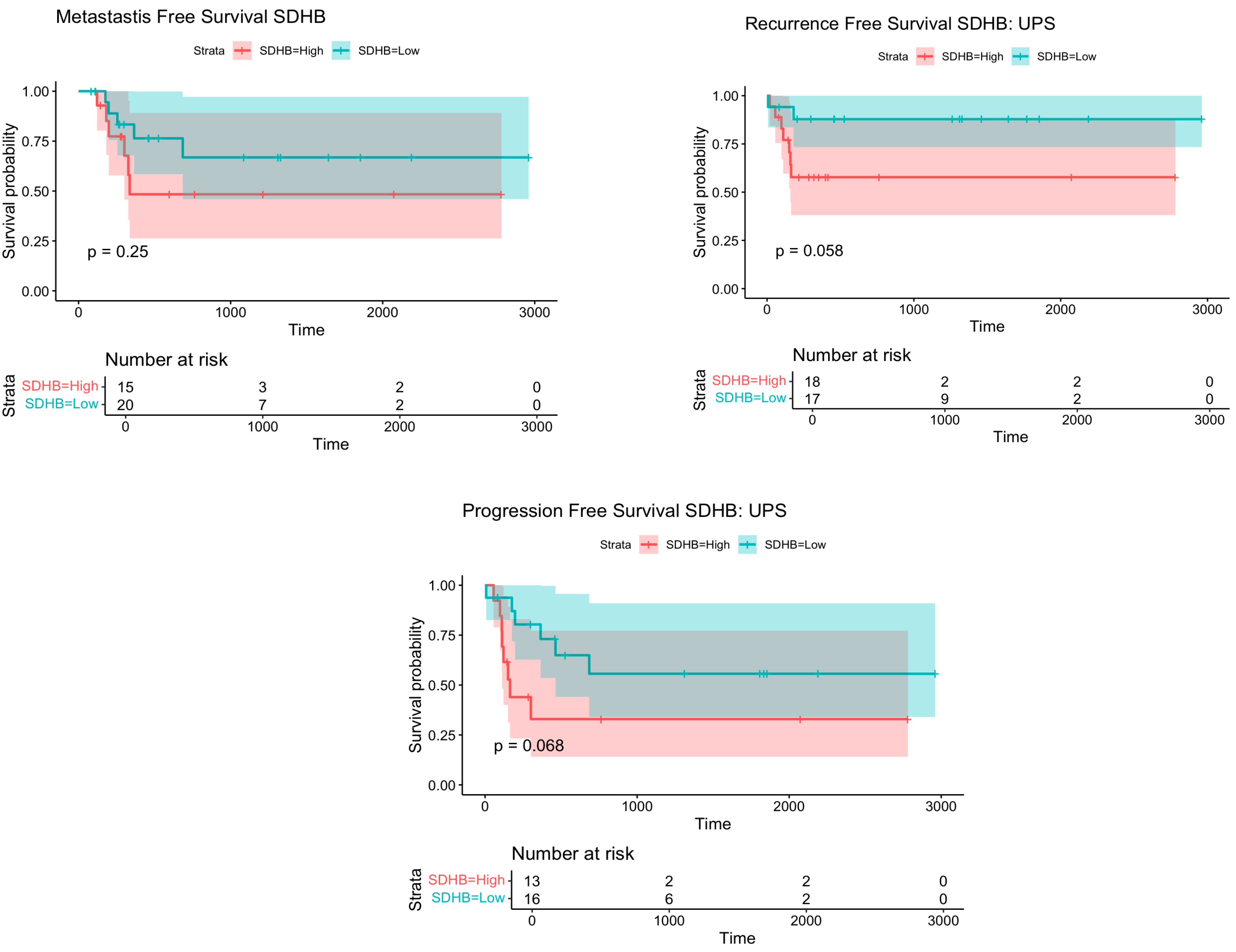

**Supplementary Figure 6** - Correlations, within the UPS population, between the over expression of SDHC and OS, MFS, RFS and PFS.

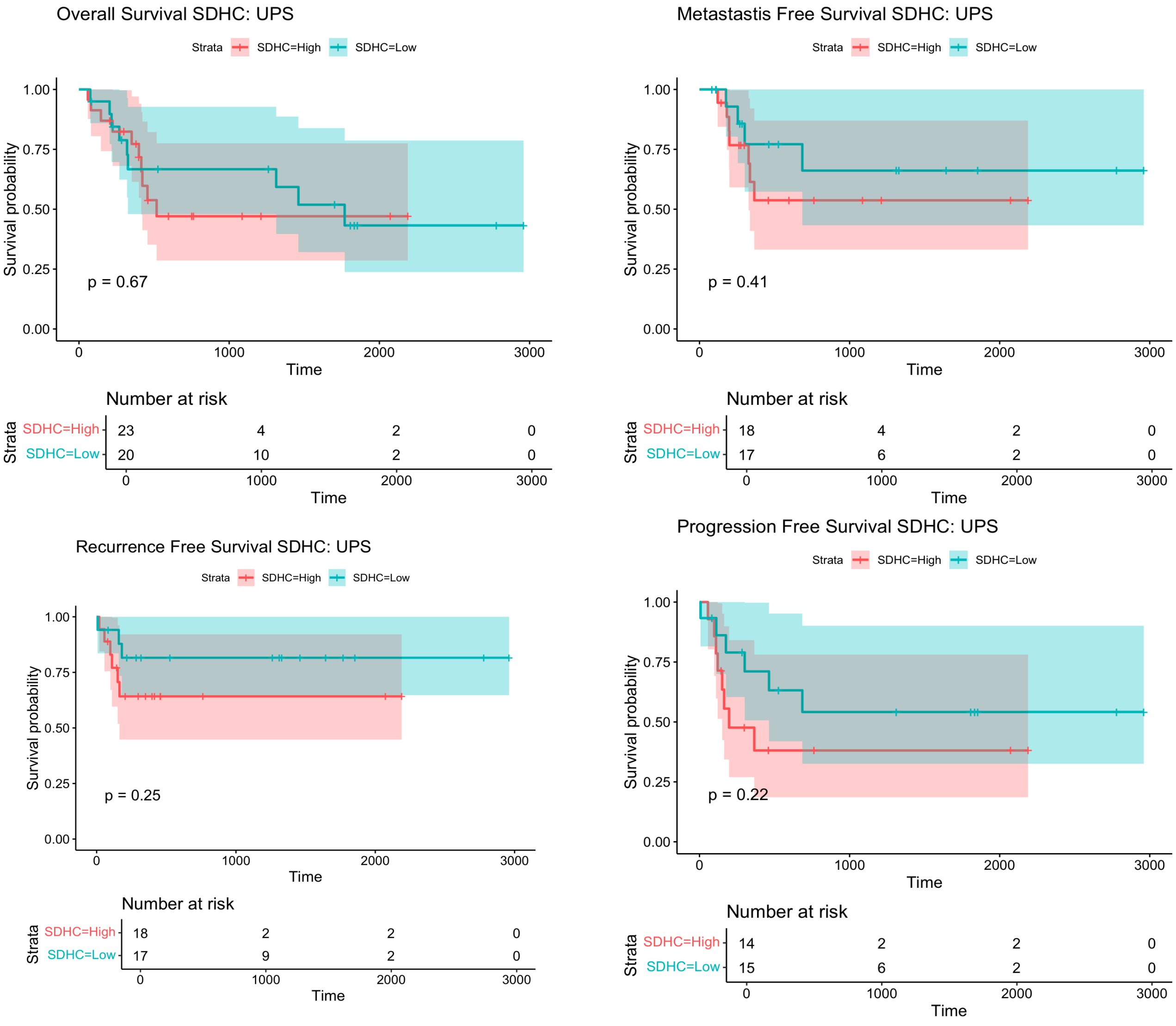

**Supplementary Figure 7** - Correlations, within the UPS population, between the over expression of SDHD and OS, MFS, RFS and PFS.

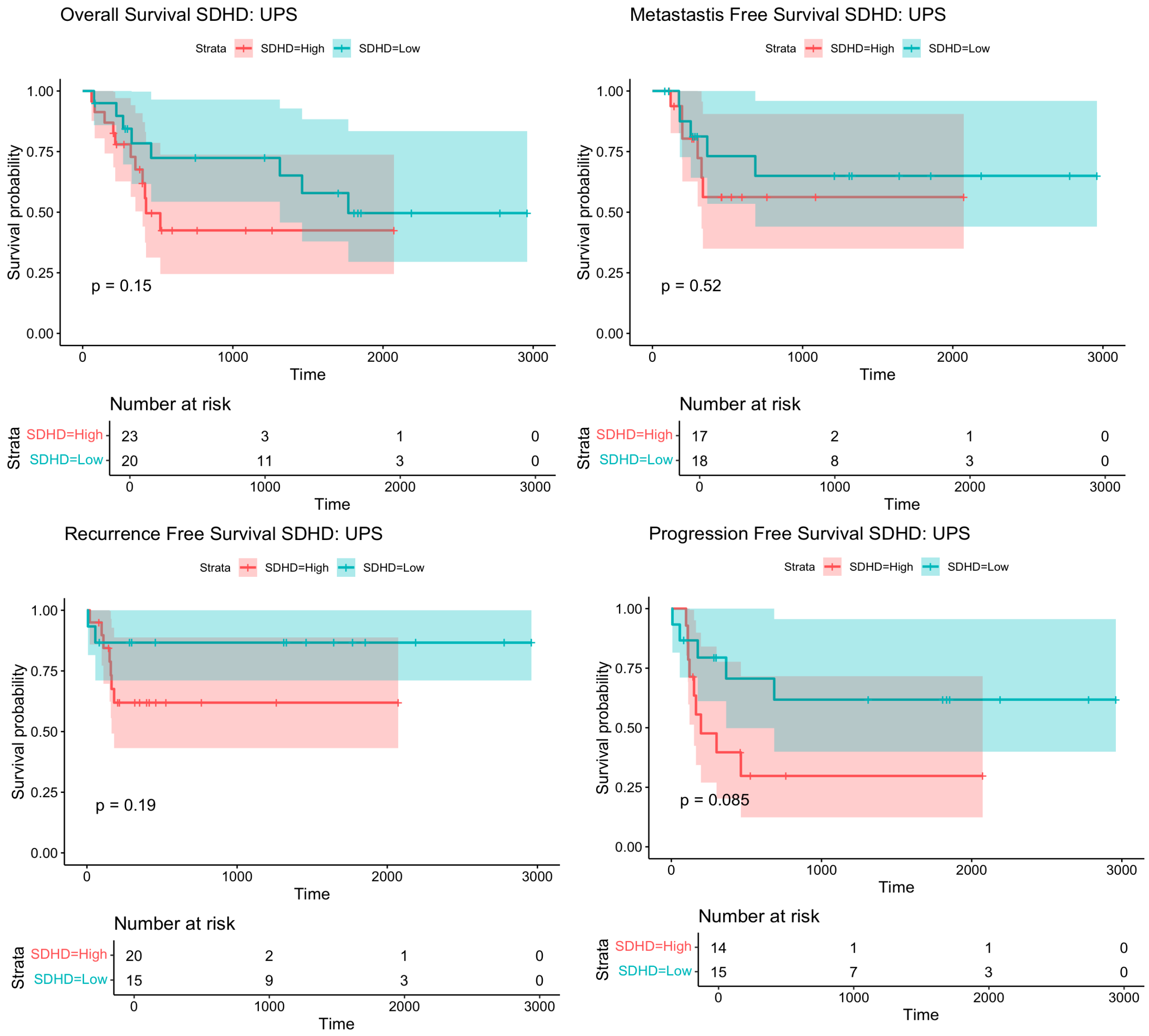

**Supplementary Figure 8** - Correlations, in the whole cohort population, between the over expression of SDHB and OS using the KMM (Supplementary Figure 8a) and using a CPHM (Supplementary Figure 8b).

### Supplementary Figure 8a)

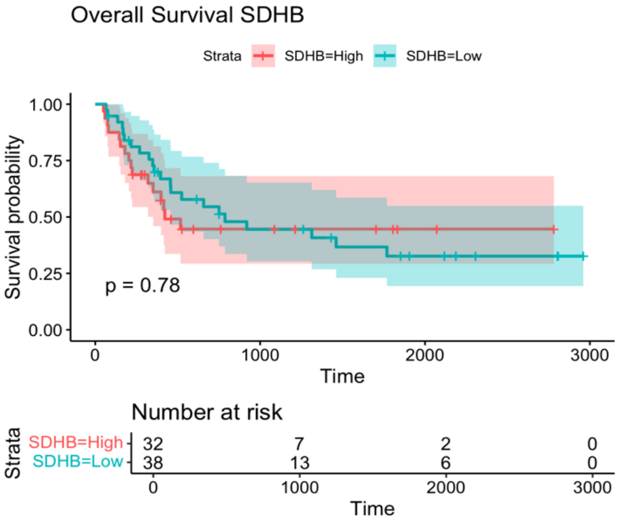


### Supplementary Figure 8b)

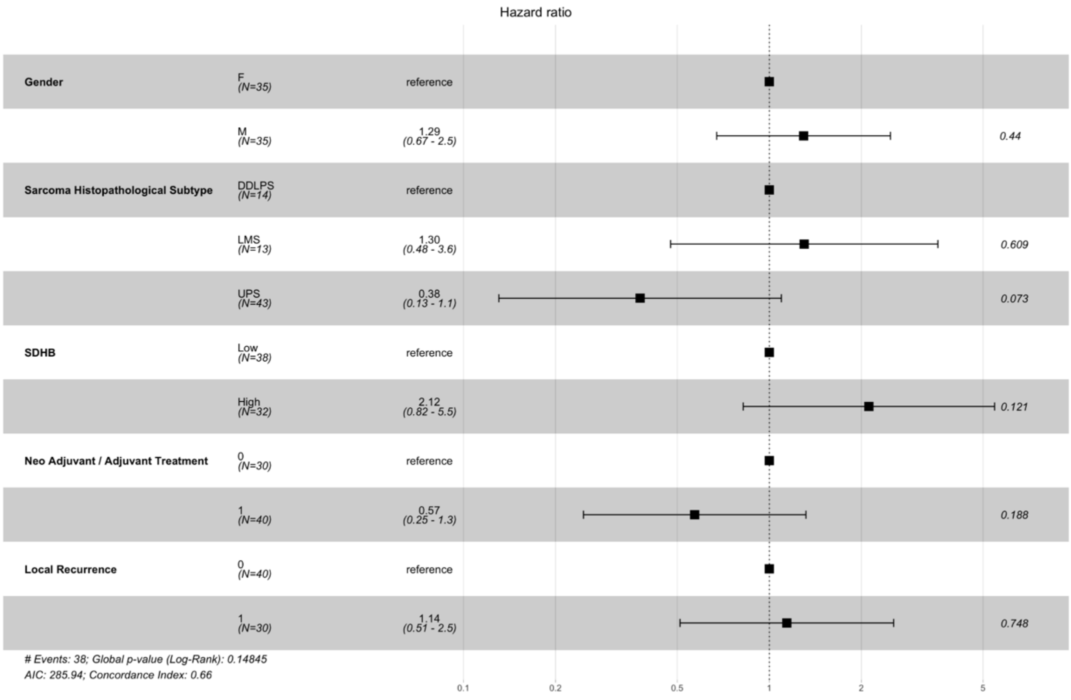

**Supplementary Figure 9** - Correlations, in the whole cohort population, between the over expression of SDHB and MFS, RFS and PFS.

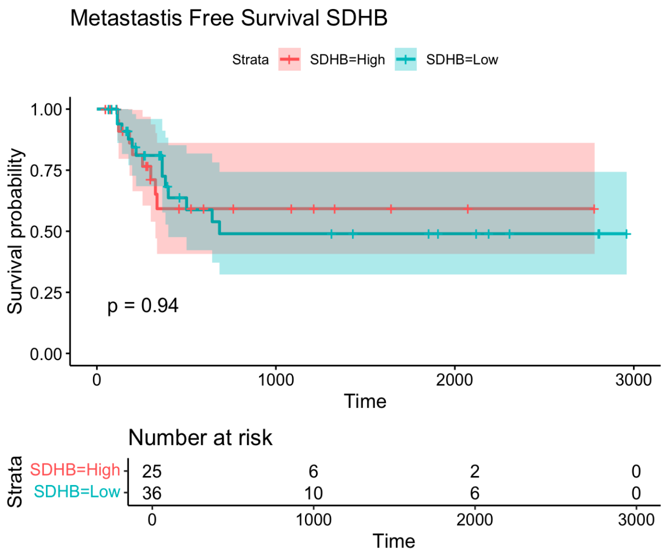


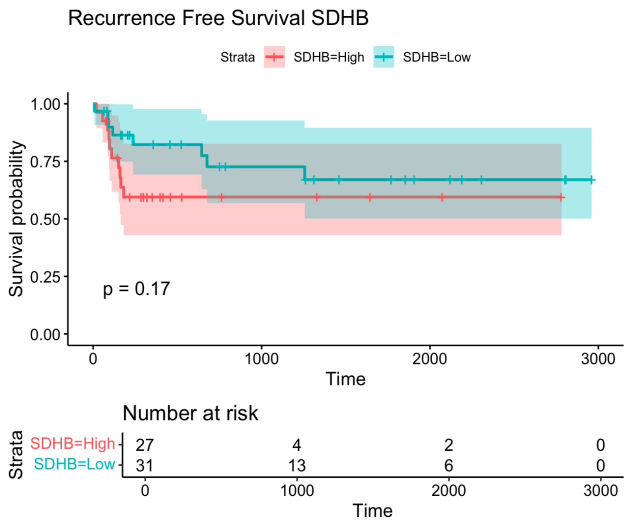


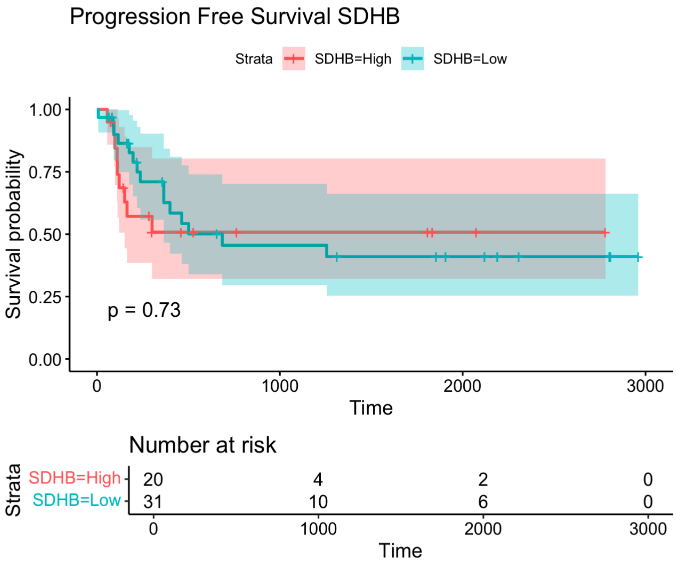

**Supplementary Figure 10** - Correlations, in the whole cohort population, between the over expression of SDHC and OS, MFS, RFS and PFS.

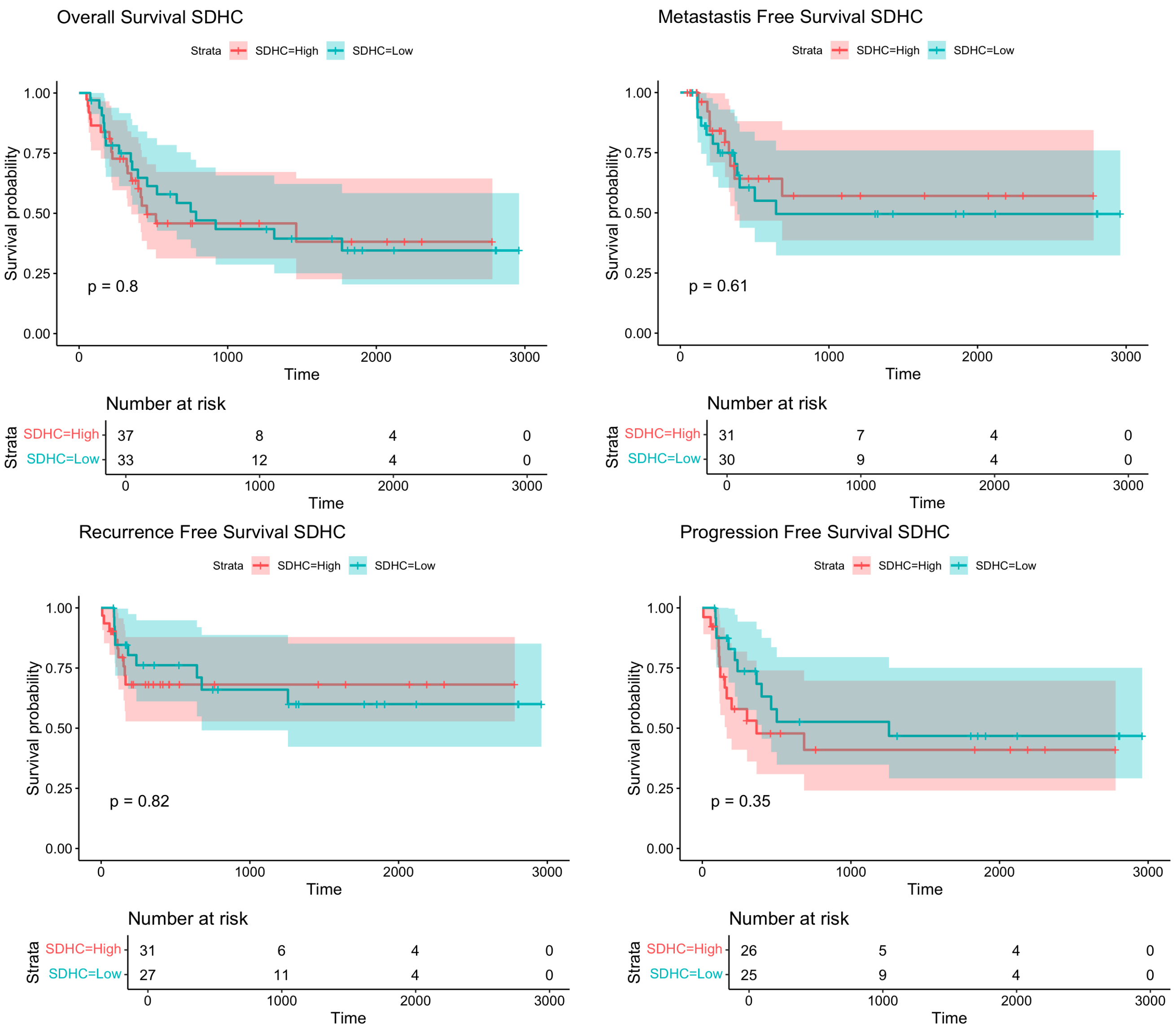

**Supplementary Figure 11** - Correlations, in the whole cohort population, between the over expression of SDHD and OS, MFS, RFS and PFS.

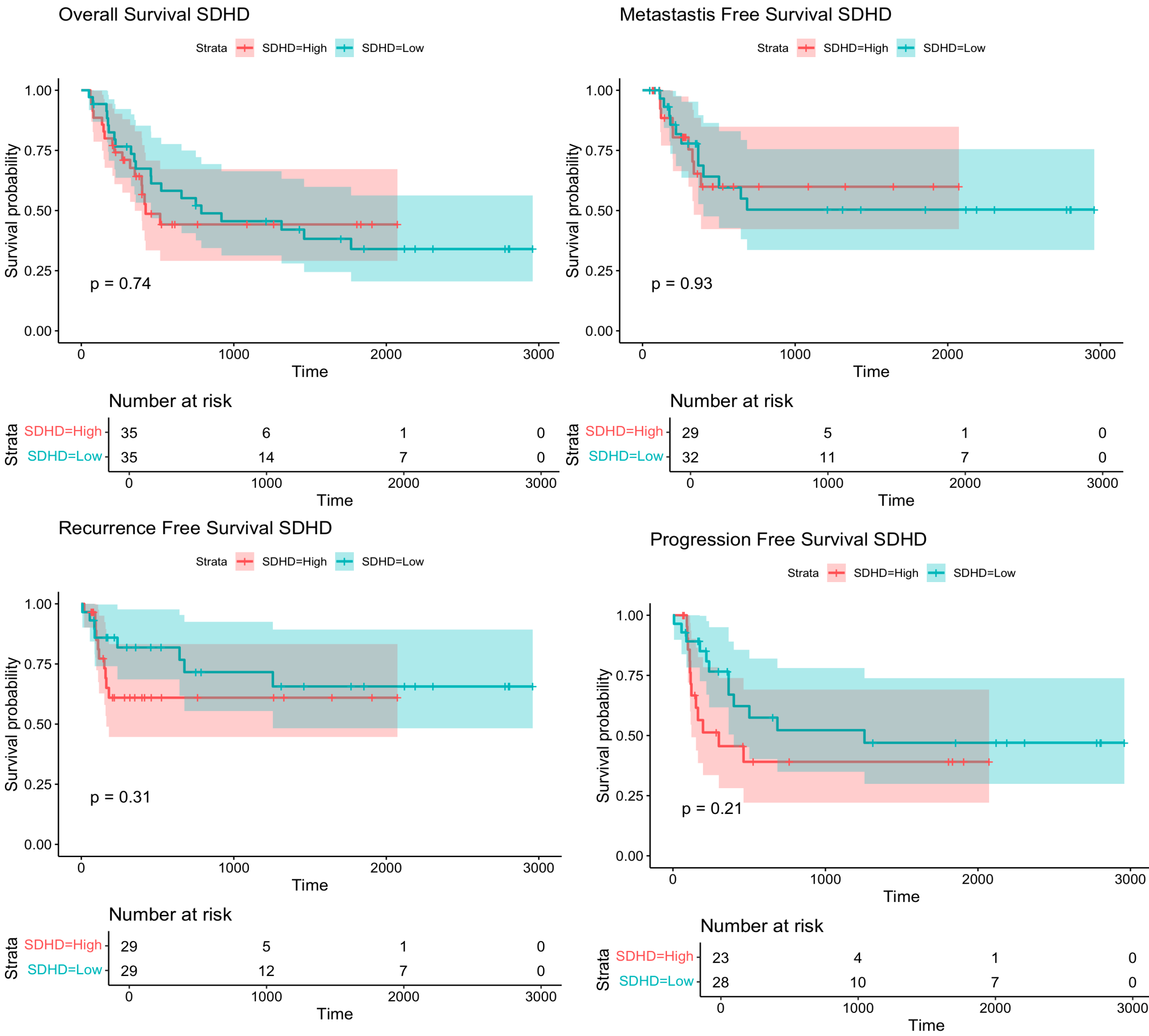

**Supplementary Figure 12** - Differentially expressed genes between LMS and DDLPS, between UPS and LMS and between UPS and DDLPS, using samples of an independent cohort (TCGA-SARC) (provided in larger size and higher resolution in a separate PPT file)

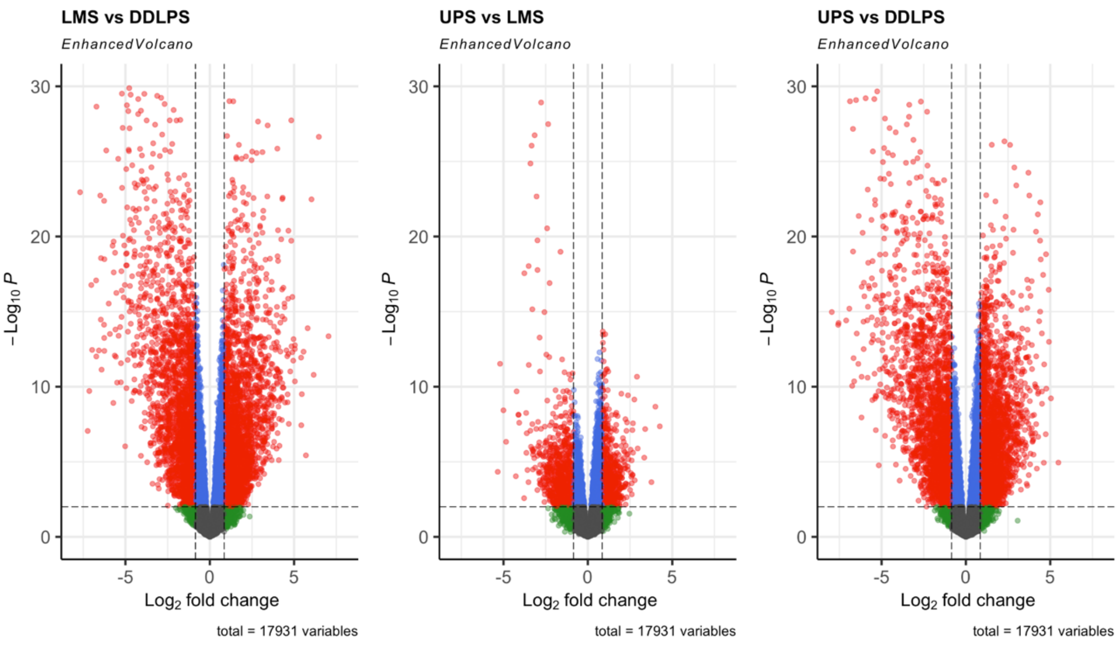

**Supplementary Figure 13** - 15 pathways whose differential over expression, between UPS and LMS samples of an independent cohort (TCGA-SARC) is more significant

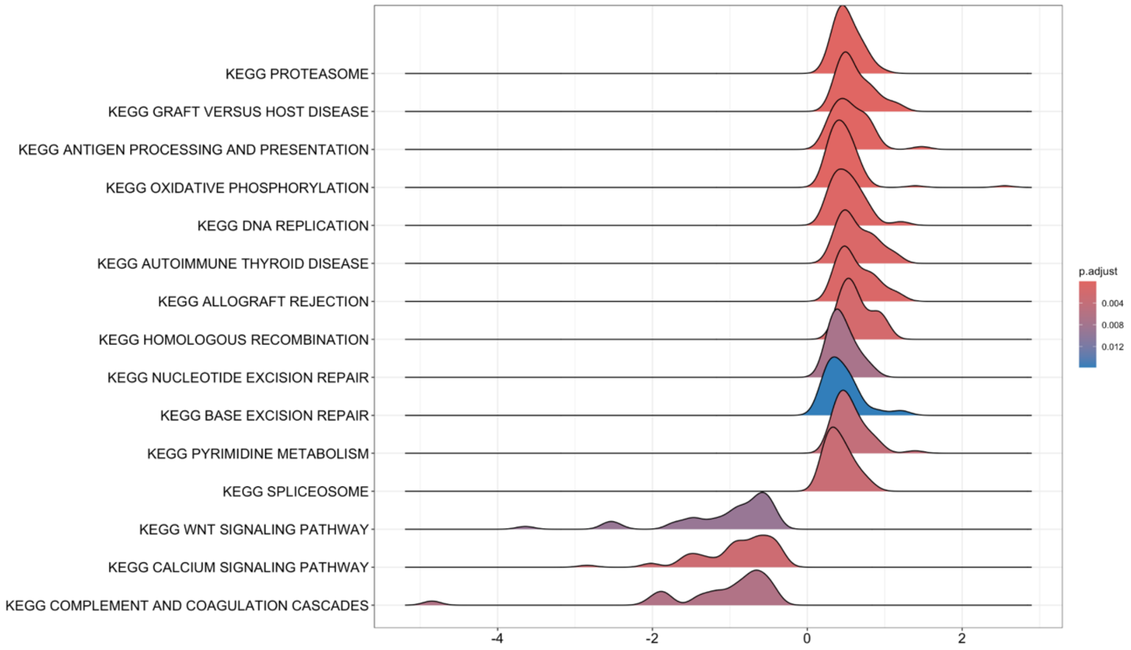

**Supplementary Figure 14** - Differentially expressed metabolism-related pathways between LMS and LPS samples, between UPS and LMS samples and between UPS and LPS samples of an independent cohort (TCGA-SARC) (provided in larger size and higher resolution in a separate PPT file)

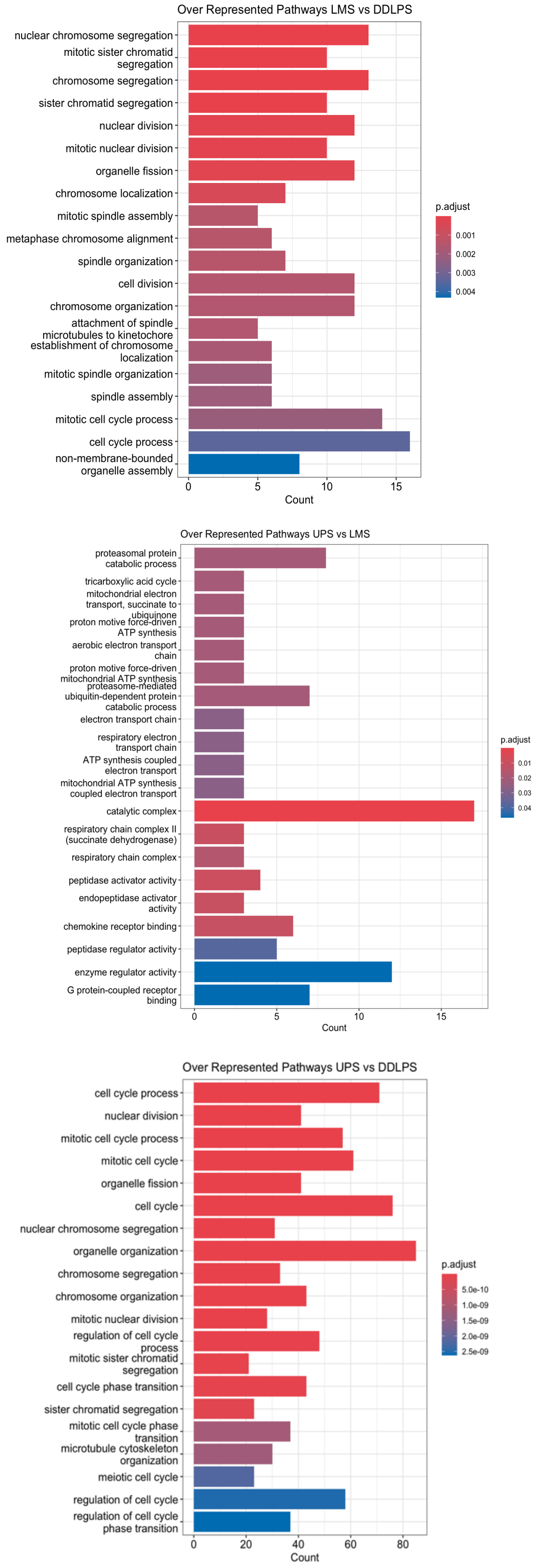

**Supplementary Figure 15** - Comparison between the logFC of differentially expressed genes both in the study and in the independent cohort, in order to determine directional concordance (UPS vs. LMS)

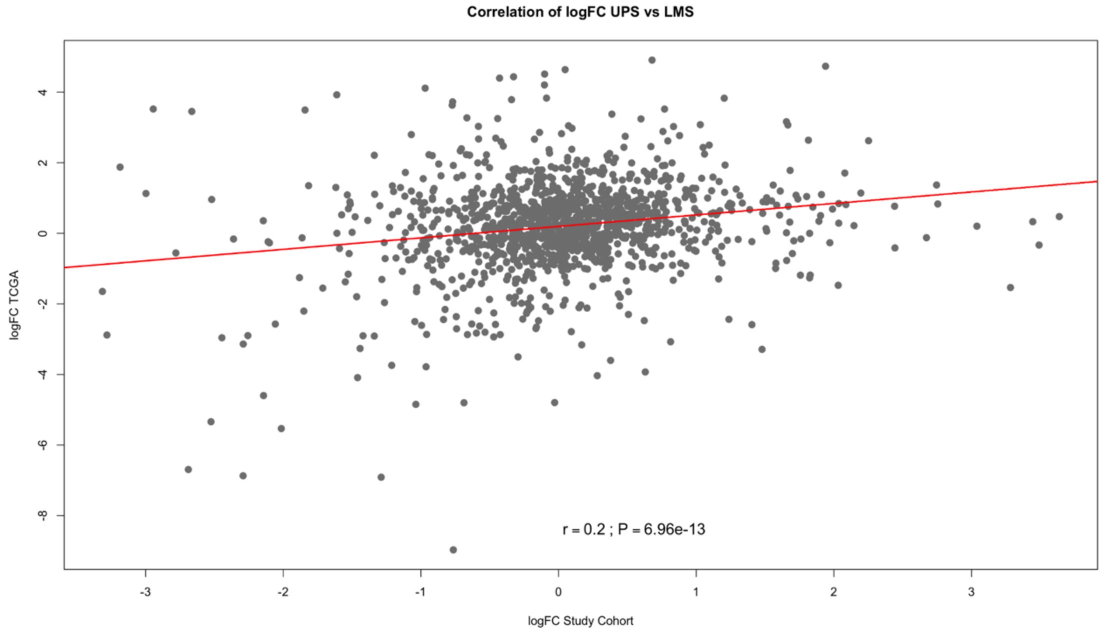

**Supplementary Figure 16** - Comparison between the logFC of differentially expressed genes both in the study and in the independent cohort, in order to determine directional concordance (UPS vs. DDLPS)

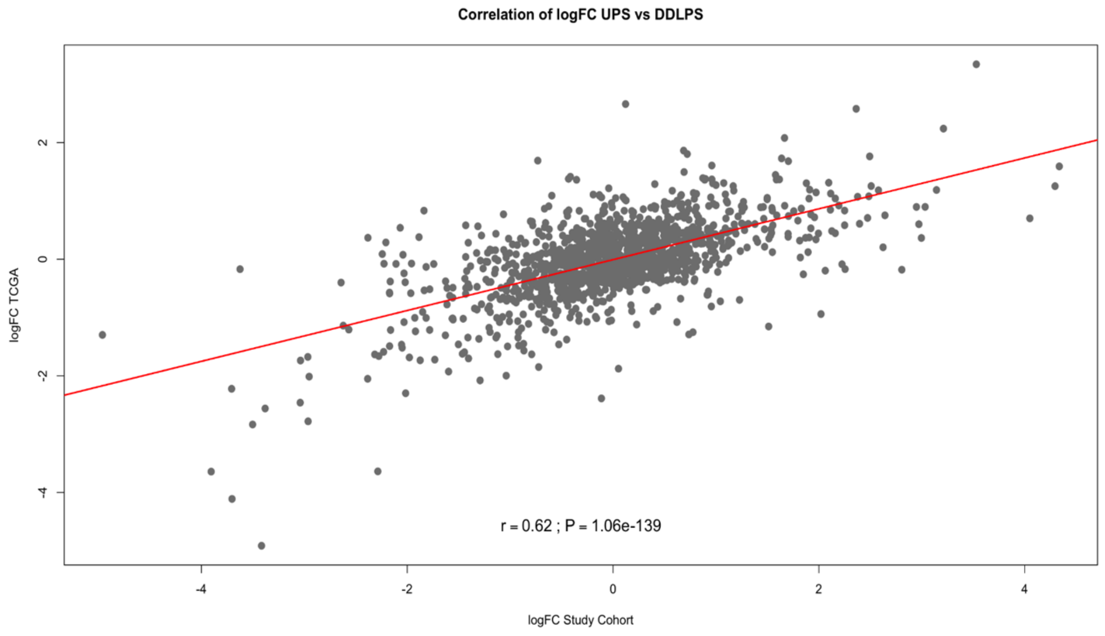

**Supplementary Figure 17** - Comparison between the logFC of differentially expressed genes both in the study and in the independent cohort, in order to determine directional concordance (LMS vs. DDLPS)

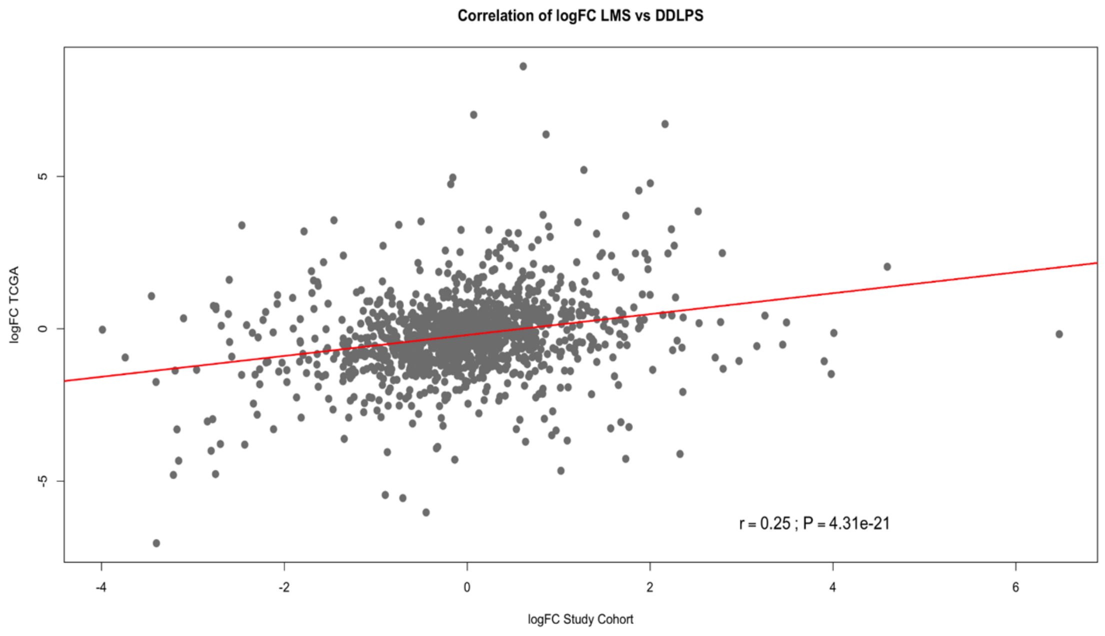

**Supplementary Figure 18** - Evaluation of the statistical significance of directional agreement (in terms of differential gene expression) for all comparisons (UPS vs. LMS; UPS vs. DDLPS; LMS vs. DDLPS) based on Chi-square tests ((provided in larger size and higher resolution in a separate PPT file)

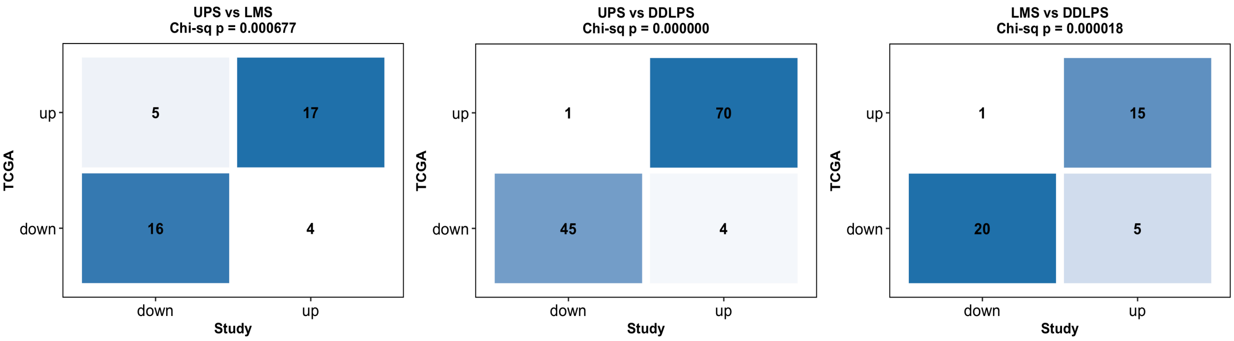

**Supplementary Figure 19** - Evaluation of the statistical significance of the survival direction concordance for UPS samples (of the study cohort and of the TCGA-SARC cohort) based on Chi-square tests ((provided in larger size and higher resolution in a separate PPT file)

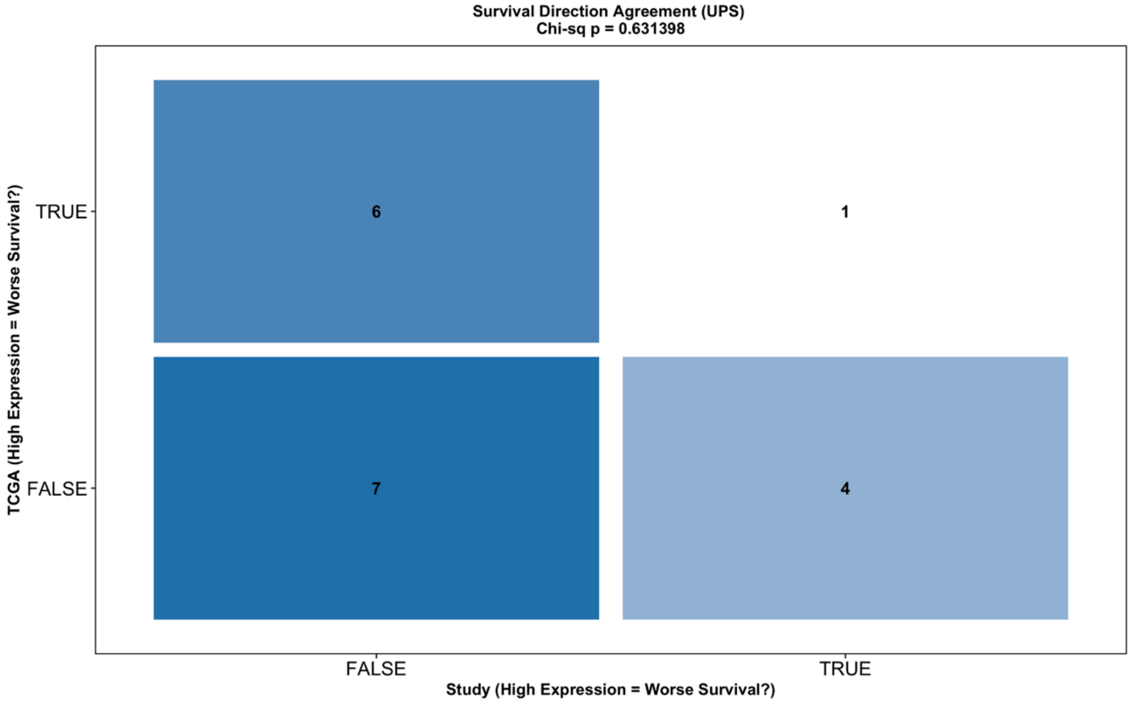

**Supplementary Figure 20** - Evaluation of the statistical significance of the survival direction concordance for the UPS, LMS and DDLPS samples (of the study cohort and of the TCGA-SARC cohort) based on Chi-square tests (provided in larger size and higher resolution in a separate PPT file)

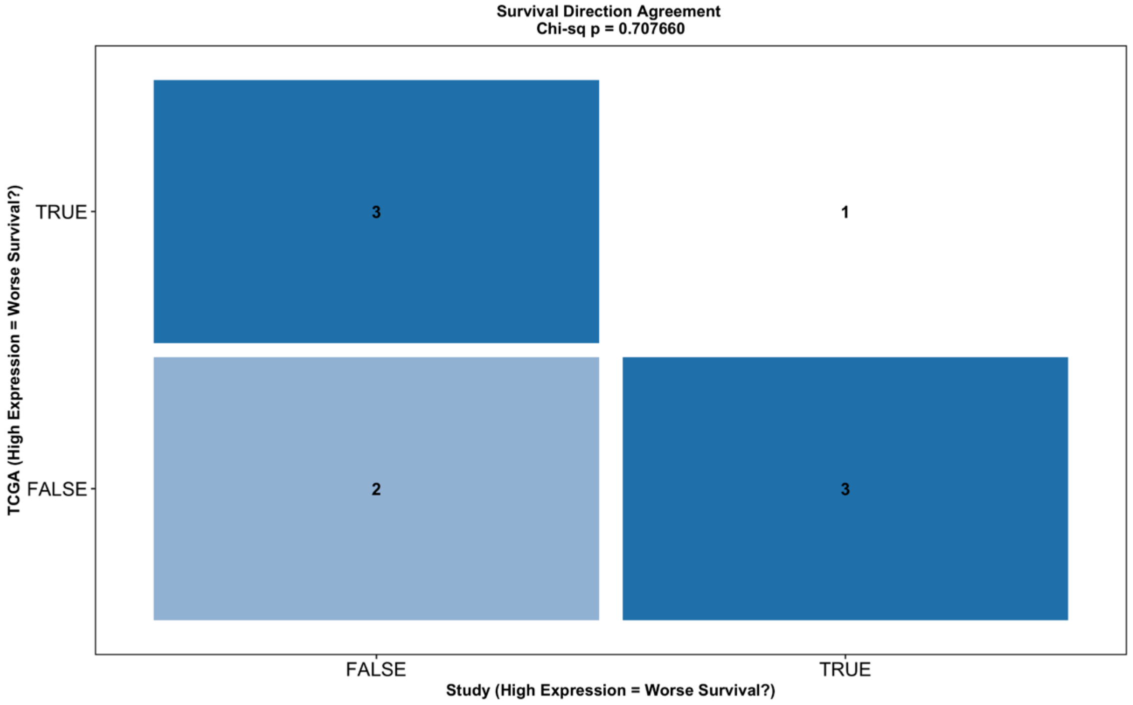

**Supplementary Figure 21** - Pathways whose over expression is correlated with the expression of genes whose high expression levels are correlated with improved prognosis within the universe of UPS samples of the TCGA-SARC cohort

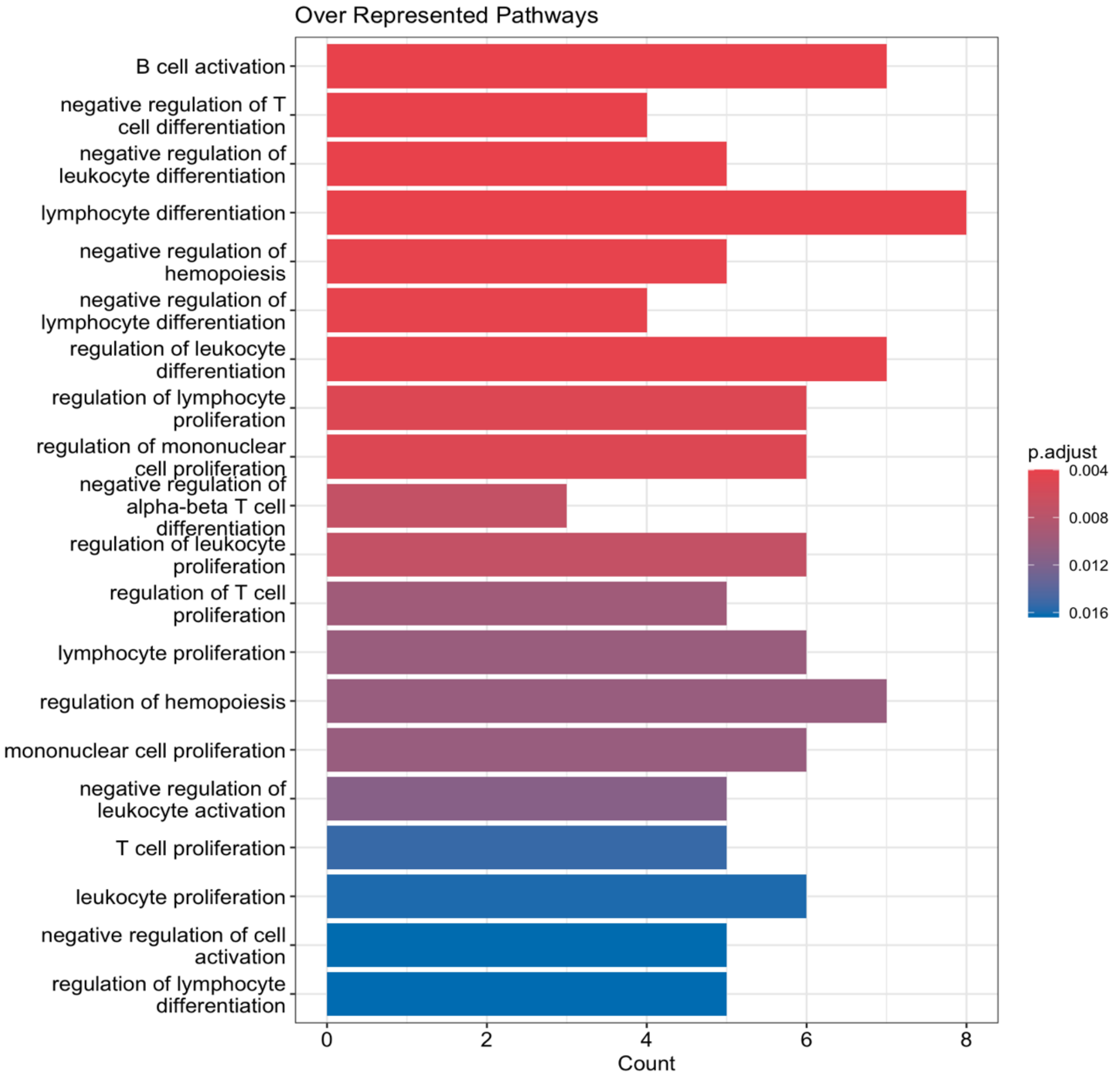

**Supplementary Figure 22** - Pathways whose over expression is correlated with the expression of genes whose high expression levels are correlated with improved prognosis within the universe of UPS samples of the study cohort

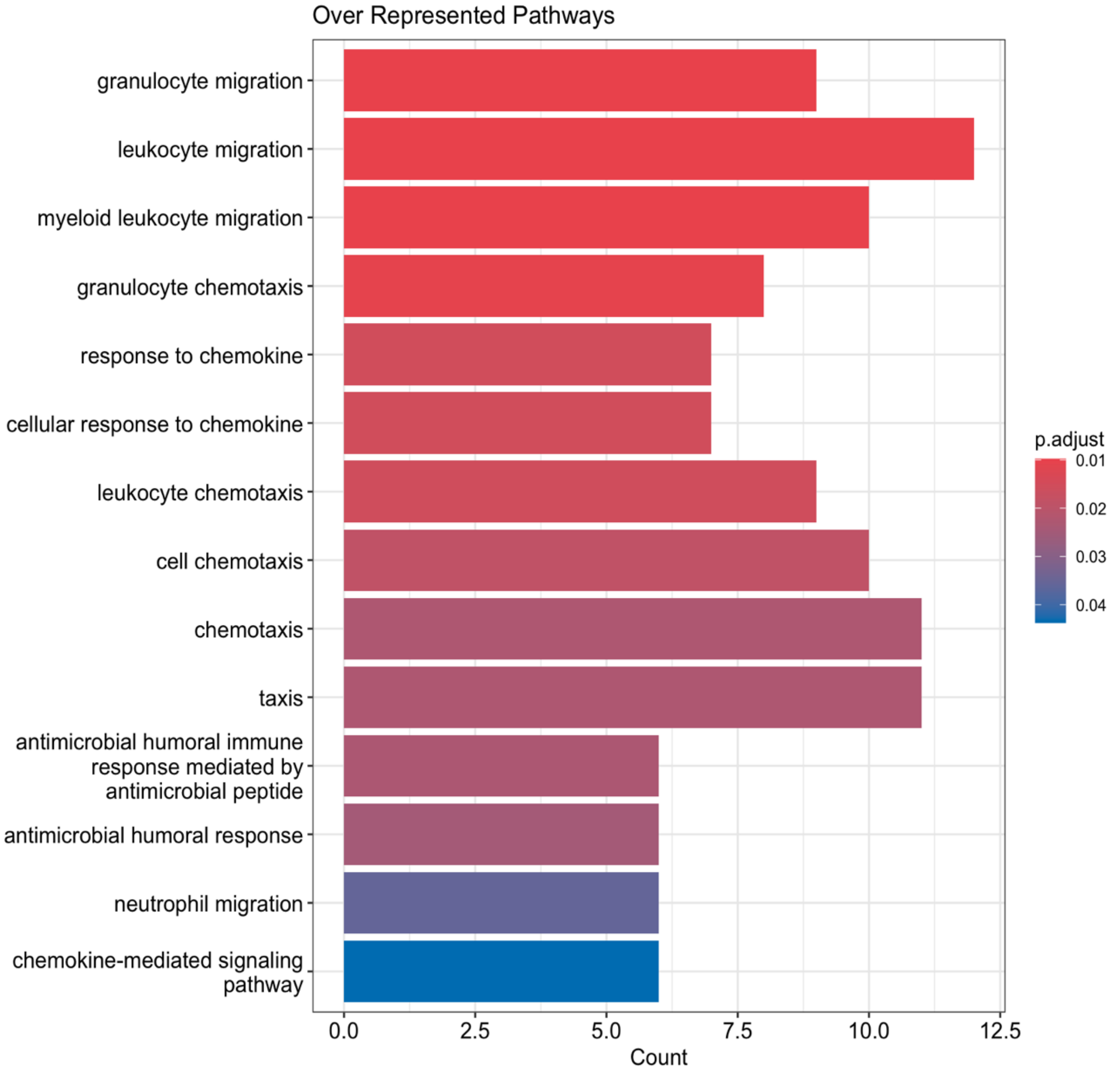

**Supplementary Figure 23** - Evaluation of the degree of coverage by the F1RNA gene set of the genes whose expression shapes the immune-related pathways expression patterns within the TCGA-SARC cohort (provided in larger size and higher resolution in a separate PPT file)

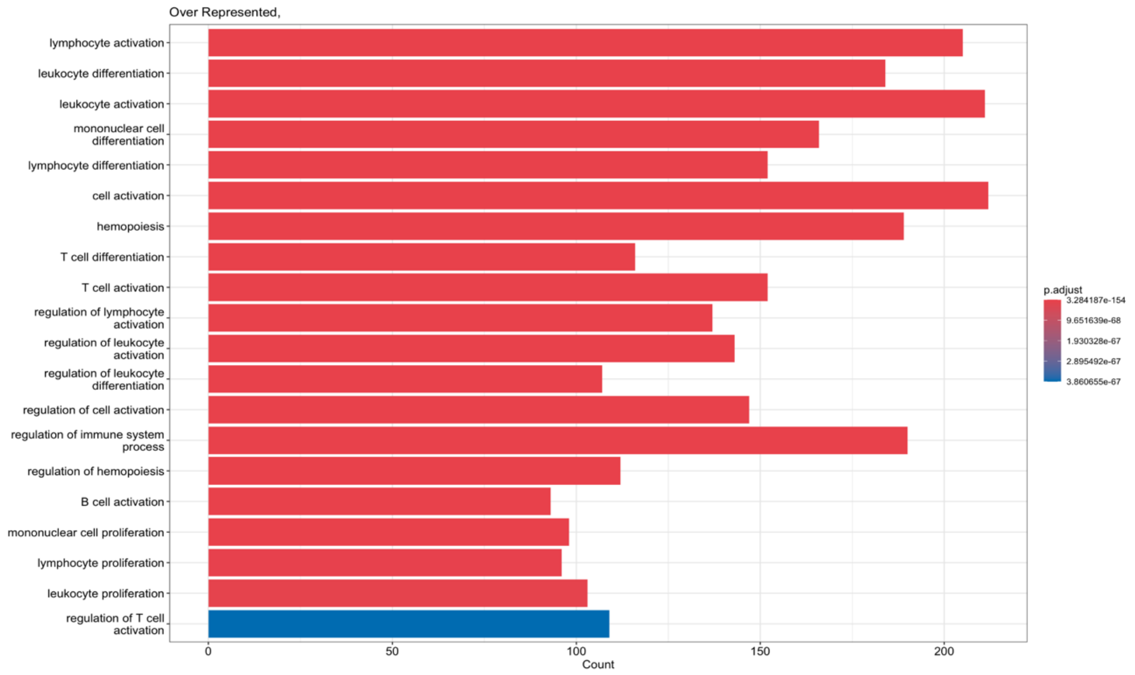

**Supplementary Figure 24** - Comparison between the number of indels per patient across different STS histotypes in the study cohort

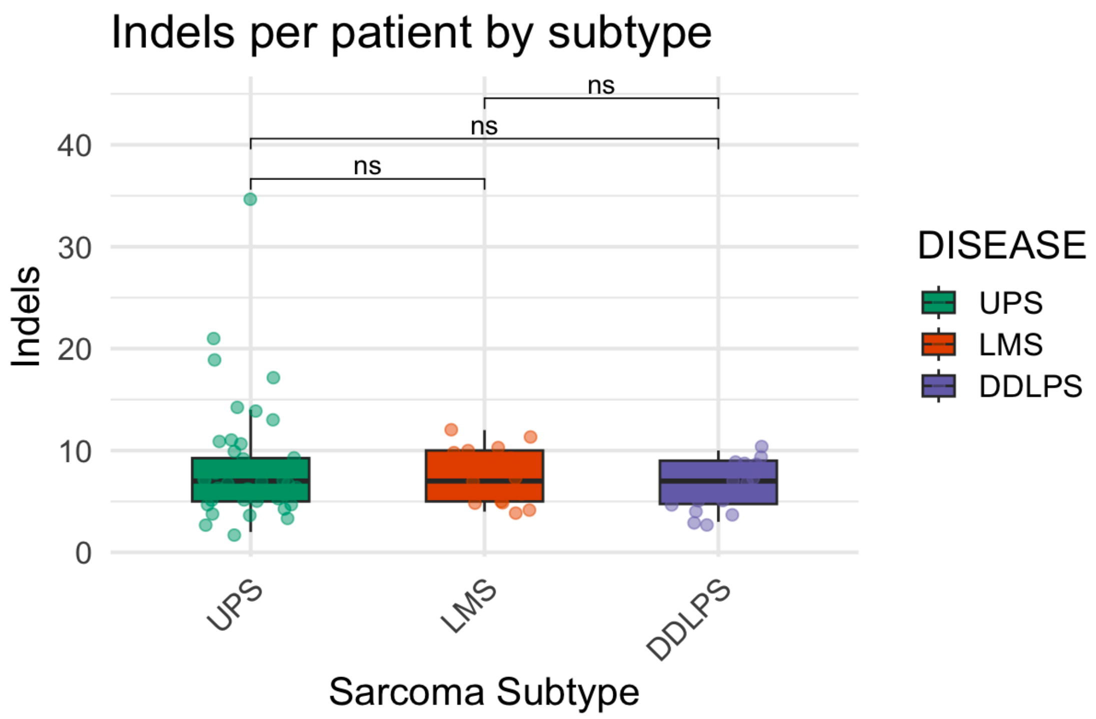

**Supplementary Figure 25** - Comparison between the number of frameshift indels per patient across different STS histotypes in the study cohort

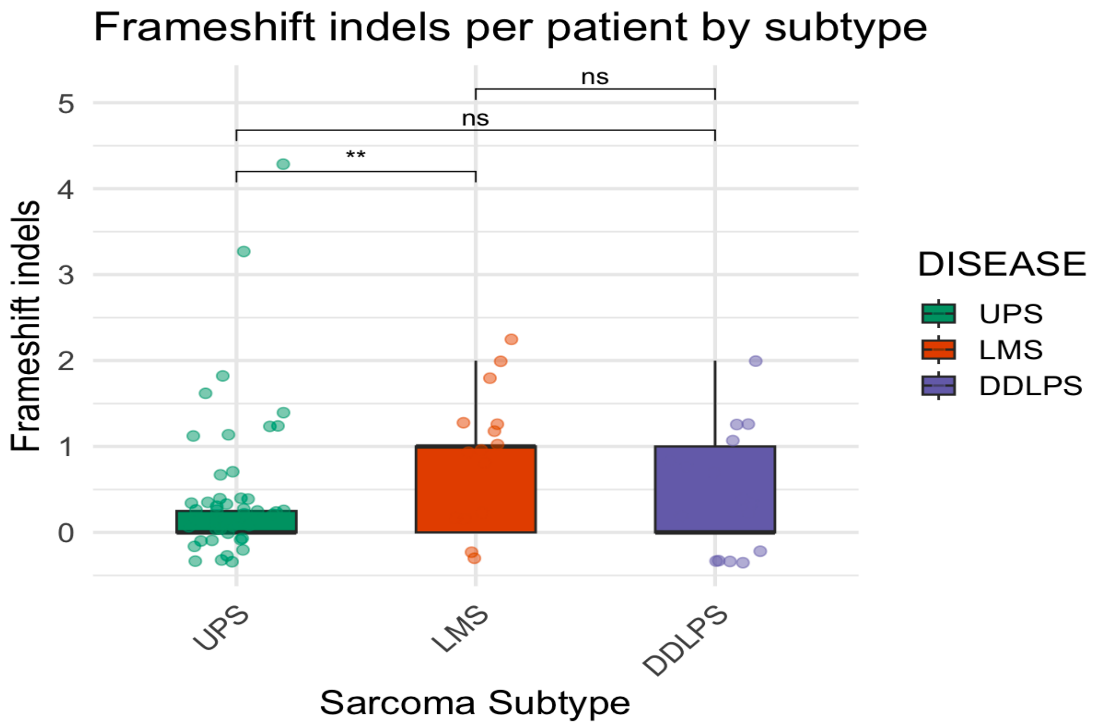

**Supplementary Figure 26** - Comparison between the number of indels per patient across different STS histotypes in the TCGA-SARC cohort

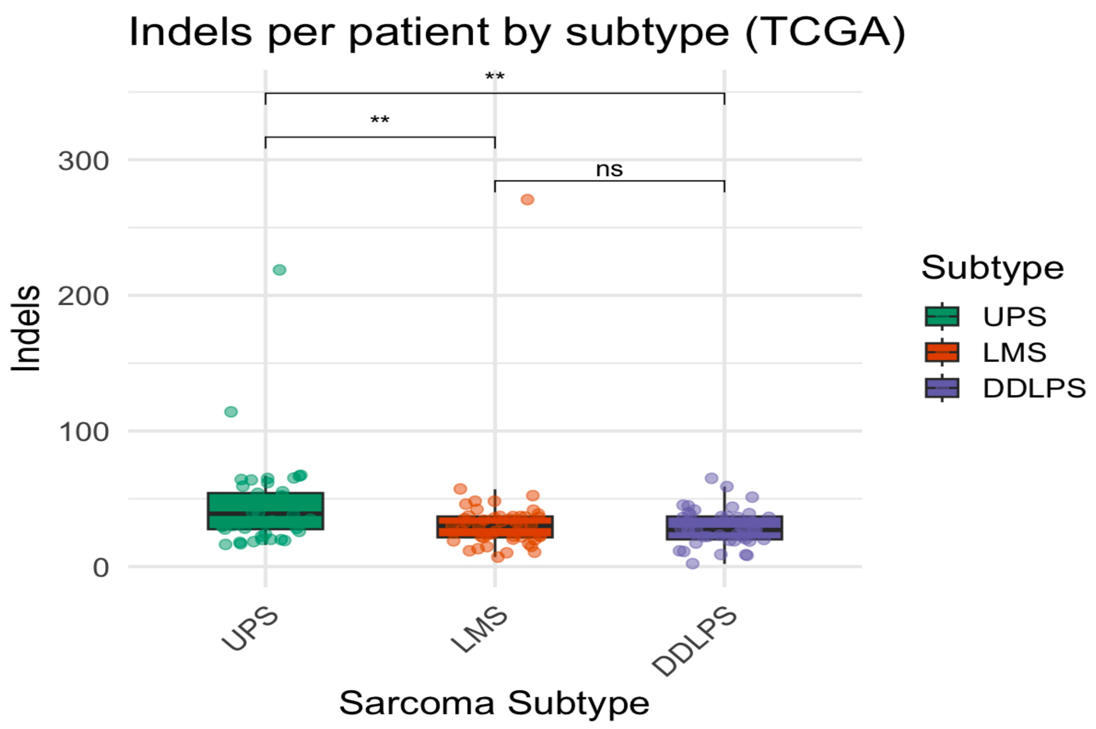

**Supplementary Figure 27** - Comparison between the number of frameshift indels per patient across different STS histotypes in the TCGA-SARC cohort

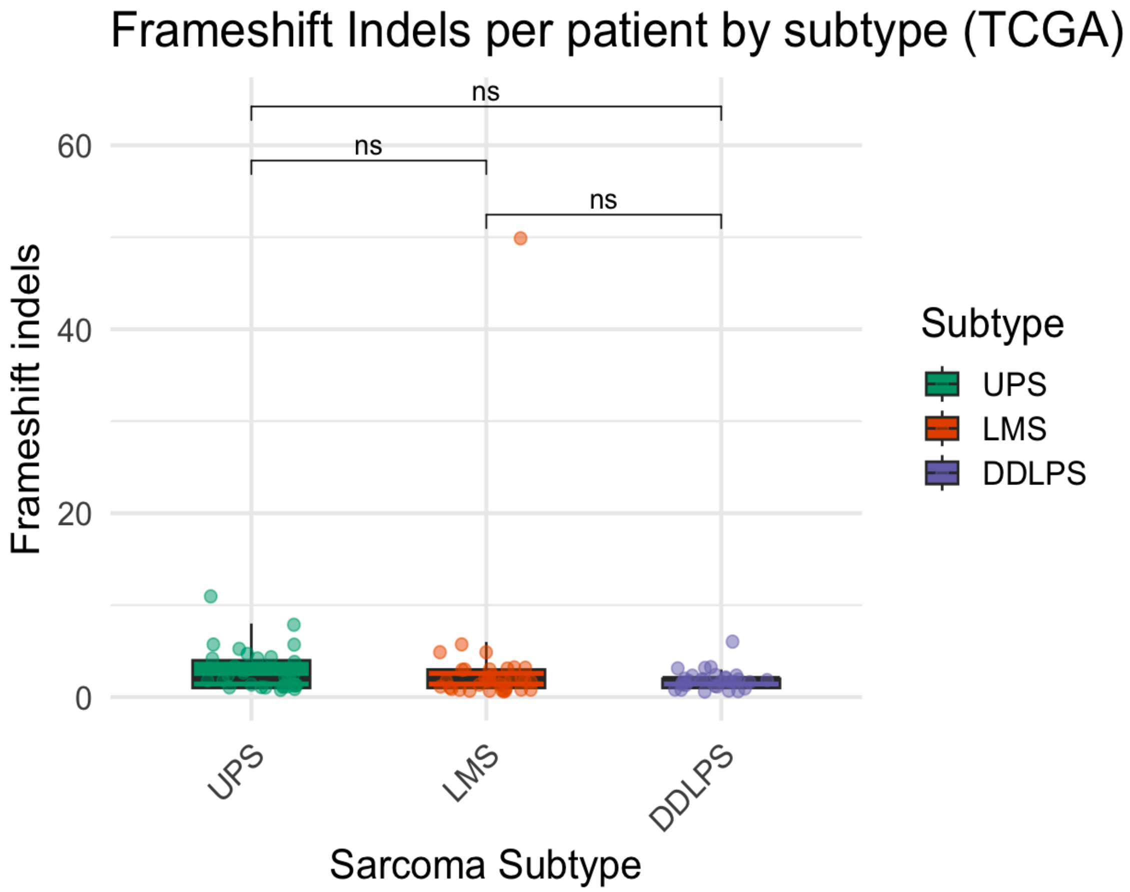
